# ATP13A4 gates extracellular polyamine levels to control excitatory synaptogenesis

**DOI:** 10.1101/2025.04.04.25325117

**Authors:** Sarah van Veen, Emily Meeus, Dolores Irala, Kristina Sakers, Zhaolin Liu, Justin Savage, Gabrielle Séjourné, Dhanesh Sivadasan Bindu, Elke Ausloos, Hanna Elzbieta Grzesik, Hanne Dhondt, Nina Schoonvliet, Chris Van den Haute, Joris Van Asselberghs, Marta Montpeyó Garcia-Moreno, Keimpe Wierda, Veerle Baekelandt, Konrad Platzer, Kevin Rostasy, Kai Lee Yap, Jan Eggermont, Matthew Holt, Cagla Eroglu, Peter Vangheluwe

## Abstract

Polyamines such as spermidine and spermine are critical regulators of brain development, yet the mechanisms controlling their cellular uptake and extracellular availability remain poorly defined. Here, we identify ATP13A4, a P5B-type ATPase enriched in glia with prominent expression in astrocytes, as a key determinant of brain polyamine homeostasis. Using biochemical, cellular, and *in vivo* approaches, we show that ATP13A4 drives cellular polyamine uptake and constrains extracellular polyamine levels. Loss of ATP13A4 reduces astrocyte morphological complexity and enhances astrocyte-driven excitatory synapse formation. Exogenous spermidine recapitulates these effects, identifying extracellular spermidine as a synaptogenic cue and implicating astrocytic ATP13A4 in regulating extracellular polyamine availability. *In vivo*, constitutive *Atp13a4* knockout mice exhibit a striking redistribution of brain polyamines, with reduced cortical levels and accumulation in cerebrospinal fluid, indicating disrupted compartmentalization between intra-and extracellular pools. These changes are accompanied by increased excitatory synapse number and synaptic activity, as well as delayed early postnatal neurodevelopment followed by mild, female-biased behavioral alterations in adulthood. Furthermore, we identify rare ATP13A4 variants associated with neurodevelopmental disorders and show that these variants impair transporter function. Together, our findings reveal that astrocyte-mediated polyamine clearance via ATP13A4 shapes extracellular spermidine availability, thereby regulating excitatory synaptogenesis throughout neurodevelopment.

## Introduction

Polyamines, such as spermidine and spermine, are abundant organic polycations with dynamic roles in various biological processes (1), and are dysregulated in aging (2, 3), cancer (4), and brain disorders (5, 6). They function as anti-oxidants and anti-inflammatory molecules, and are critical for fundamental cellular processes such as protein synthesis, membrane stability, signal transduction, neurotransmission, autophagy, and cell growth (reviewed in (7)). Maintaining proper polyamine homeostasis in the brain is essential not only for neurodevelopment but also for neuronal survival and network stability (8–13). Polyamines shape neuronal connectivity and circuit formation through their modulation of ion channels, receptors, and influence on synaptic plasticity (14–17). While balanced levels confer neuroprotective benefits (18, 19), disruptions in polyamine homeostasis – whether through excessive accumulation or depletion – can lead to neuronal death, heightened seizure susceptibility, or other forms of neurological dysfunction (5, 6, 10, 11, 13, 20–24). Supporting this, mutations in polyamine synthesis genes are often associated with neurodevelopmental disorders (25–30), highlighting the critical need for tightly regulated polyamine levels during brain development.

Within the brain, spermidine and spermine are produced predominantly by neurons but accumulate mainly in astrocytes, the peri-synaptic glial cells of the central nervous system (31–35). Astrocytes have been proposed to actively clear polyamines from the extracellular space, preventing excessive polyamine accumulation and potential toxicity, while also being capable of releasing polyamines to modulate neuronal activity (31, 32, 36, 37). This bidirectional exchange may position astrocytes as key regulators of extracellular polyamine tone, yet the molecular identity of the astrocytic polyamine uptake system has remained unknown.

Astrocytes are central regulators of synapse development and circuit function. They maintain excitatory-inhibitory balance by buffering extracellular neurotransmitters and ions (38–40), and actively promote synapse formation and maturation through secreted synaptogenic factors and direct contact with pre-and postsynaptic elements via their elaborate arborized morphology (41–53). During early postnatal development, astrocytes acquire this complex morphology in parallel with neuronal synapse formation (45, 54–61). In turn, neuronal cues such as synaptic activity and cell-cell adhesion instruct astrocytes’ morphological and functional maturation, revealing a rich and bidirectional signaling between astrocytes and neurons that governs brain development (52, 59, 62–68). Whether polyamines, whose extracellular levels are controlled by astrocytic uptake, participate in this signaling has not been investigated.

P5B-type ATPases are emerging as a novel class of polyamine transporters (5, 69–73), shedding light on the long-standing mystery of the mammalian polyamine transport system. Among these ATPases, ATP13A2 is the most extensively studied, with well-defined substrate specificity and transport mechanisms (5, 74–79). In contrast, ATP13A3 and ATP13A4 have been implicated in cellular polyamine uptake (69–73), but their precise transport mechanism and substrate specificities remain poorly defined, leaving significant gaps in our understanding of their functional roles.

Here, we identify ATP13A4 as a polyamine transporter that constrains extracellular polyamine availability in the brain. We demonstrate that ATP13A4-mediated uptake in astrocytes regulates both astrocyte morphogenesis and excitatory synaptogenesis, and that extracellular spermidine acts as a synaptogenic cue linking polyamine homeostasis to circuit development. *Atp13a4* knockout mice exhibit disrupted brain polyamine compartmentalization and delayed early neurodevelopment, whereas in humans, rare ATP13A4 variants with impaired transporter function are associated with neurodevelopmental disorders, establishing disease relevance. Together, these findings establish glial polyamine clearance via ATP13A4 as a previously unrecognized mechanism linking extracellular polyamine homeostasis to excitatory synaptogenesis and neurodevelopmental disease.

## Results

### ATP13A4 is expressed in astrocytes during postnatal development

P5B-type transport ATPases have recently emerged as key determinants of cellular polyamine uptake. To identify which isoform(s) may mediate polyamine uptake in astrocytes, we systematically analyzed human and mouse transcriptomic datasets spanning multiple brain regions, cell types and developmental stages (**Supplementary Fig. 1-2**). ATP13A4 shows a clear astrocyte-enriched expression profile, with stronger astrocyte specificity in mouse as compared to humans where also expression in oligodendrocyte precursor cells (OPCs) can be found (80–90). Notably, *msAtp13a4* is one of 825 commonly expressed genes highly enriched in astrocytes across 13 different regions of the central nervous system (CNS) (91) (**Supplementary Fig. 2f**), suggesting a core function for msAtp13a4 in these cells. Comparative analysis of the P5B ATPase family indicates that ATP13A4 is the only member with pronounced astrocytic enrichment, identifying it as a primary candidate for mediating polyamine uptake in this cell type. The developmental increase in ATP13A4 expression during late fetal and early postnatal stages further supports a role in astrocyte-dependent processes during postnatal brain development.

To validate astrocytic *Atp13a4* expression in rodent models, we isolated astrocytes and neurons from postnatal day 1 (P1) rat cortices (**Fig. 1a**) and confirmed strong enrichment of rat *Atp13a4* (*rAtp13a4*) mRNA in cortical astrocytes compared to neurons (**Fig. 1b**). Next, we verified astrocytic *msAtp13a4* mRNA expression during postnatal cortical development (P7-21) using multiplex RNA fluorescence *in situ* hybridization (FISH) in Aldh1L1-eGFP transgenic mice, in which all astrocytes are labeled with eGFP (**Fig. 1c**). Across cortical layers (L1, L2/3, L4, L5) and developmental stages (P7, P15, P21), the majority of *msAtp13a4* puncta were localized within eGFP-positive astrocytes relative to neighboring GFP-negative cells (**Fig. 1d**), consistent with transcriptomic datasets showing astrocyte-enriched expression (**Supplementary Fig. 2**). Together, these findings demonstrate that Atp13a4 is expressed in astrocytes during early postnatal brain development, in line with its glia-enriched expression in mouse and human datasets.

**Fig. 1:**
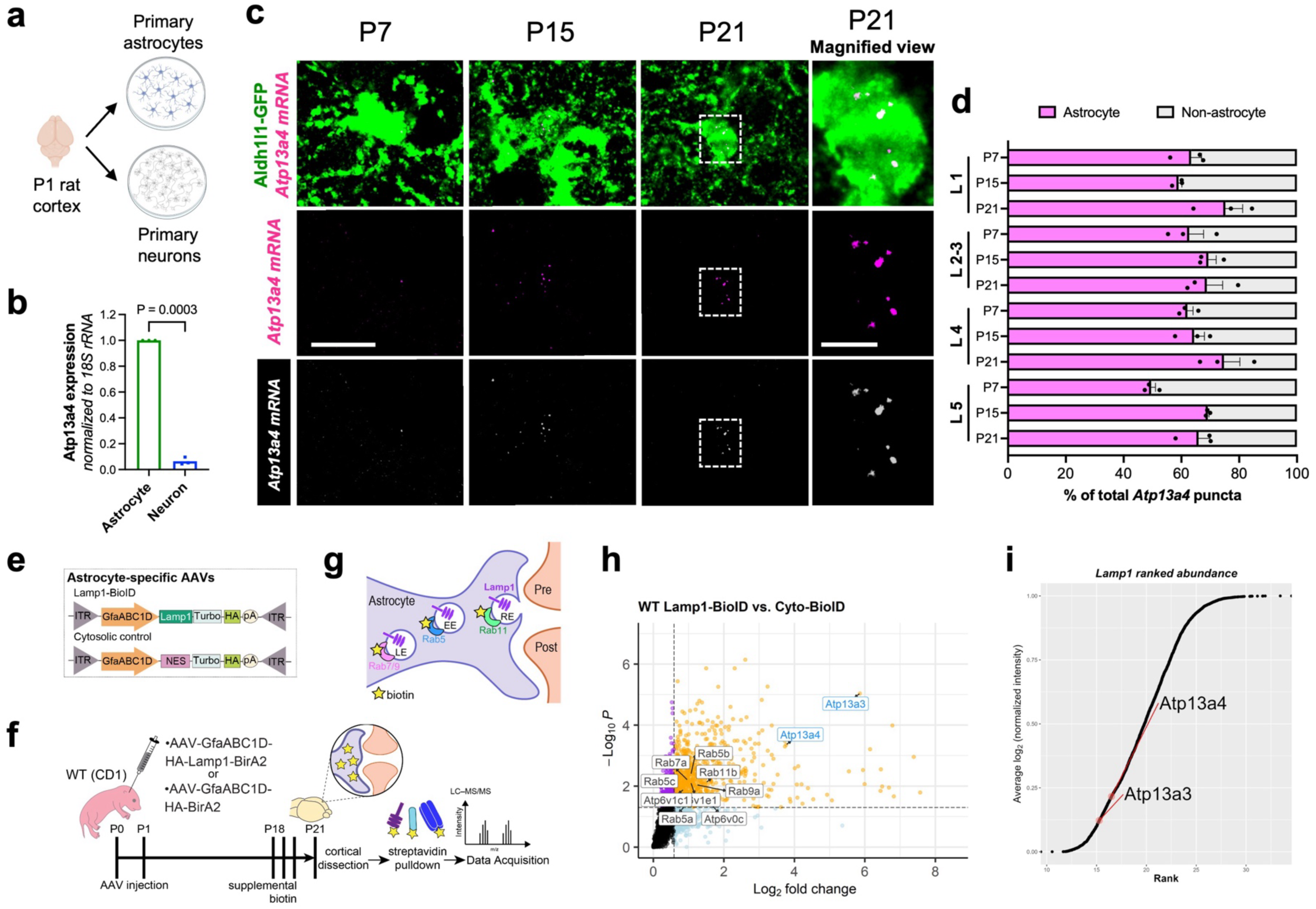
**ATP13A4 is expressed in astrocytes**. **a**, Schematic representation of isolating primary astrocyte and primary neuron cultures from postnatal day 1 (P1) rat cortex. **b**, qPCR analysis showing significantly higher expression of *rAtp13a4* in rat primary cortical astrocytes compared to neurons. *n* = 3 independent cultures. Paired *t* test. **c**, Detection of *msAtp13a4* transcripts in cortical astrocytes by RNA FISH in Aldh1L1-eGFP mice at P7, P15, and P21. Scale bars, 10 μm; magnified view, 3 μm. The top row shows merged images of Aldh1L1-eGFP-positive astrocytes (green) and *msAtp13a4* RNA FISH puncta (magenta). The middle row shows the *msAtp13a4* RNA FISH channel alone, and the bottom row shows white-on-black renderings of the *msAtp13a4* channel highlighting individual RNA puncta to facilitate visualization. The rightmost column shows a magnified view of the region indicated by the dashed box in the P21 panels. **d**, Quantification of astrocyte-specific versus non-astrocyte *mAtp13a4* puncta across all cortical layers (L1, L2/3, L4, and L5) of the V1 cortex at P7, P15, and P21. Each bar represents 100% of total *msAtp13a4* mRNA puncta, with the astrocyte fraction (magenta) defined as puncta overlapping with Aldh1L1-eGFP-positive astrocyte soma volume. Data shown as mean ± SEM. 3 sections per animal, 3 animals per time point. **e**, Astrocyte-specific AAVs (serotype PHP.eB) were used to express Lamp1 or nonspecific cytosolic control. NES, nuclear export sequence; ITR, inverted terminal repeats; GfaABC1D truncated GFAP promoter; HA, hemagglutinin tag; Turbo, TurboID biotin ligase; pA, polyadenylation. **f**, Outline of the experimental paradigm. *n* = 3 biological replicates per construct per genotype (1 replicate = 2 animals per pooled sample). **g**, Schematic of Lamp1 protein localization within astrocyte perisynaptic process (PAP). **h**, Volcano plot showing the differential abundance of proteins detected by Astro-Lamp1-TurboID compared to Astro-Cyto-TurboID. Dotted lines indicate threshold values FC > 1.5 (log2FC > 0.58), p-value < 0.05 (log10p <-1.3). **i**, Empirical cumulative distribution function (eCDF) listing potential endo-lysosomal polyamine transporters (red dots) among the most abundant proteins detected by Astro-Lamp1-TurboID.

Next, we investigated the subcellular localization of ATP13A4. Since no validated antibodies are available for immunohistochemistry or immunocytochemistry of ATP13A4, we used an overexpression construct to visualize its subcellular distribution. In a HeLa overexpression model, mCherry-labeled msAtp13a4 predominantly localized to late endo-/lysosomal compartments (**Supplementary Fig. 3**), consistent with our previous observations and with the reported endo-/lysosomal localization of related P5B-type transporters ATP13A2 and ATP13A3 (70, 92, 93). To confirm these findings and assess endogenous msAtp13a4 localization *in vivo*, we performed astrocyte-specific *in vivo* proximity biotinylation (iBioID) using a Lamp1-TurboID fusion protein (94) (**Supplementary Fig. 4, Fig. 1e-i**). This approach leverages astrocyte-specific AAVs, which were injected into P1 mice, to selectively biotinylate proteins within the endo-/lysosomal compartment of astrocytes, while preserving interactions with neighboring cells, thereby providing a physiologically relevant view of msAtp13a4 expression and localization. Unlike conventional immunofluorescence, TurboID labeling captures proteins in close proximity over time, allowing detection of dynamic or transient associations. Quantitative mass spectrometry revealed enrichment of msAtp13a4, confirming its presence in LAMP1-positive compartments in astrocytes (**Fig. 1h**). msAtp13a3 was also detected, albeit at lower levels (**Fig. 1i**), supporting the view that msAtp13a4 is a major P5B-type transport ATPase in the astrocytic endo-/lysosomal system.

### Biochemical validation of ATP13A4 polyamine transport activity

The precise molecular function of ATP13A4 remains unclear. Although ATP13A4 was implicated in polyamine uptake in breast cancer cells (71), conclusive evidence regarding its polyamine transport function and substrate selectivity is lacking. Therefore, we conducted biochemical studies on purified human ATP13A4 (hATP13A4) to determine if ATP13A4 is a *bona fide* polyamine transporter. ATP13A4 is a member of the P-type ATPase family, which use ATP hydrolysis to form an auto-phosphorylated intermediate driving the selective transport of ions, polyamines, or lipids. These transporters cycle between four main conformations (E1, E1P, E2P and E2), providing alternating access to the substrate binding site from the cytosolic (in the E1 state) to the extracytosolic side (in the E2P conformation) (**Fig. 2a**). ATP13A2 is the prototypical P5B-type ATPase, which we previously biochemically characterized as a polyamine transporter (5), a finding later validated at the structural level (74–78). ATP13A4 shares 37.1% sequence identity and 52.7% sequence similarity with ATP13A2. Despite this moderate overall similarity, AlphaFold predicts a conserved protein structure with high conservation of residues within the luminal substrate-binding cavity (**Supplementary Fig. 5a**), suggesting a conserved polyamine transport function and mechanism.

**Fig. 2:**
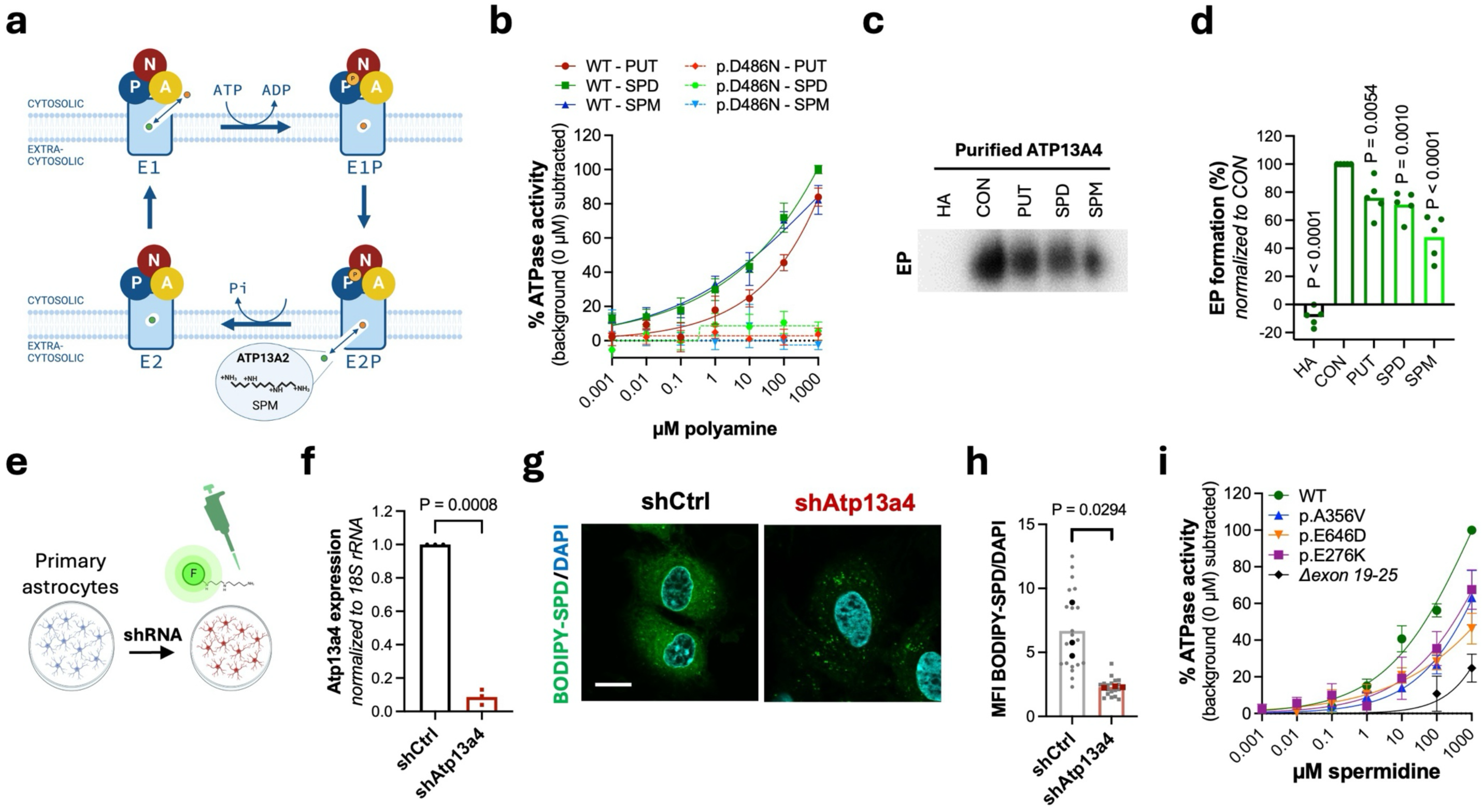
ATP13A4 is a broad-specificity polyamine transporter. **a**, Schematic of the catalytic cycle of P-type ATPases. The enzyme alternates between cytosolic-facing (E1/E1P) and extracytosolic-facing (E2P/E2) conformations driven by ATP-dependent phosphorylation and dephosphorylation. The “extracytosolic” side refers to polyamines located outside the cytosol, including either the extracellular space or the lumen of endo-/lysosomal compartments. Orange and green circles denote alternating ligands bound and released during the transport cycle; in the case of the archetypical P5B ATPase ATP13A2, the orange circle represents the polyamine substrate translocated from the extracytosolic to the cytosolic side, while no counter-transported ligand has been identified. Created in BioRender. van Veen, S. (2026) https://BioRender.com/choqqfq. **b**, Dose-response curves showing the effect of putrescine (PUT), spermidine (SPD) and spermine (SPM) on the ATPase activity of solubilized purified ATP13A4 (WT or catalytically dead p.D486N mutant). The number of independent biological experiments was as follows: *n* = 3 (D486N PUT, SPD, SPM), *n* = 5 (WT PUT), *n* =7 (WT SPD) and *n* = 4 (WT SPM). **c-d**, ATP13A4 phosphoenzyme (EP) levels in control (CON) conditions and in the presence of PUT, SPD, SPM, or hydroxylamine (HA). **c**, Representative autoradiogram. **d**, Quantification of EP. *n* = 5 independent experiments. One-way ANOVA with Dunnett’s multiple comparisons test. **e**, Schematic illustrating the BODIPY-polyamine uptake assay in primary astrocytes with ATP13A4 knockdown. Created in BioRender. van Veen, S. (2026) https://BioRender.com/94lyr9v. **f**, shRNA targeting mouse/rat *Atp13a4* efficiently reduces Atp*13a4* expression in astrocytes compared to control in which the shRNA sequence was scrambled (shCtrl). *n* = 3 independent cultures. Paired *t* test. **g-h**, Confocal microscopy of BODIPY-labelled SPD distribution. Scale bar, 10 μm. Mean fluorescence intensities (MFI) of BODIPY-SPD (shCtrl, 20 cells; shAtp13a4, 18 cells). *n* = 3 independent cultures. Paired *t* test. **i**, Dose-response curve showing spermidine-dependent ATPase activity of purified, solubilized ATP13A4 (WT and disease-associated variants p.E276K, p.A356V, p.E646D and *Δexon 19-25*). *n* = 4 independent experiments.

To biochemically characterize ATP13A4, we overexpressed hATP13A4 WT or the catalytically dead p.D486N mutant (which harbors a mutation in the catalytic site required for autophosphorylation) in HEK293T cells or *Saccharomyces cerevisiae* for subsequent purification via affinity chromatography (**Supplementary Fig. 5b-g**). As substrate binding triggers ATP hydrolysis in P-type ATPases, we assessed substrate specificity of hATP13A4 by testing a panel of polyamine-related compounds for their ability to stimulate ATPase activity. Putrescine, spermidine, spermine, cadaverine, and acetylated polyamine derivatives each stimulated ATPase activity of purified wild-type (WT) hATP13A4 in a concentration-dependent manner, whereas the polyamine precursor ornithine did not (**Fig. 2b, Supplementary Fig. 5h**). No stimulation was observed for the transport-deficient mutant p.D486N (**Fig. 2b**). ATP13A4 exhibits the highest apparent affinity to spermine and spermidine although a broader substrate specificity is observed as compared to ATP13A2 (5).

To analyze the auto-phosphorylation activity of hATP13A4, we incubated purified hATP13A4 with radiolabeled [γ-^32^P] ATP and observed that, in the absence of polyamines, hATP13A4 spontaneously formed a phosphoenzyme, similar to ATP13A2 (95). This phosphoenzyme was sensitive to hydroxylamine, a hallmark of P-type ATPase auto-phosphorylation on a conserved aspartate residue (**Supplementary Fig. 5i**) and showed minimal response to ADP (**Supplementary Fig. 5j-k**). The latter finding suggests that phosphorylated hATP13A4 mainly exists in the E2P state, since only the E1P state can interact with ADP to regenerate ATP (**Fig. 2a**). We also observed a slow dephosphorylation rate when non-radiolabeled ATP was added in the absence of polyamines suggesting low phosphoenzyme turnover (**Supplementary Fig. 5j-m**). However, the presence of spermidine or spermine significantly accelerated this dephosphorylation (**Supplementary Fig. 5l-m**), consistent with the polyamine-induced ATPase activity (**Fig. 2b**). Corroborating this, putrescine, spermidine, and spermine reduced the steady-state levels of the hATP13A4 phosphoenzyme (**Fig. 2c-d, Supplementary Fig. 5n-o**). Thus, polyamines are not required for auto-phosphorylation of hATP13A4, which occurs in the E1 (cytosolic-facing), but instead promote dephosphorylation of the E2P state, in which the substrate binding site is exposed to the extracytosolic side (**Fig. 2a**). These findings indicate that hATP13A4 transports polyamines from the extracytosolic side of the membrane into the cytosol, consistent with the highly conserved luminal polyamine binding pocket and structural conservation with hATP13A2, pointing to a conserved transport direction from the exoplasmic to cytoplasmic side (5) (**Fig. 2a**).

Taken together, our results establish ATP13A4 as a broad-specificity polyamine transporter with highest apparent affinity for spermine and spermidine.

### ATP13A4 mediates polyamine uptake in astrocytes

Given its high expression in astrocytes, we next assessed whether ATP13A4 mediates polyamine uptake into these glial cells as a candidate member of the so-far unidentified astrocytic polyamine uptake system (96, 97). To this end, we used two glial cell lines with modulated ATP13A4 expression and measured uptake of fluorescently labeled BODIPY-polyamines, which are genuine transported substrates of P5B-type ATPases ATP13A2 and ATP13A3 (69).

We first used stable neuroglioma H4 cell lines with lentiviral overexpression of hATP13A4 WT or p.D486N (**Supplementary Fig. 6a-b**) and performed flow cytometry-based BODIPY-polyamine uptake measurements. Compared to control cells expressing the p.D486N mutant or firefly luciferase (FLUC), hATP13A4 WT protein enhanced uptake of BODIPY-labelled putrescine, spermidine, and spermine (**Supplementary Fig. 6c-e**), in line with the broad polyamine specificity of ATP13A4. Pharmacological inhibition of endocytosis or incubation at 4 °C markedly reduced BODIPY-spermidine uptake in control and catalytically inactive cells, whereas uptake in hATP13A4 WT-expressing cells was only partially affected (**Supplementary Fig. 6f-g**). These findings support a transporter-mediated contribution to cellular polyamine accumulation that at least in part is endosomal mediated.

To assess the role of ATP13A4 under endogenous conditions, we next performed loss-of-function experiments in C8-D1A cells, a mouse cerebellar astrocyte cell line with endogenous *msAtp13a4* expression. Lentiviral microRNA-based knockdown reduced *msAtp13a4* mRNA levels by 80% (**Supplementary Fig. 6h**) and significantly decreased BODIPY-polyamine uptake compared to control cells transduced with a microRNA targeting the *FLUC* gene (mirFLUC), as evidenced by flow cytometry (**Supplementary Fig. 6i-k**) and confocal microscopy analysis (**Supplementary Fig. 6l-o**). Notably, in *msAtp13a4* knockdown cells, BODIPY-polyamines accumulate in intracellular puncta consistent with endosomal/vesicular compartments (**Supplementary Fig. 6l, n**), suggesting that ATP13A4 facilitates cytosolic access of polyamines and prevents their sequestration within these compartments. To rule out any influence of serum polyamine oxidase activity, we repeated the uptake experiments in a serum-free medium and observed a similar phenotype (**Supplementary Fig. 6p-q**). Consistent with reduced uptake, mass spectrometry analysis demonstrated significantly lower endogenous spermidine levels and a trend towards lower putrescine levels in *msAtp13a4* deficient C8-D1A cells compared to control cells (**Supplementary Fig. 6r**), supporting a key role for ATP13A4 in cellular polyamine uptake and homeostasis.

Finally, we confirmed the polyamine uptake role of Atp13a4 in primary rat astrocytes from P1 rat cortices (**Fig. 2e**). We silenced *rAtp13a*4 expression by more than 90% using short hairpin RNA (shRNA, shAtp13a4) compared to a non-targeting scrambled shRNA (shCtrl) (**Fig. 2f**). Using confocal microscopy, we found that shAtp13a4 astrocytes displayed significantly reduced uptake of BODIPY-labeled spermidine (**Fig. 2g-h**). While the uptake of BODIPY-labeled spermine showed a trend toward lower levels in knockdown cells, this difference was not statistically significant (**Supplementary Fig. 6s-t**). These findings suggest that rAtp13a4 plays a key role in mediating spermidine uptake in primary astrocytes, with a potential but less pronounced contribution to spermine uptake.

Altogether, these data establish ATP13A4 as a key component of the astrocytic polyamine uptake system that enables uptake of extracellular polyamines.

### Human ATP13A4 loss-of-function variants are linked to neurodevelopmental disorders

Several rare ATP13A4 variants have been reported in individuals with neurodevelopmental phenotypes, prompting us to examine whether they impair transporter function. ATP13A4 has been implicated in neurodevelopmental disorders through copy number variations, including the 3q29 deletion associated with schizophrenia (98) and duplications at 3q28-q29 linked to epileptic encephalopathies (99). In addition, a predicted loss-of-function variant (p.Q273X) was identified in an infant with epilepsy (100), and a chromosomal inversion disrupting the *ATP13A4* gene has been reported in individuals with specific language impairment (101). Missense variants (p.E646D and p.A356V) have also been associated with speech and autism-related phenotypes (101–103). Based on data from Genome Aggregation Database (gnomAD), p.A356V is rare (allele frequency 7.99 × 10^-5^), whereas p.E646D is relatively common (allele frequency ∼0.10). Notably, both variants affect residues that are highly conserved across diverse species (**Supplementary Fig. 7a-b**). To assess the impact of patient-derived missense substitutions on ATP13A4 function, we focused on the recurrent p.E646D and p.A356V variants. Both variants exhibited loss-of-function, marked by reduced phospho-enzyme levels, polyamine-induced ATPase and/or dephosphorylation activities (**Fig. 2i; Supplementary Fig. 7c-n**). Consistent with these biochemical defects, H4 cells expressing these variants displayed significantly impaired uptake of BODIPY-labeled polyamines (**Supplementary Fig. 6c-e, p**), indicating compromised transporter function.

Through *GeneMatcher* (104), we identified additional candidate pathogenic ATP13A4 variants. First, a novel homozygous p.E276K variant (**Supplementary Fig. 7a-b**), with an allele frequency of 4.337 × 10^-6^ (gnomAD), was identified in an individual with intellectual disability. Second, a heterozygous deletion spanning exons 19-25 of ATP13A4, predicted to disrupt catalytically important transmembrane and auto-phosphorylation domains (**Supplementary Fig. 7b**), was identified in a patient with generalized epilepsy. Although no alternative genetic causes or candidate variants were identified in the patient, this variant was inherited from an apparently unaffected mother, indicating incomplete penetrance or compensation at later life (prior history is unknown). Functional analysis showed that p.E276K reduces spermidine-stimulated ATPase activity to a similar extent as p.A356V and p.E646D, whereas the exon 19-25 deletion causes a more pronounced reduction (**Fig. 2i)**.

Together, these findings demonstrate that ATP13A4 variants impair transporter function across a spectrum of genetic contexts, supporting a loss-of-function mechanism with variable severity and penetrance in neurodevelopmental disorders. Consistent with this model, analysis of human RNA-seq data revealed that ATP13A4 expression is significantly reduced in astrocytes from patients with temporal lobe epilepsy (TLE) compared to temporal lobe (TL) controls (**Supplementary Fig. 7o**) (105), further linking altered ATP13A4 expression to disease-relevant contexts.

### ATP13A4 regulates astrocyte morphogenesis

Having established that ATP13A4 is enriched in astrocytes and that patient-derived variants impair its function, we next asked whether reduced ATP13A4 activity affects astrocyte morphology during the early postnatal period when astrocytes undergo morphological maturation.

To model partial loss of ATP13A4 function, we used an shRNA-mediated knockdown approach in a well-established cortical astrocyte-neuron co-culture assay (59, 106, 107) (**Fig. 3a**), which enables assessment of neuronal contact-dependent astrocyte morphogenesis of GFP-expressing astrocytes. This assay takes advantage of the fact that astrocytes gain a complex arborization morphology *in vitro*, visualized by GFP expression, only when cultured on top of neurons, mimicking what happens *in vivo*. For these assays, we independently isolated astrocytes and neurons from P1 rat cortices and specifically silenced *rAtp13a4* expression in astrocytes (**Fig. 2f**). After 48 h of co-culture with WT neurons, the morphological complexity of GFP-labeled control (shCtrl) or Atp13a4 silenced (shAtp13a4) astrocytes was assessed by Sholl analysis (108), which measures the number of intersections at defined concentric radii, with a higher count indicating a more complex arborization pattern. When co-cultured with neurons, astrocytes transfected with shCtrl displayed the typical complex arborization morphology (**Fig. 3b-c**). In contrast, shAtp13a4 astrocytes displayed a pronounced reduction in morphological complexity (**Fig. 3b-c**). To determine whether this phenotype reflects a general consequence of ATP13A4 loss rather than an artefact of the knockdown approach, we next analyzed primary astrocytes derived from a CRISPR-Cas9-generated *Atp13a4* knockout (KO) mouse line. This mouse model carries a deletion of exons 3-5 resulting in a premature stop codon at position 86 (p.A85*) and was validated as a catalytically inactive loss-of-function allele (**Supplementary Fig. 8**). Using a similar astrocyte-neuron co-culture paradigm, in which *Atp13a4* KO or WT astrocytes were plated onto WT neurons, KO astrocytes exhibited a significant reduction in arborization and morphological complexity compared to WT controls (**Fig. 3d-e**), consistent with the phenotype observed in the shRNA-mediated knockdown experiments. These findings demonstrate that loss of Atp13a4 function is sufficient to impair astrocyte morphogenesis.

**Fig. 3:**
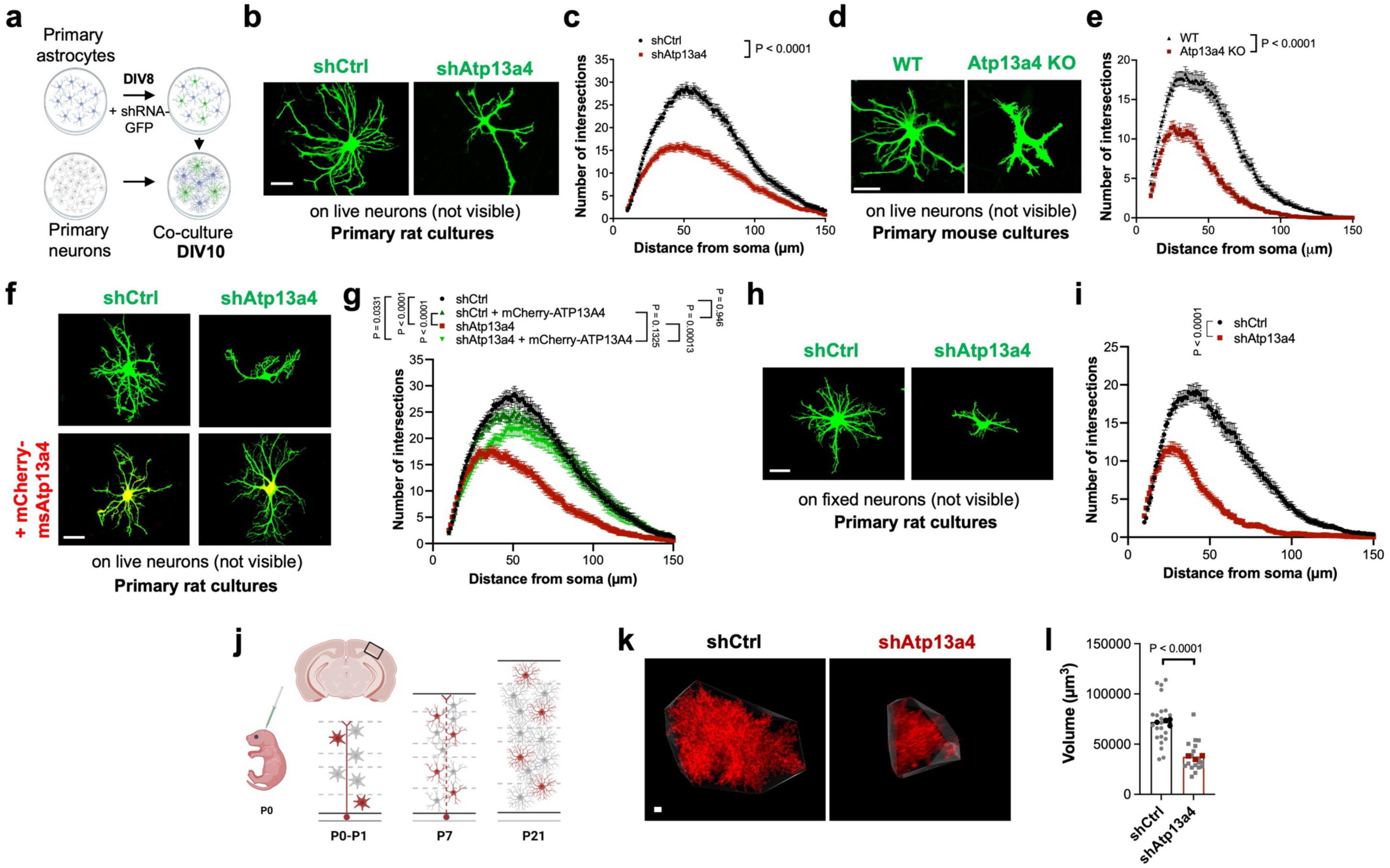
ATP13A4 knockdown impairs astrocyte arborization. **a**, Schematic of astrocyte-neuron co-culture used to assess astrocyte morphology. Created in BioRender. van Veen, S. (2026) https://BioRender.com/7x2wn63. **b**, Representative images of astrocytes (green) transfected with either shRNA targeting mouse/rat *Atp13a4* (shAtp13a4) or a scrambled control (shCtrl), co-cultured with WT neurons (not visible) for 48 h. Scale bar, 50 μm. **c**, Quantification of astrocyte complexity via Sholl analysis for conditions in **b**. *n* = 72 astrocytes per condition from three independent experiments. **d**, Representative images of GFP-labeled astrocytes (green) from WT and *Atp13a4* KO mice following 48 h co-culture with WT neurons. Scale bar, 50 μm. **e**, Quantification of astrocyte complexity by Sholl analysis for conditions shown in **d**. *n* = 103 (WT) and 61 (KO) astrocytes per condition from four (WT) or three (KO) independent experiments. **f**, Representative images of shRNA expressing astrocytes (green) in the presence or absence of an shRNA-resistant mouse Atp13a4 (msAtp13a4) construct with an N-terminal mCherry tag (red) after 48 h co-culture with WT neurons. Scale bar, 50 μm. **g**, Quantification of astrocyte complexity for conditions in **f**. *n* = 60 astrocytes per condition from three independent experiments. **h**, Representative images of shRNA expressing astrocytes co-cultured with methanol-fixed WT neurons for 48 h. Scale bar, 50 μm. **i**, Quantification of astrocyte complexity for conditions in **h**. *n* = 60 astrocytes per condition from three independent experiments. **c**, **e**, **g**, **i**, Mixed effects ANOVA (**c**, **e**, **g**) with Tukey’s post-hoc test (**g**). **j**, Schematic of PALE (Postnatal Astrocyte Labeling by Electroporation). Plasmids were injected into the lateral ventricle of CD1 mice at late P0, followed by electroporation into radial glial stem cells, leading to sparse knockdown and labeling of cortical astrocytes. Created in BioRender. van Veen, S. (2026) https://BioRender.com/fhbbf7l. **k**, Representative images of *in vivo* shRNA-transfected astrocytes (labelled with mCherry) at P21. **l**, Quantification of *in vivo* astrocyte territory volumes by convex hull analysis in Imaris. Only astrocytes from V1 cortex were imaged and analyzed. The average astrocyte territory volume of individual mice is plotted in black (shCtrl) or red (shAtp13a4). *n* = 18–22 astrocytes from 3-4 mice, nested *t* test. All data are presented as mean ± SEM, except in **l** where individual data points are shown.

To further validate the specificity of *Atp13a4* loss, we performed rescue experiments using an shRNA-resistant msAtp13a4 construct tagged with mCherry (mCherry-msAtp13a4; **Supplementary Fig. 9a**). Co-expression of mCherry-msAtp13a4 with shAtp13a4 resulted in a partial rescue of astrocyte morphogenesis, as demonstrated by a significant increase in overall astrocyte complexity (**Fig. 3f-g**). Sholl analysis showed that astrocytes co-expressing shAtp13a4 and mCherry-msAtp13a4 were not significantly different from astrocytes expressing shCtrl and mCherry-msAtp13a4, but they remained statistically distinct from shCtrl astrocytes (**Fig. 3g**). Overexpression of mCherry-msAtp13a4 in shCtrl astrocytes did not further enhance complexity (**Fig. 3g**). These findings suggest that the morphological defects in shAtp13a4 astrocytes are largely attributable to loss of rAtp13a4, although the incomplete rescue suggests dose-dependent or regulatory effects.

Next, we wondered whether Atp13a4 controls astrocyte morphology through a mechanism involving dynamic neuron signaling. To test this, we fixed neurons with methanol before astrocyte co-culture, preserving neuronal structure and adhesion signals while eliminating synaptic activity and secretion. We found that loss of astrocytic *Atp13a4* still resulted in diminished astrocyte morphology when cultured on methanol-fixed neurons (**Fig. 3h-i**), indicating that the phenotype depends on neuronal contact but not necessarily on synaptic signaling or secreted neuronal cues.

To determine whether spermidine or spermine induce effects similar to Atp13a4 loss, we treated astrocyte-neuron co-cultures with these polyamines and assessed astrocyte morphology. We found that 8 h treatment with spermidine or spermine led to a dose-dependent reduction in the arborization of shCtrl astrocytes, with increasing concentrations progressively impairing astrocyte complexity (**Supplementary Fig. 9b-e**). In contrast, shAtp13a4 astrocytes showed no further reduction in arborization upon polyamine treatment, suggesting that the impact of ATP13A4 loss and exogenous polyamines converge on the same pathway regulating astrocyte arborization. Notably, a similar reduction in complexity was observed when shCtrl astrocytes were pre-treated with polyamines for 24 h before co-culture (**Supplementary Fig. 9f-i**). This pre-treatment experiment confirms that the observed effects are intrinsic to astrocytes, as polyamine exposure occurred prior to neuronal interaction.

Finally, we evaluated whether Atp13a4 function is required for cortical astrocyte morphogenesis *in vivo* and turned to postnatal astrocyte labeling by electroporation (PALE) (59, 106, 107) (**Fig. 3j**). For this approach, the previously described shRNAs targeting mouse/rat *Atp13a4* (shAtp13a4) and a scrambled control (shCtrl; **Fig. 2f**) were cloned into a PiggyBac transposon system expressing mCherry-CAAX. The shRNA plasmids were then electroporated into radial glial cells in the subventricular zone at P0, leading to selective knockdown of *msAtp13a4* in a subset of cortical astrocytes. Analysis of the knockdown astrocytes revealed a striking reduction in overall territory volume compared to control astrocytes (**Fig. 3k-l**). Taken together, these findings demonstrate an important role for ATP13A4-mediated polyamine homeostasis in astrocyte morphogenesis during postnatal development.

### ATP13A4 loss enhances astrocyte-driven excitatory synaptogenesis *in vitro*

While neuronal cues guide astrocyte morphogenesis, astrocytes promote the assembly and maturation of excitatory and inhibitory synaptic circuits (42–51). We therefore asked whether astrocytic polyamine transport regulates synaptogenesis by controlling extracellular polyamine levels. To address this question, we used purified cortical neuron cultures that were treated with an astrocyte-conditioned media (ACM). Many previous studies have shown that ACM strongly promotes synapse formation between neurons*, in vitro* (44, 50, 51, 109–111). We treated primary WT neurons with either i) ACM derived from astrocytes transfected with control shRNA (shCtrl ACM), ii) ACM from astrocytes with *rAtp13a4* knockdown (shAtp13a4 ACM), or iii) non-conditioned medium (no ACM).

We first assessed the role of astrocytic rAtp13a4 in neuronal dendrite morphogenesis by quantifying neuronal morphology after incubation with the different media. Consistent with previous literature (44), treatment with ACM significantly increased the length of dendritic processes compared to non-conditioned medium (**Supplementary Fig. 10a-b**). However, there were no significant differences in dendritic length between neurons treated with shCtrl ACM *versus* shAtp13a4 ACM (**Supplementary Fig. 10a-b**), suggesting that ATP13A4 is not essential for promoting dendritic growth through astrocyte-secreted factors.

Next, we investigated the effect of ACM on synaptogenesis by labeling excitatory synapses with the presynaptic marker Bassoon and the postsynaptic marker Homer1 (**Fig. 4a**). Leveraging the confocal resolution limit, we quantified the colocalization of pre-and postsynaptic terminals to assess synapse formation. As expected, neurons treated with ACM from shCtrl astrocytes exhibited a significant increase in excitatory synaptogenesis, as indicated by the close apposition of Bassoon and Homer1 puncta (**Fig. 4b-c**) (109, 110). Notably, ACM from *rAtp13a4* knockdown astrocytes further enhanced excitatory synapse formation (**Fig. 4b-c**), without altering the total number of individual Bassoon or Homer1 puncta (**Supplementary Fig. 10c-d**). Interestingly, this effect resulted in significantly more excitatory synaptic colocalizations compared to neurons treated with shCtrl ACM (**Fig. 4c**).

**Fig. 4:**
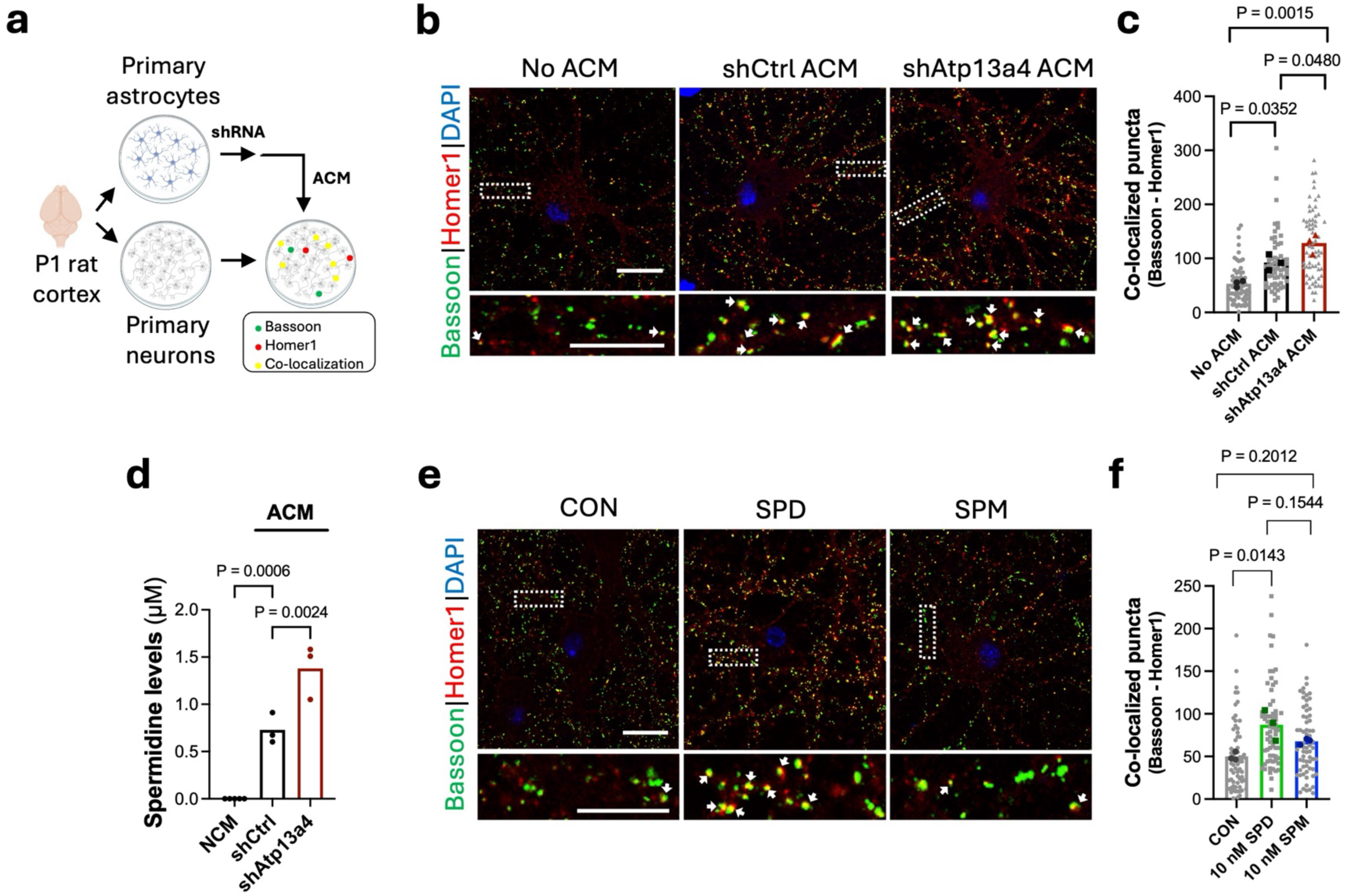
ATP13A4 controls excitatory synapse numbers and function. **a**, Schematic of neuronal culture assay to evaluate the effects of astrocyte-conditioned medium (ACM) on synapse formation. Created in BioRender. van Veen, S. (2026) https://BioRender.com/4lgtu1p. **b**, Excitatory synapses marked with Bassoon and Homer1. Scale bars, 20 μm. **c**, Quantification of excitatory synapses (Bassoon and Homer1 co-localization) in **b**. *n* = 58-64 cells per condition from three independent experiments. Individual data points are shown, with the average of each experiment plotted in grey (no ACM), black (shCtrl ACM) or red (shAtp13a4 ACM). Nested one-way ANOVA with Tukey’s post-test. **d**, Metabolomics analysis of spermidine levels in non-conditioned medium (NCM) and ACM from shCtrl and shAtp13a4 cells. *n* = 3 independent experiments. One-way ANOVA with Dunnett’s post-test. **e**, Excitatory synapses marked with Bassoon and Homer1 in neuronal cultures treated with 10 nM spermidine (SPD) or spermine (SPM). Scale bars, 20 μm. **f**, Quantification of excitatory synapses (Bassoon and Homer1 co-localization) in **e**. *n* = 64-66 cells per condition from three independent experiments. **c**, **f**, Individual data points are shown, with the average of each experiment plotted in grey (no ACM), black (shCtrl ACM), red (shAtp13a4 ACM), green (SPD) or blue (SPM). Nested one-way ANOVA with Tukey’s multiple comparisons test.

Given that Atp13a4 is important for polyamine uptake by astrocytes (**Fig. 2g-h**), we next investigated whether astrocytic Atp13a4 knockdown alters extracellular polyamine levels. Mass spectrometry analysis of ACM revealed a selective increase in endogenous spermidine in ACM from *rAtp13a4* knockdown astrocytes compared to control ACM (**Fig. 4d**), whereas levels of ornithine, putrescine, spermine and acetylated derivatives were unchanged (**Supplementary Fig. 10e-j**). Spermidine was not detectable in non-conditioned medium (**Fig. 4d**), indicating that astrocytes are the primary source of extracellular spermidine under these conditions. We then tested whether spermidine is sufficient to drive synaptogenesis using defined concentrations, selecting 10 nM (112) as the lowest concentration eliciting a significant effect on astrocyte morphology (**Supplementary Fig. 9b-e**). Treatment of purified neurons with spermidine significantly increased excitatory synapse numbers, phenocopying the effects of shAtp13a4 ACM, whereas spermine had a weaker effect (**Fig. 4e-f, Supplementary Fig. 10k-l**). Notably, spermidine did not alter dendritic length (**Supplementary Fig. 10m-n**), indicating that these effects are not driven by global changes in neuronal morphology.

Together, these results indicate that loss of Atp13a4 in astrocytes elevates extracellular spermidine, which promotes excitatory synapse formation.

### ATP13A4 loss disrupts brain polyamine homeostasis

To investigate the impact of ATP13A4 loss on brain polyamine homeostasis, we performed targeted LC-MS/MS metabolomic profiling of tissues from two-month-old *Atp13a4* WT and KO mice (validated in **Supplementary Fig. 8**). Plasma levels of ornithine, putrescine, spermidine, spermine, and their major acetylated derivatives did not differ between genotypes (**Supplementary Fig. 11a-g**), indicating that systemic polyamine levels are not affected by ATP13A4 loss. In contrast, analysis of cortical tissue revealed a clear, brain-specific disruption of polyamine homeostasis. Putrescine levels were significantly reduced in KO cortices, whereas ornithine, spermidine and spermine remained unchanged (**Fig. 5a-b, Supplementary Fig. 11h-i**). In addition, the acetylated derivatives N^1^-acetylspermine and N^1/8^-acetylspermidine were significantly decreased, while N-acetylputrescine was unaffected (**Fig. 5c-d, Supplementary Fig. 11j**). Strikingly, analysis of the extracellular compartment revealed an opposing phenotype. Cerebrospinal fluid (CSF) from *Atp13a4* KO mice showed significantly increased levels of putrescine and N^1/8^-acetylspermidine, whereas other polyamine species were unchanged (**Fig. 5e-g, Supplementary Fig. 11k-m**). Together, these findings demonstrate a compartmental redistribution of brain polyamines upon ATP13A4 loss, characterized by selective depletion within cortical tissue and accumulation in the extracellular compartment, supporting a model in which altered extracellular polyamine availability contributes to the enhanced excitatory synaptogenesis observed in KO mice.

**Fig. 5:**
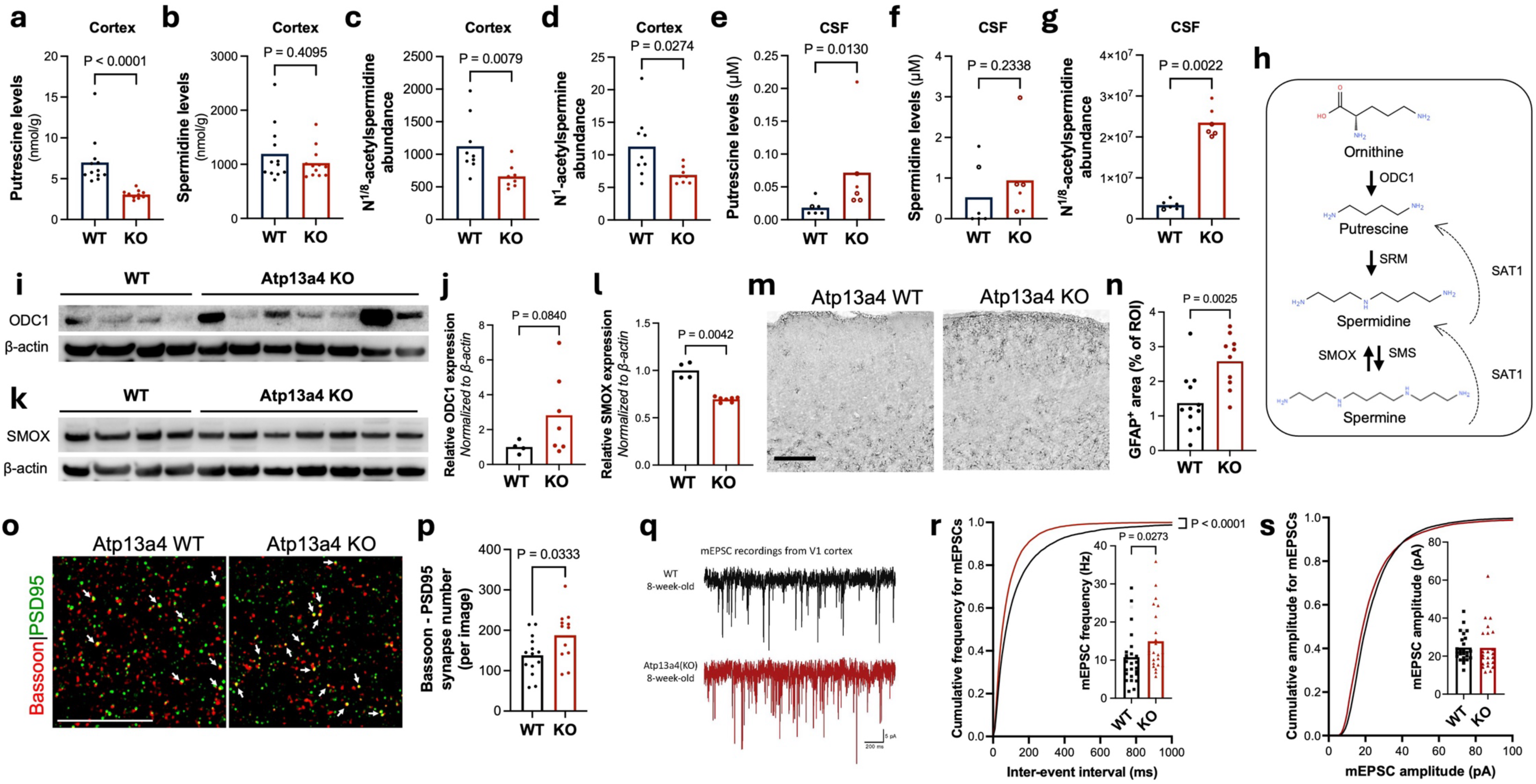
ATP13A4 loss disrupts brain polyamine homeostasis and enhances excitatory connectivity *in vivo*. **a**-**d**, Targeted LC-MS/MS quantification of putrescine (**a**), spermidine (**b**), N^1/8^-acetylspermidine (**c**) and N^1^-acetylspermine (**d**) in cortical tissue from two-month-old *Atp13a4* WT and KO mice. For **a** and **b**, *n* = 12 (WT) and 12 (KO) mice; for **c** and **d**, *n* = 9 (WT) and 8 (KO) mice. Data are presented as mean, with individual data points representing individual mice. Statistical comparisons were performed using a two-tailed Mann-Whitney U test. **e**-**g**, Targeted LC-MS/MS quantification of putrescine (**e**), spermidine (**f**) and N^1/8^-acetylspermidine (**g**) in cerebrospinal fluid (CSF). Data are presented as mean, with individual data points representing biological samples. *n* = 6 biological samples per genotype. Filled symbols indicate single-animal CSF samples; open symbols indicate pooled CSF samples derived from 2-3 mice. Following blank subtraction, some spermidine values fell below the limit of detection (LOD) and are reported as <LOD; for visualization, these values are plotted as 0. Statistical comparisons were performed using two-tailed Mann-Whitney U tests. **h,** Schematic overview of polyamine biosynthetic and catabolic pathways, indicating enzymatic steps catalyzed by ODC1 (ornithine decarboxylase 1), SRM (spermidine synthase), SMS (spermine synthase), SMOX (spermine oxidase), and SAT1 (spermidine/spermine N^1^-acetyltransferase 1). **i**-**l**, Western blot analyses and corresponding quantifications of ODC1 (**i**-**j**) and SMOX (**k**-**l**) expression in primary astrocytes from *Atp13a4* WT and KO mice. β-actin served as a loading control. Data are presented as mean, with individual data points representing independent biological replicates (individual primary astrocyte preparations). Statistical comparisons were performed using unpaired two-tailed *t* tests. **m**, Gfap immunostaining in cortex of two-month-old *Atp13a4* WT and KO mice. Scale bar, 50 µm. **n**, Quantification of Gfap immunoreactivity. Data points represent mouse averages. *n* = 10 (KO) and 12 (WT) mice, with 2 sections per mouse and 5 ROIs per section. Statistical comparisons were performed using an unpaired two-tailed *t* test. **o**, Excitatory synapses (Bassoon and PSD95 co-localization) in V1 cortex of 8-week-old *Atp13a4* WT and KO mice. White arrows indicate excitatory synapses. Scale bar, 10 μm. **p**, Quantification of excitatory synapse numbers in *Atp13a4* WT and KO mice. *n* = 12 (KO), 14 (WT) mice with 2 sections/mouse and 3-4 images/section. Data points represent mouse averages. Unpaired two-tailed *t* test. **q**, Representative mEPSC (miniature excitatory postsynaptic currents) traces from V1 cortex in acute brain slices of two-month-old WT and *Atp13a4* KO mice. **r**, Quantification of frequency average (inset) and cumulative probability of mEPSC from *Atp13a4* WT and KO neurons. **s**, Quantification of amplitude average (inset) and cumulative probability of mEPSC from *Atp13a4* WT and KO neurons. **r**-**s,** *n* = 26 (WT), 23 (*Atp13a4* KO) neurons from 3 mice per genotype. Mann-Whitney U test (inset). Kolmogorov-Smirnov test (cumulative probability). Individual data points are shown.

To determine whether this imbalance triggers adaptive responses within the polyamine pathway (**Fig. 5h**), we next quantified the expression of key biosynthetic and catabolic enzymes in both brain tissue and astrocytes. ODC1 (ornithine decarboxylase 1), the rate-limiting enzyme for polyamine synthesis, was significantly upregulated in *Atp13a4* KO brain (**Supplementary Fig. 12a-b**) and showed a similar increasing trend in KO astrocytes (**Fig. 5i-j**). In addition, SRM (spermidine synthase) expression was significantly increased in KO astrocytes, whereas it remained unchanged in brain tissue (**Supplementary Fig. 12c-d, k-l**). In contrast, SMS (spermine synthase) expression was not altered in either condition (**Supplementary Fig. 12e-f, m-n**). Among catabolic enzymes, SMOX (spermine oxidase) expression was significantly reduced in KO brain and astrocytes (**Supplementary Fig. 12g-h, Fig. 5k-l**), whereas SAT1 (spermidine/spermine N^1^-acetyltransferase) expression was not significantly altered (**Supplementary Fig. 12i-j, o-p**). These changes indicate a coordinated shift upon ATP13A4 loss toward a polyamine-conserving metabolic state, characterized by increased biosynthetic capacity with reduced catabolism. Although astrocytes are not considered a major source of *de novo* polyamine synthesis under basal conditions (31–35), these findings suggest that impaired ATP13A4-dependent uptake triggers a compensatory, cell-intrinsic rewiring of polyamine metabolism to preserve intracellular polyamine availability.

Together, ATP13A4 emerges as a key determinant of polyamine compartmentalization in the brain, and its loss triggers extracellular accumulation alongside compensatory metabolic adaptation aimed at preserving intracellular polyamine availability.

### ATP13A4 loss enhances excitatory synaptogenesis *in vivo*

Having established that ATP13A4 loss disrupts polyamine compartmentalization in the brain, we next examined the impact of ATP13A4 deficiency on astrocyte state and synaptic connectivity *in vivo*. GFAP immunostaining revealed increased astrocyte reactivity in the cortex of *Atp13a4* KO mice compared to WT controls (**Fig. 5m-n**), indicative of altered astrocyte homeostasis, while microglial activation (Iba1) was not altered between genotypes (**Supplementary Fig. 12q-r**). We next quantified excitatory synapses in the V1 cortex of 8-week-old mice based on co-localization of presynaptic Bassoon and postsynaptic PSD95. We found a significant increase in excitatory synapse numbers (∼25%) in *Atp13a4* KO mice compared to WT littermates (**Fig. 5o-p**). To assess whether the altered synapse numbers in the V1 of *Atp13a4* KO mice affect synaptic function, we performed whole-cell patch-clamp recordings of miniature excitatory postsynaptic currents (mEPSCs) and miniature inhibitory postsynaptic currents (mIPSCs) in layer 2/3 pyramidal neurons of the V1 visual cortex from acute brain slices of 8-week-old *Atp13a4* KO and WT mice (**Fig. 5q-s, Supplementary Fig. 10o-q**). *Atp13a4* KO neurons exhibited a significant increase (∼25%) in mEPSC frequency and inherent leftward shift in the cumulative distribution of mEPSC inter-event intervals compared to WT neurons (**Fig. 5r**), while mean mEPSC amplitudes remained similar between genotypes (**Fig. 5s**). Conversely, we did not observe any change in either the amplitude or the frequency of mIPSCs (**Supplementary Fig. 10p-q**).

Collectively, these findings demonstrate that loss of Atp13a4 disrupts astrocytic polyamine handling, leading to extracellular polyamine imbalance and enhanced excitatory synaptogenesis *in vivo*, as observed *in vitro*.

### ATP13A4 deficiency delays early postnatal neurodevelopment

Given that ATP13A4 loss disrupts brain polyamine homeostasis and enhances excitatory synaptogenesis, we next asked whether these alterations impact behavioral development across lifespan. During early postnatal development, *Atp13a4* KO mice exhibited delays in neurodevelopmental progression. Body weight trajectories showed a modest but significant difference between genotypes (**Fig. 6a**), indicating altered growth dynamics. Consistent with disrupted developmental timing, *Atp13a4* KO pups also exhibited a significant delay in eye opening (**Fig. 6b**). Notably, unilateral eye development was observed in a subset of *Atp13a4* KO pups (3/31), but not in WT animals (0/26), representing a low-frequency developmental abnormality (Fisher’s exact test, P = 0.25). To systematically assess neurodevelopmental progression, we quantified a series of postnatal developmental milestones (**Fig. 6c-k**). *Atp13a4* KO pups displayed significant delays across multiple assays, with rooting reflex and forelimb grasp reaching statistical significance after Bonferroni correction (**Fig. 6f, h**). These effects were consistent in direction across the cohort and largely independent of sex (**Supplementary Fig. 13**), indicating a reproducible pattern of delayed early postnatal development in *Atp13a4* KO mice.

**Fig. 6:**
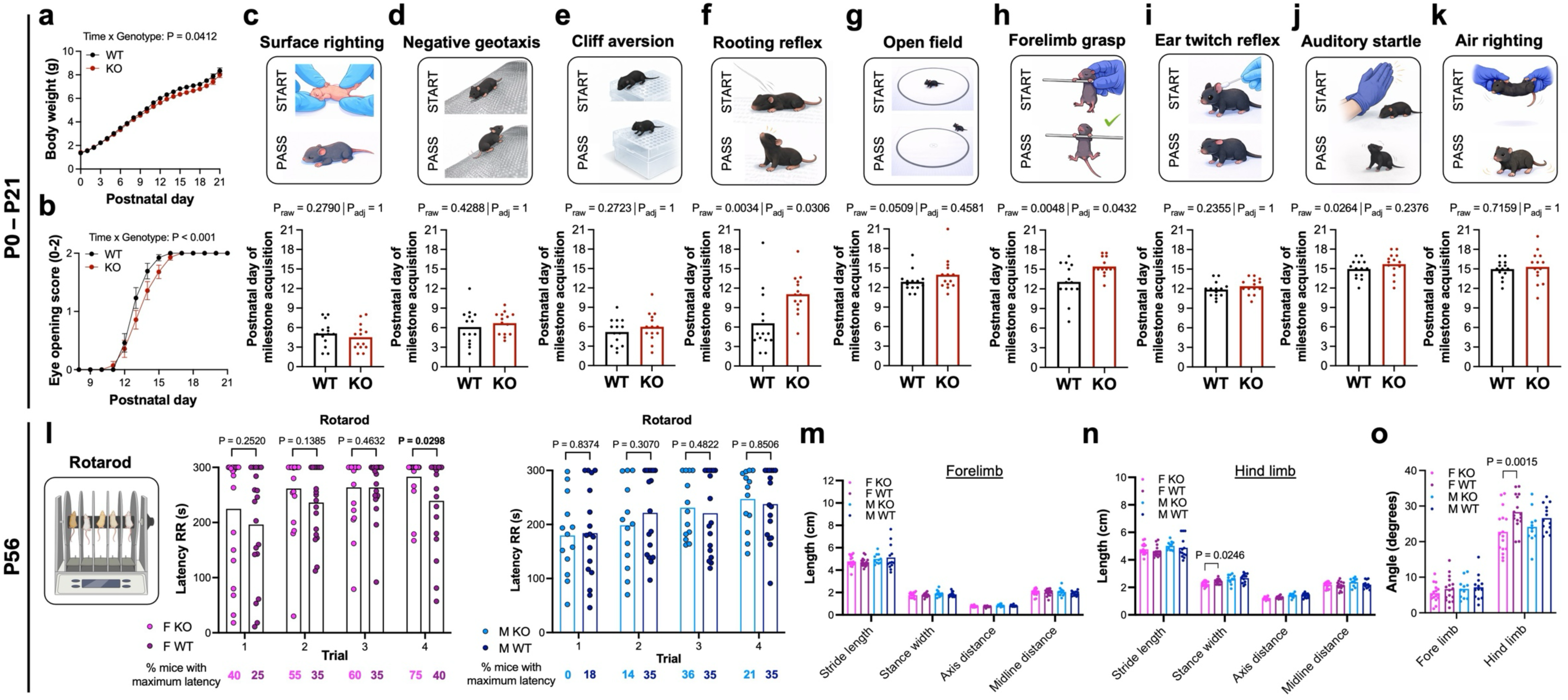
Atp13a4 deficiency causes early developmental delays and subtle, sex-specific adult behavioral alterations. **a**-**b**, Postnatal development of *Atp13a4* WT and KO pups. Body weight progression (**a**) and eye opening (**b**; 0 = no eyes open, 1 = one eye open, 2 = both eyes open) are shown as mean ± SEM. Statistical comparisons were performed using a linear mixed-effects model (**a**) and a cumulative link mixed model (**b**). For **a**, *n* = 26 (WT) and 31 (KO) mice; for **b**, *n* = 26 (WT) and 28 (KO) mice; animals with unilateral eye development were excluded from this analysis. **c-k**, Developmental milestone acquisition, including surface righting (**c**), negative geotaxis (**d**), cliff aversion (**e**), rooting (**f**), open field (**g**), forelimb grasp (**h**), ear twitch reflex (**i**), auditory startle (**j**), and air righting (**k**). Representative schematics are shown above the corresponding panels. Plotted values correspond to the second consecutive day on which pups met predefined acquisition criteria. Data are presented as mean, with individual data points representing matched litter averages (*n* = 14 litters). Statistical comparisons were performed using paired two-tailed *t* tests (**c**-**e**, **h**-**k**) or Wilcoxon matched-pairs signed-rank tests (**f**, **g**), as appropriate based on normality. P-values were adjusted for multiple comparisons across the nine milestone tests using Bonferroni correction (adjusted threshold α = 0.0056; or equivalently, raw p-values multiplied by 9 and compared to 0.05). **l**-**o**, Adult behavioral assessment in two-month-old mice, including rotarod performance (**l**) and DigiGait analysis (**m**-**o**). **l**, Schematic of the rotarod apparatus for motor coordination testing. Created in BioRender. van Veen, S. (2026) https://BioRender.com/6xrflic. Latency to fall from the rotating drum over four test trials in two-month-old female (left) and male (right) mice; percentages at the bottom indicate the proportion of mice that reached the maximum latency. **m-o**, Quantitative DigiGait analysis of locomotor parameters in *Atp13a4* KO and WT mice. Automated DigiGait recordings of ventral paw movement were used to extract temporal and spatial gait parameters describing stride geometry and timing. **m-n**, Spatial parameters for forelimbs (**m**) and hindlimbs (**n**). These include stride length (distance between successive placements of the same paw), stance width (distance between left and right paws), axis distance (lateral distance between the paw trajectory and the body’s central axis, indicating limb placement relative to the centerline), and midline distance (distance of each paw placement from the body midline, reflecting lateral symmetry). **o**, Paw angle (angular orientation of the paw relative to the body axis during stance) was measured to assess limb alignment. **l-o**, Data are presented as mean with individual data points shown. Statistical comparisons were performed using a Mann-Whitney U test (**l**) and two-way ANOVA with Šídák’s (**m**-**n**) or Tukey’s post hoc tests (**o**).

To determine whether these early delays are accompanied by lasting consequences, we assessed two-month-old *Atp13a4* KO mice across multiple behavioral domains. In contrast to the robust and largely sex-independent early phenotype, adult alterations were modest and largely confined to female mice. Home-cage locomotion and circadian activity were unaffected (**Supplementary Fig. 14a-d**). In the open field, female KO mice showed a trend toward reduced center time and significantly increased thigmotaxis, while male KO mice were indistinguishable from controls (**Supplementary Fig. 14e-j**). In the Morris water maze, female KO mice exhibited greater average distances to the hidden platform during acquisition, consistent with a mild learning delay, whereas performance in probe trials was comparable across all groups, indicating intact memory retention (**Supplementary Fig. 14k-q**). Social preference behavior was unaffected regardless of sex or genotype (**Supplementary Fig. 14r**). Motor phenotyping revealed modest, sex-specific differences. Female KO mice showed improved performance on the accelerating Rotarod, with longer latencies to fall and a higher proportion reaching the maximal cutoff (57.5% vs. 35.5%, P = 0.0067), whereas male KO mice were unaffected (**Fig. 6l**). DigiGait analysis revealed reduced hind limb stance width and paw angle in female KO mice (**Fig. 6m-o**), consistent with a more centrally aligned hind limb posture that may underlie the improved Rotarod performance.

In line with the epilepsy observed in human ATP13A4 carriers, *Atp13a4* loss in mice influences seizure susceptibility albeit in a sex-specific manner. Following a low dose of pentylenetetrazol (PTZ, 25 mg/kg), a GABAergic antagonist (113), full seizures were rare across groups (3/20 female KO mice, 1/19 female WT mice, 0 male mice), and overall activity levels were comparable between genotypes (**Supplementary Fig. 15a-d**). However, RM ANOVA revealed altered dynamics: female KO mice exhibited an earlier onset and faster progression into immobility/inactivity compared to WT females and male groups (**Supplementary Fig. 15a-d**), suggesting an accelerated transition into seizure-related behaviors. Furthermore, analysis of the longest immobility episode across 3 min time bins revealed a significant interaction between genotype and time in female, but not male, KO mice (**Supplementary Fig. 15e-f**), indicating that Atp13a4 loss alters the progression of seizure-related immobility over time.

Overall, ATP13A4 deficiency causes a robust delay in early neurodevelopment, whereas adult behavioral alterations are mild, consistently female-biased, and do not indicate global functional impairment. These findings support a model in which early disruptions in polyamine-dependent circuit maturation are largely compensated over time in a sex-dependent manner, resulting in subtle and context-dependent behavioral outcomes in adulthood. Notably, sex-stratified analyses of molecular and cellular readouts in adult mice show that cortical polyamine levels and astrocyte reactivity are similarly altered across sexes, whereas excitatory synapse density displays sex-dependent differences (**Supplementary Fig. 16**), suggesting that downstream circuit-level adaptations diverge despite shared underlying metabolic perturbations.

## Discussion

Polyamines are increasingly recognized as critical regulators of brain development and synaptic function (8–17), yet the mechanisms governing their cellular uptake and extracellular availability have remained poorly defined (96, 97). Here, we identify ATP13A4 as a glia-enriched polyamine transporter that controls polyamine compartmentalization in the brain and thereby regulates astrocyte-neuron communication. By integrating biochemical, cellular, and *in vivo* approaches, we show that ATP13A4 promotes polyamine uptake in astrocytes and constrains extracellular polyamine levels. Loss of ATP13A4 disrupts this balance, leading to extracellular accumulation of polyamines, impaired astrocyte morphogenesis, and enhanced excitatory synaptogenesis (**Fig. 7**). These findings establish ATP13A4 as a key regulator of extracellular polyamine signaling and reveal a previously unrecognized mechanism by which astrocytes shape synaptic circuit development.

**Fig. 7:**
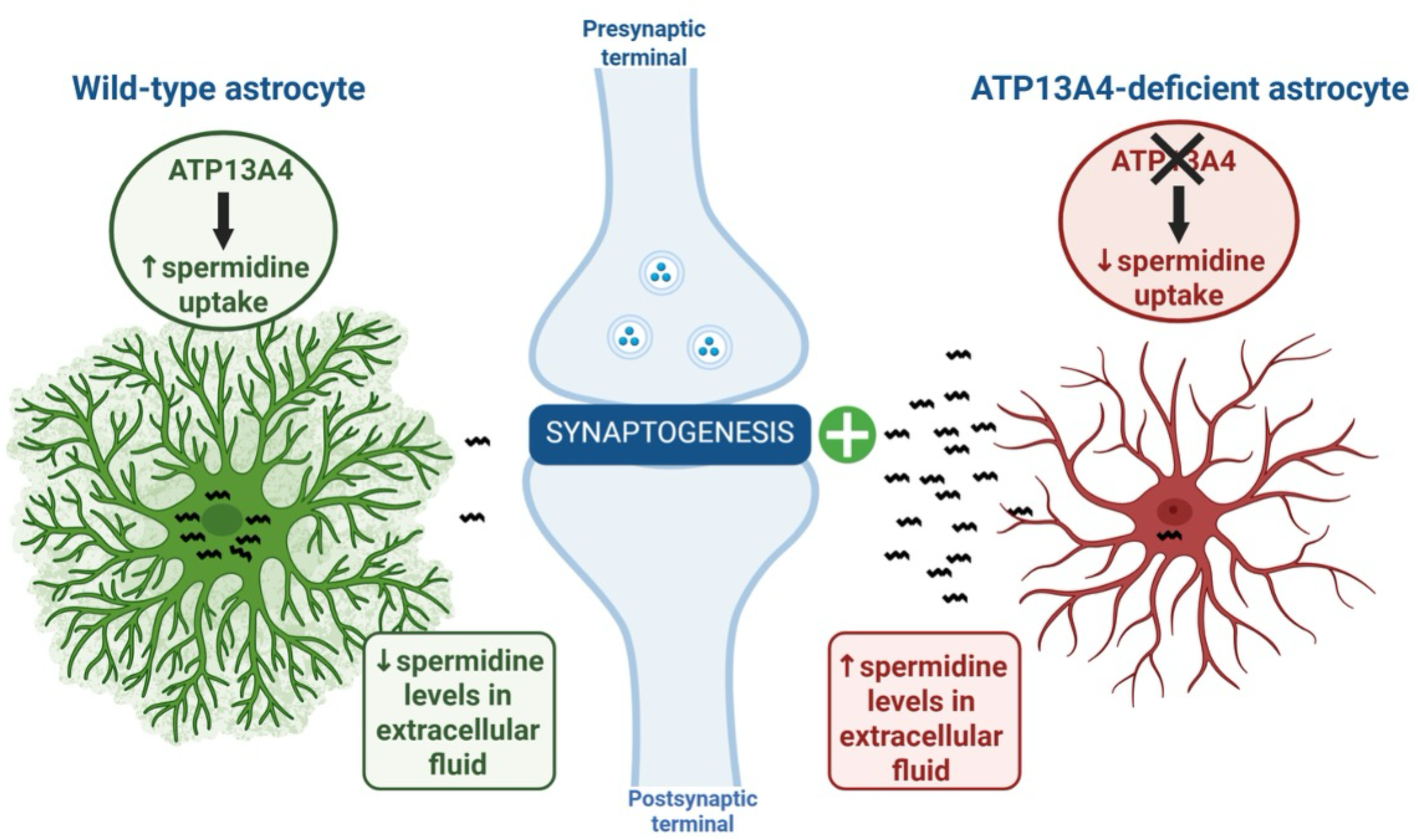
Astrocytic ATP13A4 restricts extracellular spermidine to control excitatory synaptogenesis. Illustration of the role of ATP13A4 in astrocytic spermidine uptake and its impact on astrocyte arborization and synaptogenesis. Created in BioRender. van Veen, S. (2026) https://BioRender.com/kd5oeym.

### ATP13A4 governs brain polyamine homeostasis

A central advance of this study is the demonstration that *in vivo* ATP13A4 regulates polyamine distribution across brain compartments. Targeted metabolomic analyses revealed a striking divergence between intracellular and extracellular polyamine pools, with selective depletion of specific polyamine species in cortical tissue and concomitant accumulation in CSF. Although CSF does not directly measure local interstitial or perisynaptic polyamine concentrations, these findings provide *in vivo* evidence for altered extracellular polyamine compartmentalization following ATP13A4 loss. The opposing changes observed in cortical tissue and CSF support a model in which ATP13A4-mediated uptake contributes to limiting extracellular polyamine accumulation in the brain. Importantly, systemic polyamine levels were unaffected, indicating that ATP13A4 loss induces a brain-specific redistribution rather than a global metabolic defect.

Consistent with this, ATP13A4-deficient astrocytes exhibit increased extracellular spermidine levels in conditioned media, linking impaired uptake to extracellular accumulation. The redistribution of polyamines is accompanied by a coordinated rewiring of polyamine metabolism, characterized by upregulation of the biosynthetic enzyme ODC1 and downregulation of the catabolic enzyme SMOX. This shift toward a polyamine-conserving state likely reflects a compensatory response to reduced intracellular polyamine availability resulting from impaired ATP13A4-mediated uptake. However, these adaptations fail to restore polyamine homeostasis, indicating that ATP13A4-mediated transport represents a major mechanism controlling intracellular polyamine availability in astrocytes. Taken together, these findings highlight the tight coupling between polyamine transport and metabolism and underscore the importance of astrocytic uptake in maintaining local biochemical equilibrium.

Given the well-established role of astrocytes in buffering extracellular neurotransmitters and ions (38–40), our findings extend this paradigm to metabolic signaling molecules and position astrocytes as key regulators of extracellular polyamine tone (31, 32). By controlling the balance between intracellular and extracellular polyamine pools, ATP13A4 establishes a compartmentalized metabolic framework that underlies both astrocyte function and neuron-glia communication. Although ATP13A4 is glia-enriched rather than strictly astrocyte-specific and the knockout model is constitutive, the convergence of transcriptomic, primary astrocyte and *in vivo* metabolomic data supports a predominant astrocyte-centered mechanism, while not excluding contributions from other cell types.

### Mechanistic model of ATP13A4-mediated polyamine transport

Our data support a model in which ATP13A4 facilitates cytosolic access of polyamines from extracytosolic compartments. This process is at least partially dependent on endocytosis, consistent with mechanisms described for ATP13A2 (5) and ATP13A3 (72). The relative contribution of endocytosis likely depends on ATP13A4 expression levels and its subcellular distribution between the plasma membrane and endocytic compartments. Notably, ATP13A3 dynamically redistributes from endosomes to the plasma membrane under conditions of polyamine depletion (73). In this context, ATP13A4 may operate in concert with the previously characterized polyamine transporter SLC18B1 (VPAT), mediating intracellular vesicular storage and secretion of polyamines in different CNS cell types, including astrocytes (114, 115). While ATP13A2 is also broadly expressed in the CNS, including in astrocytes, it has been primarily linked to neuroinflammation and neurodegeneration (116, 117), suggesting that both polyamine transporters may exert distinct but complementary neuroprotective roles. Together, these observations support a model in which distinct but functionally interconnected polyamine transporters coordinate cytosolic uptake, vesicular storage, and extracellular buffering. This integrated network establishes a multilayered regulatory system that governs polyamine homeostasis in the brain.

### ATP13A4 regulates synaptogenesis by controlling extracellular spermidine

Our data identify extracellular spermidine as a bioactive signal that promotes excitatory synaptogenesis. Using complementary approaches, including astrocyte-conditioned media, quantitative metabolomics, and defined spermidine supplementation, we show that increased extracellular spermidine is sufficient to enhance excitatory synapse formation. These effects occur at low nanomolar concentrations, indicating that neurons are highly sensitive to small changes in extracellular polyamine availability. Together, these observations position spermidine alongside established astrocyte-derived synaptogenic factors and suggest that metabolites can function as signaling molecules that directly influence synaptic development, further establishing astrocytes as active regulators of synaptogenesis and circuit formation (42–51, 53, 61, 65, 118). In this context, ATP13A4 acts as an upstream regulator of a spermidine-dependent signaling axis that links astrocytic polyamine handling to neuronal connectivity and circuit formation. Unlike classical synaptogenic cues – such as thrombospondins (46, 119), hevin (120), and cholesterol (121) – polyamines are small, diffusible molecules whose extracellular availability is tightly regulated under physiological conditions, in part through astrocyte-mediated uptake (31, 32, 37). To our knowledge, this is the first evidence that a polyamine functions as a synaptogenic signal, revealing a previously unrecognized mechanism of astrocyte-neuron communication.

While these findings support a synaptogenic role for extracellular spermidine, its direct contribution to synapse formation *in vivo* remains to be established. Astrocyte-derived synaptogenic factors typically act through specific neuronal receptors or scaffolding proteins (42, 68), but the precise pathway and/or receptor through which spermidine enhances excitatory synapse formation remains unknown. One possibility is direct modulation of ionotropic glutamate receptors, as N-methyl-D-aspartate receptors (NMDARs) contain extracellular polyamine binding sites (112) and are sensitive to polyamine-dependent changes in channel conductance, gating, and rectification. However, these receptors typically display equal or greater sensitivity to spermine than spermidine (122–124), making them less likely to fully account for the predominantly spermidine-dependent effects observed here. Alternatively, spermidine may act through eIF5A hypusination, a spermidine-dependent post-translational modification (125) that regulates local translation in neurons and supports the synthesis of synaptic proteins, including scaffolding proteins and cytoskeletal regulators critical for synaptogenesis and synaptic plasticity (126–128). Given the prolonged exposure paradigm used in our experiments, such translation-dependent mechanisms are compatible with the observed synaptic changes. In addition, polyamines have been shown to regulate postsynaptic receptor trafficking and clustering, as well as cytoskeletal dynamics, processes that may contribute to the increased number of excitatory synapses observed upon polyamine accumulation. Given that glutamatergic synapses are particularly sensitive to changes in local protein synthesis and receptor organization, these mechanisms may underlie the selective effects on excitatory synapses. While these mechanisms remain to be tested directly, defining spermidine-responsive signaling pathways and their cell type-specific engagement will be essential to establish the causal link between extracellular polyamine accumulation and synaptic remodeling.

### ATP13A4 and spermidine regulate astrocyte arborization and morphology

Beyond its cell non-autonomous role in regulating extracellular polyamine signaling, ATP13A4-dependent polyamine homeostasis also shapes astrocyte-intrinsic properties, including their morphological maturation. Loss of ATP13A4 consistently impaired astrocyte arborization across knockdown, knockout, and *in vivo* labeling approaches, indicating that astrocyte development is highly sensitive to polyamine availability. Notably, exogenous polyamines phenocopied these morphological defects in a dose-dependent manner, supporting the idea that elevated extracellular or mislocalized polyamines negatively impact astrocyte maturation, likely reflecting impaired intracellular handling of polyamines upon ATP13A4 loss. Given the essential role of astrocyte morphology in synapse formation and circuit organization, these findings underscore the need to tightly regulate extracellular polyamine levels in the brain and support a model in which disrupted polyamine compartmentalization alters astrocyte morphology and function, with downstream consequences for neuronal connectivity.

### ATP13A4 loss shapes neurodevelopment and circuit function

Our findings have direct implications for human disease. We show that multiple ATP13A4 variants associated with neurodevelopmental disorders impair transporter function, supporting a loss-of-function mechanism. We showed *in vivo* that ATP13A4 loss disrupts astrocytic polyamine uptake leading to extracellular polyamine imbalance, altered excitatory synaptogenesis, and impaired circuit maturation. Consistent with this, *Atp13a4* knockout mice exhibit delays in postnatal developmental milestones, including impairments in sensorimotor reflexes and delayed eye opening, indicating that ATP13A4-dependent polyamine regulation is required for timely circuit maturation. Notably, these developmental impairments are largely sex-independent and become most apparent during later postnatal stages, coinciding with astrocyte maturation and peak ATP13A4 expression. This temporal profile suggests that ATP13A4 function becomes increasingly critical as astrocytes acquire their roles in regulating extracellular signaling and synaptic refinement.

In contrast to the robust early phenotype, adult behavioral alterations are subtle and more pronounced in females, suggesting that early circuit perturbations are at least partially compensated during maturation. This aligns with neurodevelopmental models in which homeostatic mechanisms buffer early disruptions, resulting in mild or context-dependent adult phenotypes. The predominance of behavioral effects in females, despite largely shared molecular alterations across sexes, suggests that sex-dependent compensatory mechanisms act downstream of the primary polyamine imbalance, potentially reflecting differences in astrocyte-neuron interactions or circuit sensitivity to altered excitatory-inhibitory balance (129, 130). Importantly, current clinical evidence for ATP13A4-associated disorders does not indicate a consistent sex-dependent phenotype, although the limited number of reported cases precludes definitive conclusions.

Despite mild behavioral phenotypes, female *Atp13a4* KO mice exhibit increased excitatory synapse density and elevated mEPSC frequency, consistent with a shift in excitatory-inhibitory balance. Enhanced motor performance in female KO mice parallels observations in neurodevelopmental models such as *Cntnap2* mutants, where moderate corticostriatal circuit alterations can facilitate motor learning despite underlying deficits (131–133). Notably, *Cntnap2* KO mice also exhibit increased seizure susceptibility due to the loss of cortical inhibitory interneurons (134). Increased excitatory drive may similarly contribute to the behavioral alterations in female *Atp13a4* KO mice. We employed a constitutive Atp13a4 KO model that captures the systemic genetic context of human ATP13A4 deficiency, thereby strengthening the translational relevance of the observed phenotypes. However, as this model is not cell type-specific, contributions from non-astrocytic cell types cannot be excluded; future conditional approaches will be required to define the astrocyte-specific contribution and to assess whether ATP13A4 dysfunction converges on similar circuit mechanisms.

## Conclusion

The identification of extracellular spermidine as a synaptogenic cue, regulated by astrocytic ATP13A4-mediated uptake, defines a previously unrecognized metabolic axis linking glial polyamine handling to excitatory circuit formation. Given that polyamine dysregulation and astrocyte dysfunction have been implicated in neurodevelopmental disorders, including epilepsy and intellectual disability (25, 29, 30, 135–137), and that ATP13A4 is significantly downregulated in astrocytes from patients with temporal lobe epilepsy (105), our findings provide a mechanistic framework connecting impaired astrocytic polyamine clearance to disease-relevant alterations in circuit development and function. Although the germline knockout model likely underestimates astrocyte-specific contributions, the convergence of biochemical, cellular, *in vivo*, and human genetic evidence supports ATP13A4 as a central regulator of extracellular polyamine signaling that shapes excitatory circuit formation. These findings open new avenues for understanding how disrupted glial metabolism contributes to neurodevelopmental disease and position astrocytic polyamine uptake as a potential target for therapeutic intervention.

## Methods

### Clinical presentation and history

Patient LCTS01 is an individual evaluated in childhood for a global developmental delay with accompanying neuromuscular and behavioral symptoms. Cognitive assessment indicated below-average performance, and subtle physical features were noted. This study received ethical approval from the University of Leipzig (402/16-ek). Written informed consent for molecular genetic testing and data publication was obtained from the individual’s legal representatives.

Patient LCTS02 is an individual with epilepsy, with seizure onset in adolescence and a history of brief episodic motor events. The first major seizure was characterized by unresponsiveness and motor stiffening following a day of physical exertion. Initial clinical investigations were unremarkable. The individual was subsequently treated with anti-epileptic medication, with no recurrence of generalized seizures. Brief motor symptoms persist, occurring more frequently in the morning or during periods of physiological stress. Written informed consent for molecular genetic testing and data publication was obtained from the individual’s legal representatives.

Clinical descriptions have been de-identified to protect patient privacy.

### Animal studies

All animal experiments were approved by the local ethical committees: according to the KU Leuven ethical guidelines and approved by the KU Leuven Ethical Committee for Animal Experimentation (P167/2023, experiments involving the Atp13a4(KO) mouse strain), or alternatively, in accordance with the Institutional Animal Care and Use Committee and with oversight by the Duke Division of Laboratory Animal Resources under Institutional Animal Care and Use Committee Protocol Numbers A147-17-06 and A117-20-05. All animals were housed under standard conditions with continuous access to food and water, following a diurnal light/dark cycle (12-h/12-h), except for Atp13a4(KO) mice, which were maintained on a 14-h light/10-h dark cycle. Aldh1L1-EGFP (RRID: MMRRC_011015-UCD) mice were obtained through the Mutant Mouse Resource and Research Center (MMRRC) and backcrossed on a C57BL/6J background. Timed-pregnant WT CD1 mice (RRID: IMSR_CRL:022) used for PALE and wildtype Crl:CD(SD) Sprague-Dawley rats (RRID: RGD_734476) used for primary culture preps were purchased from Charles River Laboratories. The Atp13a4(KO) mouse strain, C57BL/6NJ-*Atp13a4^em1(IMPC)Bay^*/Mmnc (RRID:MMRRC_050715-UNC) was obtained from MMRRC at University of North Carolina at Chapel Hill, an NIH-funded strain repository, and was donated to the MMRRC by Arthur Beaudet, Ph.D., Baylor College of Medicine. Mice and rats of both sexes were included in all experiments.

### Cell culture and lentiviral transduction

#### H4 Cell Culture

H4 human neuroglioma cells (Cat# HTB-148, RRID:CVCL_1239) were obtained from ATCC and maintained in Dulbecco’s Modified Eagle Medium (DMEM; GIBCO, Cat# 41965039), supplemented with 10% heat-inactivated fetal bovine serum (FBS; PAN BioTech, Cat# P30-3306), 2% penicillin/streptomycin (Sigma, Cat# P4458), 1% non-essential amino acids (Sigma, Cat# M7145), and GlutaMAX™ (Thermo Fisher, Cat# 35050038). H4 cells were transduced with lentiviral vectors to obtain stable overexpression of firefly luciferase (FLUC) or human ATP13A4 (WT (ID: Q4VNC1), indicated catalytic or disease-associated variants). The catalytically dead mutant p.D486N and variants associated with autism (p.A356V (101)) and/or childhood apraxia of speech (p.E646D (101, 102, 138)) were generated. All cell lines were produced at varying viral vector titers and assessed for equal expression to WT ATP13A4. After lentiviral transduction, cells were selected with 2 μg mL^−1^ puromycin (Invivogen, Cat# ant-pr-1). The protocol for generating stable cell lines using lentiviral vectors is also described in protocols.io (doi.org/10.17504/protocols.io.bw57pg9n).

#### C8-D1A Cell Culture

C8-D1A mouse astrocytic cells (Cat# CRL-2541, RRID:CVCL_6379) were purchased from ATCC and cultured in DMEM (GIBCO, Cat# 41965039) supplemented with 10% heat-inactivated FBS (PAN BioTech, Cat# P30-3306), 2% penicillin/streptomycin (Sigma, Cat# P4458), 1% non-essential amino acids (Sigma, Cat# M7145), 1 mM sodium pyruvate (GIBCO, Cat# 11360-070), and GlutaMAX™ (Thermo Fisher, Cat# 35050038). C8-D1A cells with stable knockdown of Atp13a4 (miR2: 5’-GCCAATATTTCAGCAGCTTATT-3’, miR3: 5’-AGCCCATGAACTTCAAGCTCTA-3’, miR4: 5’-GCGATACCTAAACAGTTATCAA-3’) were produced via lentiviral vector transduction, as described in doi.org/10.17504/protocols.io.bw57pg9n. A miRNA targeting firefly luciferase (miRFLUC: 5’-GCGCTGAGTACTTCGAAATGTC-3’), was used as a negative control. After lentiviral transduction, cells were selected with 5 μg mL^−1^ blasticidin (Invivogen, Cat# ant-bl-05).

#### HEK293T Cell Culture

HEK293T/17 cells (Cat# CRL-11268, RRID:CVCL_1926) were obtained from ATCC and cultured in DMEM (GIBCO, Cat# 41965039) with 8% heat-inactivated FBS (PAN BioTech, Cat# P30-3306) and 2% penicillin/streptomycin (Sigma, Cat# P4458). HEK293T cells were transduced with lentiviral vectors, as described in doi.org/10.17504/protocols.io.bw57pg9n, to obtain stable overexpression of hATP13A4 (WT, indicated catalytic or disease variants) or mAtp13a4 (WT or Δexon 3-6). After lentiviral transduction, cells were selected with 2 μg mL^−1^ puromycin (Invivogen, Cat# ant-pr-1).

#### HeLa Cell Culture and Transfection

HeLa cells (ATCC, Cat# CCL-2, RRID:CVCL_0030) were obtained from Synthego and cultured in DMEM (GIBCO, Cat# 41965039) with 10% heat-inactivated FBS (PAN BioTech, Cat# P30-3306) and 2% penicillin/streptomycin (Sigma, Cat# P4458). For the subcellular localization of ATP13A4, HeLa cells were transiently transfected with pcDNA3.1-mCherry-msAtp13a4 (RRID:Addgene_234055) using PEI MAX^TM^ (Polysciences, Cat# 24765-100) at a 1:4 ratio, following the manufacturer’s instructions, in 15 cm culture dishes. 24 h post-transfection, cells were trypsinized and reseeded onto coverslips. An additional 24 h later, coverslips were fixed (30 min, 37 °C) with 4% paraformaldehyde (PFA; Thermo Fisher, Cat# J61899.AP) for subsequent analysis. This protocol is described in more detail on protocols.io and can be accessed at doi.org/10.17504/protocols.io.kxygxwy4kv8j/v1.

#### General Culture Conditions

All cell lines described above were maintained at 37 °C in a humidified incubator with 5% CO₂. Routine mycoplasma testing was performed, and all cultures were confirmed negative. Heat-inactivated FBS was used to deplete polyamine oxidase activity, which could interfere with fluorescent polyamine probes or supplemented polyamines.

### Immunocytochemistry - subcellular localization of ATP13A4

After PFA fixation, cells were washed with DPBS containing 0.5% Tween-20 (PanReac AppliChem, Cat# A4974.0500) (PBS-T) and permeabilized with 0.5% Triton X-100 (Sigma, Cat# 93443) for 30 min, followed by incubation in 0.1 M glycine (Sigma, Cat# G8898) for 1 h. Blocking was done for 1 h in PBS-T supplemented with 10% FBS (PAN BioTech, Cat# P30-3306) and 1% w/v BSA (Sigma, Cat# A4161). The cells were then incubated overnight at 4 °C with primary antibodies (diluted 1:200 in PBS-T with 1% FBS and 0.1% BSA). The following primary antibodies were used: anti-EEA1 (BD Biosciences, Cat# 610457, RRID:AB_397830), anti-CD63 (EXBIO Praha, Cat# 11-343-C025, RRID:AB_10755612), anti-LAMP1 (Cell Signaling Technology, Cat# 9091, RRID:AB_2687579) and anti-TOMM22 (Sigma, Cat# HPA003037, RRID:AB_1080329).

The next day, cells were re-blocked for 15 min before incubation with Alexa-Fluor 488 labeled secondary antibodies (30 min, diluted 1:1,000 in PBS-T containing 1% FBS and 0.1% w/v BSA). Nuclei were stained with DAPI (1:5,000 in PBS-T; Thermo Fisher, Cat# 62248) for 15 min. Between each step, the samples were washed thoroughly with PBS-T. After staining, the samples were mounted, and images were acquired using a Zeiss LSM880 confocal microscope with a 63× objective (Cell and Tissue Imaging Cluster (CIC), supported by Hercules AKUL/15/37_GOH1816N and FWO G.0929.15 to Pieter Vanden Berghe, University of Leuven). This protocol is described in more detail on protocols.io: doi.org/10.17504/protocols.io.kxygxwy4kv8j/v1.

### ATP13A4 purification

#### Purification of ATP13A4 from yeast

This protocol is described in more detail on protocols.io and can be accessed at doi.org/10.17504/protocols.io.ewov1d9jpvr2/v1. In brief, the *S. cerevisiae* BY4743; MATa/MATα strain (his3Δ1/his3Δ1, leu2Δ0/leu2Δ0, LYS2/lys2Δ0, met15Δ0/MET15; ura3Δ0/ura3Δ0, YPL217c/YPL217c::kanMX4; a kind gift from J. Winderickx, KU Leuven) was transformed according to the lithium acetate/single-stranded carrier DNA/polyethylene glycol method, with the pYSG-IBA162 vector containing a yeast codon-optimized version of human *ATP13A4* with C-terminal Twin-Strep tag (pYSG-IBA162-FLAG-hATP13A4-TwinStrep). The transformation mixture was grown for 48 h at 30 °C on minimal medium agar plates lacking uracil [0.54% yeast nitrogen base without amino acids (Sigma, Cat# Y0626), 0.12% yeast drop-out mix without uracil (Sigma, Cat# Y1501), 2% glucose (Sigma, Cat# G8270) and 2% agar (BD, Cat# 214010)] to select yeast colonies that acquired the plasmid. These colonies were then cultured in 20 mL of MM-Ura medium (0.67% yeast nitrogen base without amino acids, 0.19% yeast drop-out mix without uracil, and 2% glucose) and grown for 24 h at 28 °C and 175 rpm. The MM-Ura yeast pre-culture was used to inoculate 200 mL of MM-Ura medium to a final OD600 of 0.2, followed by a 12 h incubation period (28 °C and 175 rpm). The second pre-culture was inoculated into 4 L of MM-Ura medium to a final OD600 of 0.05 and grown overnight (28 °C and 175 rpm). Yeast cell pellets were collected via centrifugation (1,000*g*, 10 min, 4 °C) and resuspended in 4 L of MM-Leu medium [0.67% yeast nitrogen base without amino acids, 0.19% yeast drop-out mix without leucine (Sigma, Cat# Y1376), and 2% glucose], while ATP13A4 expression was induced with 1 mM CuSO4 (Sigma, Cat# C1297). After another 24 h, the pellet was collected (1,000*g*, 10 min, 4 °C) and yeast membrane fractions were prepared as in (5). In brief, yeast cells were disrupted using glass beads (Sigma, Cat# G8772) in a BeadBeater (BioSpec products, Cat# 1107900-105). The lysis buffer was composed of 50 mM Tris-HCl (pH 7.5) (Tris: Sigma, Cat# T1503; HCl: Merck, Cat# 1.00317.2501), 1 mM EDTA (BDH Laboratory Supplies, Cat# 280214S), 0.6 M sorbitol (Sigma, Cat# S1876), 1 mM phenylmethylsulfonyl fluoride (Sigma, Cat# 93482), and SigmaFast protease inhibitor cocktail (Sigma, Cat# S8830). To eliminate cell debris and nuclei, the crude lysate was centrifuged at 2,000*g* for 20 min at 4 °C. The resulting supernatant (S1) underwent further centrifugation at 20,000*g* for 20 min at 4 °C to sediment the heavy membrane fraction (P2), and the subsequent supernatant (S2) was then centrifuged at 200,000*g* for 1 h at 4 °C. The final pellet, which represents the light membrane fraction (P3), was resuspended in 20 mM HEPES-Tris (pH 7.4) (HEPES: Sigma, Cat# H3375), 0.3 M sucrose (Sigma, Cat# S7903), and 0.1 mM CaCl2 (Sigma, Cat# C3881). The total protein concentration was measured using a Bradford assay (Sigma, Cat# B6916).

To purify ATP13A4, yeast membranes were first diluted in solubilization buffer [50 mM MOPS-KOH (pH 7) (MOPS: VWR, Cat# A1076.1000; KOH: Sigma, Cat# 221473), 100 mM KCl (Sigma, Cat# P9541), 5 mM MgCl2 (Sigma, Cat# M2670), 20% glycerol (Carl Roth, Cat# 3783.5), 1 mM dithiothreitol (DTT; VWR, Cat# A2948.0025), 0.5% (w/v) n-dodecyl-β-D-maltoside (DDM; Inalco, Cat# 1758-1350) and SigmaFast protease inhibitor cocktail] to a total protein concentration of 5 mg mL^−1^. The samples were stirred on ice for 60 min, followed by centrifugation (200,000*g*, 1 h, 4 °C) to pellet non-solubilized membranes. The solubilized material was incubated with Strep-Tactin®XT 4Flow® high-capacity resin (IBA Lifesciences, Cat# 2-5030-002) for 3 h (4 °C) to enable binding of TwinStrep-tagged ATP13A4 to the resin. To eliminate unbound material, the resin was washed five times with 10 column volumes of wash buffer [50 mM MOPS-KOH (pH 7), 100 mM KCl, 5 mM MgCl2, 20% glycerol, 1 mM DTT, 0.05% (w/v) DDM and SigmaFast protease inhibitor cocktail]. Finally, ATP13A4 was eluted with wash buffer supplemented with 50 mM biotin (IBA Lifesciences, Cat# 2-1016-002).

The quality of the purification was evaluated via SDS-PAGE followed by Coomassie staining or immunoblotting, as described on protocols.io at doi.org/10.17504/protocols.io.14egn96rql5d/v1. The protein concentration was estimated by running an SDS-PAGE gel, staining it with Coomassie dye, and comparing the intensity of the protein bands to a standard curve made with known amounts of BSA, as described on protocols.io (doi.org/10.17504/protocols.io.kqdg3qqqev25/v1).

#### Purification of ATP13A4 from HEK293T cells

HEK293T cells stably overexpressing ATP13A4 (described above) were resuspended in LIS buffer [10 mM Tris-HCl (pH 7.5) (Tris: Sigma, Cat# T1503; HCl: Merck, Cat# 1.00317.2501), 0.5 mM MgCl2 (Sigma, Cat# M2670), 1 mM DTT (VWR, Cat# A2948.0025), and SigmaFast protease inhibitor cocktail (Sigma, Cat# S8830)] and incubated for 15 min. The cells were then homogenized with 60 strokes using a Dounce homogenizer (DWK Life Sciences, Cat# 357546). Afterwards, an equal volume of 1M buffer [10 mM Tris-HCl (pH 7.3), 0.5 M sucrose (Sigma, Cat# S7903), 40 µM CaCl2 (Sigma, Cat# C3881), 0.23 mM PMSF (Sigma, Cat# 93482), 1 mM DTT] was added, and an additional 30 strokes of homogenization were performed. The cell lysate was centrifuged at 500*g* for 20 min, and the resulting pellet was resuspended in LIS buffer and subjected to a second round of homogenization (60 strokes), followed by addition of an equal volume of 1M buffer and 30 additional strokes. The lysate was centrifuged again at 500*g* for 20 min, and the supernatants from both centrifugation steps were combined. The protein concentration of the combined supernatant was measured using a Nanodrop spectrophotometer (Thermo Fisher). For membrane solubilization, three times the amount of DDM (Inalco, Cat# 1758-1350), supplemented with cholesteryl hemisuccinate (CHS; Sigma, Cat# C6013) at a DDM:CHS ratio of 10:1, was added to the supernatant and incubated for 3 h. Non-solubilized membranes were removed by centrifugation at 15,557*g* for 1 h at 4 °C. The solubilized membranes were incubated with Strep-Tactin®XT 4Flow® high-capacity resin (IBA Lifesciences, Cat# 2-5030-002) for 16 h at 4 °C. After binding, the beads were washed twice with 10 column volumes of wash buffer 1 [20 mM Tris-HCl, pH 7.5, 300 mM NaCl (Carl Roth, Cat# 0962.2), 0.03% (w/v) DDM, 0.003% (w/v) CHS, 5 mM TCEP (Thermo Fisher, Cat# 77720)] and wash buffer 2 [20 mM Tris-HCl, pH 7.5, 150 mM NaCl, 0.03% (w/v) DDM, 0.003% (w/v) CHS, 5 mM TCEP]. Finally, ATP13A4 was eluted in 100 mM Tris-HCl (pH 8.0), 150 mM NaCl, 1 mM EDTA (BDH Laboratory Supplies, Cat# 280214S), 50 mM biotin (IBA Lifesciences, Cat# 2-1016-002), 0.03% (w/v) DDM, 0.003% (w/v) CHS, and 5 mM TCEP. The quality and concentration of the purified protein were assessed as described in the section outlining the purification of ATP13A4 from yeast. This protocol is described in more detail on protocols.io and can be accessed at doi.org/10.17504/protocols.io.dm6gp9z85vzp/v1.

### ATPase activity assay

To measure the ATPase activity of ATP13A4, a luminescence-based ADP-Glo Max assay (Promega, Cat# V7002) was employed to detect the rate of ATP hydrolysis, following a previously described protocol (5) with minor adjustments. Briefly, we prepared a 25 µl reaction mixture containing 100 ng purified ATP13A4, 11 mM MgCl2 (Sigma, Cat# M2670), 100 mM KCl (Sigma, Cat# P9541), 50 mM MOPS-KOH (pH 7) (MOPS: VWR, Cat# A1076.1000; KOH: Sigma, Cat# 221473), 1 mM DTT (VWR, Cat# A2948.0025), 0,001% (w/v) DDM (Inalco, Cat# 1758-1350), and specified amine concentrations. After incubating the mixture for 5 min at 37 °C, the reaction was initiated by adding 5 mM ATP and allowed to proceed for 1 h at 37 °C, followed by an additional 1 h incubation at room temperature. Next, 25 µl of ADP-Glo reagent was added and incubated for 1 h at room temperature, followed by the addition of 50 µl of ADP-Glo Max Detection Reagent and 1 h incubation in the dark at room temperature. Luminescence was measured using a Synergy H1 microplate reader (Biotek). Data analysis, including curve fitting, was performed using GraphPad Prism Software (RRID:SCR_002798). This protocol is described in more detail on protocols.io and can be accessed at doi.org/10.17504/protocols.io.5qpvo9kxdv4o/v1.

### Auto-phosphorylation assay

The autophosphorylation activity of ATP13A4 on the conserved p.D486 residue was measured using previously established methods (5, 95). In brief, P2 yeast membranes (40 μg) or purified ATP13A4 (1 μg) were incubated for 1 min with [γ-^32^P] ATP (2 μCi, 5.125 µM; Cat# BLU502A100UC; Revvity, Inc.) in the presence or absence of 1 mM of the specified polyamines. To assess the sensitivity of the ATP13A4 phosphoenzyme to ADP or ATP, non-radioactive ADP or ATP (Promega, Cat# V703) (5 mM each) was added to yeast P2 membranes (40 μg) 30 s after the addition of [γ-^32^P] ATP, and the reaction was stopped at the indicated times. In addition, to evaluate the sensitivity of the ATP13A4 phosphoenzyme to a combination of ATP and polyamines, non-radioactive ATP (5 mM) and polyamines (1 mM) were added 30 s after [γ-^32^P] ATP. The reaction was stopped by adding 400 μL of stop solution (20% trichloroacetic acid (Sigma, Cat# T9159), 10 mM phosphoric acid (Thermo Fisher, Cat# 424045000), followed by precipitation on ice for 30 min and centrifugation (20,000*g*, 30 min, 4 °C). The resulting pellet was washed twice with 400 μL of ice-cold stop solution and dissolved in sample buffer. Where indicated, an additional wash with 0.3 M hydroxylamine (HA; Sigma, Cat# 438227-50ML) was performed. Incorporation of ^32^P was visualized by SDS-PAGE under acidic conditions and detected via autoradiography (Typhoon™ FLA 7000, GE Healthcare). This protocol is described in more detail on protocols.io and can be accessed at doi.org/10.17504/protocols.io.kxygxw3rwv8j/v1.

### Cellular polyamine uptake assay

For flow cytometry-based analysis of BODIPY-polyamine uptake, cells were seeded in 12-well plates at a density of 1.0 × 10^5^ cells per well. The following day, cells were incubated with BODIPY-labeled polyamines [putrescine (Merck, Cat# SCT248), spermidine (Merck, Cat# SCT249) and spermine (Merck, Cat# SCT250)] for the specified times and concentrations (2 h at 1 μM for C8-D1A cells; 1 h at 5 μM for H4 cells). For endocytosis inhibition experiments, cells were pre-treated for 15 min with a combination of endocytosis inhibitors, i.e. Dynasore (100 µM; Sigma, Cat# D7693), Genistein (50 µM; Abcam, Cat# ab120112) and Pitstop 2 (50 µM; Sigma, Cat# SML1169) prior to addition of BODIPY-labeled polyamines. For low-temperature experiments, cells were incubated with BODIPY-labeled polyamines at 4 °C. After incubation, cells were trypsinized and resuspended in DPBS (GIBCO, Cat# 14190144) containing 1% BSA (Carl Roth, Cat# 8076.4). Cell suspensions were then filtered through a nylon mesh to remove any clumps and kept on ice until acquisition. Uptake of polyamines was assessed by recording the mean fluorescence intensities (MFI) of 10,000 events per sample using a flow cytometer (**Supplementary Fig. 6c-f**: BD FACSCanto™ II instrument, BD Life Sciences; **Supplementary Fig. 6g, i-k, p-q**: ID7000 spectral cell analyzer, Sony). This protocol is described in more detail on protocols.io and can be accessed at doi.org/10.17504/protocols.io.q26g7mpq1gwz/v1.

For confocal microscopy-based analysis of BODIPY-polyamine uptake, cells were seeded on cover slips placed in 12-well plates (C8-D1A) or 24-well plates (primary astrocytes) at a density of 5.0 × 10^4^ cells per well. The following day, cells were incubated for 2 h with 1 μM (C8-D1A) or 10 μM (primary astrocytes) BODIPY-conjugated polyamines. After incubation, cells were fixed in 4% PFA (Thermo Fisher, Cat# J61899.AP) for 30 min at 37 °C. Thereafter, cells were washed in DPBS and stored at 4°C. The next day, nuclei were stained using DAPI (Thermo Fisher, Cat# 62248), and cover slips were mounted onto slides. Images were captured on an Olympus FV 3000 microscope (primary astrocytes) or Zeiss LSM 880 (C8-D1A cells; Cell and Tissue Imaging Cluster (CIC), supported by Hercules AKUL/15/37_GOH1816N and FWO G.0929.15 to Pieter Vanden Berghe, University of Leuven). This protocol is described in more detail on protocols.io: doi.org/10.17504/protocols.io.5qpvo9kd9v4o/v1.

### RT-qPCR

RT-qPCR was used to evaluate *msAtp13a4* mRNA expression levels following lentiviral *msAtp13a4* knockdown in C8-D1A cells. RNA was extracted from 1.0 × 10^6^ cells following the protocol provided with the NucleoSpin RNA Plus Kit (Macherey-Nagel, Cat# 740984.250). RNA concentration and purity were evaluated using a Nanodrop spectrophotometer (Thermo Fisher). The isolated RNA was then reverse transcribed into cDNA using the High-Capacity cDNA Reverse Transcription Kit (Applied Biosystems^TM^, Cat# 4374966). SYBR Green-based qPCR was performed using ATP13A4-specific primers, with actin serving as the reference gene. The sequences of the forward (F) and reverse (R) primers used (5′→3′) are as follows: *msAtp13a4*: (F): 5’-CCAGCATGCTTTACTCAATG-3’, (R): 5’-GAAGATGGATCCAATGAGAC-3’; *msGapdh*: (F): 5’-TGTGTCCGTCGTGGATCTGA-3’, (R): 5’-CCTGCTTCACCACCTTCTTGA-3’.

The amplification was carried out on a Light Cycler (Roche) under the following conditions: initial denaturation at 95 °C for 10 min, followed by 50 cycles of 10 s at 95 °C, 30 s at 55 °C, 1 min at 95 °C, and 1 min at 55 °C. A melting curve analysis was conducted from 55 °C to 95 °C, and the mean Cq values were recorded for data analysis. This protocol is described in more detail on protocols.io and can be accessed at doi.org/10.17504/protocols.io.n92ldrq98g5b/v1.

### Protein extraction and immunoblotting

Samples were maintained on ice during processing. For brain samples, whole brains were homogenized in ice-cold RIPA buffer (Cell Biolabs, Cat# AKR-191) supplemented with protease inhibitor cocktail. Tissue was disrupted using a mechanical pestle, incubated on ice for 10-30 min, and clarified by centrifugation through QIAshredder columns (Qiagen, Cat# 79656) at 4 °C for 5 min at >10,000*g*. The resulting supernatant was collected in low protein-binding tubes. This protocol is described in more detail on protocols.io and can be accessed at doi.org/10.17504/protocols.io.yxmvm8jmng3p/v1.

Proteins from (primary) cell cultures were extracted using RIPA buffer (Thermo Fisher, Cat# 89901) containing protease inhibitors (Sigma, Cat# S8830). The cell lysates were incubated on ice for at least 30 min, and then centrifuged at 15,000*g* for 30 min at 4 °C to remove non-solubilized material. The resulting supernatant was collected and stored at - 80 °C. The preparation of cell lysates is described in more detail on protocols.io and can be accessed at doi.org/10.17504/protocols.io.eq2ly66wpgx9/v1.

Protein samples (20-40 μg) were separated on NuPAGE^TM^ 4–12% Bis-Tris precast gels (Invitrogen, Cat# NP0321, Cat# NP0322, Cat# NP0323) using MOPS running buffer (Invitrogen, Cat# NP0001) and then transferred onto polyvinylidene fluoride (PVDF) membranes (Millipore, #Cat IPFL00010) following the manufacturer’s protocol. Membranes were blocked in TBS-T [50 mM Tris-HCl (pH 7.5) (Tris: Sigma, Cat# T1503; HCl: Merck, Cat# 1.00317.2501), 150 mM NaCl (Carl Roth, Cat# 0962.2), 0.1% Tween-20 (PanReac AppliChem, Cat# A4974.0500)] with 5% non-fat dry milk. Primary antibodies were applied at a 1:1,000 dilution in TBS-T with 1% BSA and incubated overnight, followed by incubation with horseradish peroxidase (HRP)-conjugated secondary antibodies (1–2 h; 1:1,000 dilution in TBS-T with 1% milk). We used primary antibodies directed against ATP13A4 (71) (RRID: AB_3741656), streptavidin (Abcam, Cat# ab76949, RRID:AB_1524455), Flag-tag (Thermo Fisher Scientific, Cat# MA1-91878, RRID:AB_1957945), ODC1 (Abcam, Cat# ab97395, RRID:AB_10679334), SRM (Abcam, Cat# ab241496, RRID:AB_3741648), SMS (Abcam, Cat# ab248996, RRID: AB_3741655), SAT1 (Proteintech, Cat# 10708-1-AP, RRID:AB_2877739), SMOX (Sigma, Cat# SAB1101510, RRID:AB_10620451), β-actin (Sigma, Cat# A5441, RRID:AB_476744), tubulin (Sigma, Cat# T5168, RRID:AB_477579) and GAPDH (Sigma, Cat# G8795, RRID:AB_1078991). The rabbit polyclonal anti-ATP13A4 antibody was home-made and raised against the epitope ^1182^VSYSNPVFESNEEQL (71). Protein detection was performed using an enhanced chemiluminescence (ECL) substrate (Thermo Fisher, Cat# 32106) and visualized with the Bio-Rad ChemiDoc MP imaging system. Quantification was conducted using the ImageJ software (http://fiji.sc;RRID:SCR_002285). The immunoblotting protocol is described in more detail on protocols.io: doi.org/10.17504/protocols.io.14egn96rql5d/v1.

### Metabolomics

For metabolomic analysis of C8-D1A cells (**Supplementary Fig. 6r**), cells were first washed with 0.9% NaCl (Carl Roth, Cat# 0962.2), followed by the addition of 150 μL of ice-cold 6% trichloroacetic acid (TCA; Sigma, Cat# T9159) to extract metabolites. The cell lysate was collected using a cell scraper and incubated on ice for 30 min. Next, the lysate was centrifuged at 20,000*g* for 20 min at 4 °C. The supernatant was collected, flash-frozen in liquid nitrogen, and stored at-80 °C for future analysis. 1,6-Hexanediamine (Sigma, Cat# H11696) was added as an internal standard to correct for signal drift. Correction was only performed when doing so improved replicate reproducibility. To 100 μL of supernatant, 900 μL of 100 mM sodium carbonate buffer (pH 9.3) (Sigma, Cat# S7795-500g) was added. Then, 25 μL of isobutyl chloroformate (Sigma, Cat# 177989-25G) was introduced, and the mixture was incubated at 35 °C for 30 min. Afterward, 800 μL of the reaction mixture was transferred to a 2 mL Eppendorf tube, followed by the addition of 1 mL of diethyl ether (Sigma, Cat# 309966-1L). The mixture was vigorously vortexed and allowed to sit at 25 °C for 15 min. Subsequently, 900 μL of the upper phase was collected in a new Eppendorf tube and dried using a vacuum centrifuge. The dried extract was then dissolved in 125 μL of 50% acetonitrile (LC–MS grade; VWR, Cat# 83640.320) in water containing 0.1% formic acid (VWR, Cat# 84865.260) and transferred to an MS vial. Next, 10 μL of the extract was loaded onto a Vanquish LC System (Thermo Scientific) coupled via heated electrospray ionization to a Q Exactive Orbitrap Focus mass spectrometer (Thermo Scientific) operated in positive mode equipped with an ACQUITY UPLC HSS C18 (1.7 μm, 2.1 × 100 mm) column from Waters^TM^. Solvent A consisted of ultrapure H2O with 0.2% acetic acid while solvent B was acetonitrile with 0.2% acetic acid; all solvents used were LC-MS grade. Flow rate remained constant at 250 μL/min, and the column temperature remained constant at 30 °C. A gradient for the separation of modified polyamines was applied as follows: from 0 to 2 min 20% B, from 2 to 10 min a linear increase to 85% B was carried out and remained at 85% B until 17 min. At 18 min the gradient returned to 20% B. The method stopped at 22 min. The mass spectrometer operated in full scan (range 70.0000-750.0000) and positive mode using Sheath gas at 25, Aux Gas at 10. Ion Transfer tube was heated at 320 °C and Vaporizer temperature was set at 310 °C. The data analyses were performed by integrating the peak areas (El-Maven - Elucidata).

For metabolomic analysis of ACM samples (**Fig. 4d, Supplementary Fig. 10e-j**), as well as cortical brain tissue, plasma, and CSF (**Fig. 5a-g, Supplementary Fig. 11**), we used an improved method with higher throughput and reproducibility by doing cleanup on the LC system itself and including labeled internal standards.

For ACM and plasma samples, 50 μL of ice-cold TCA extraction buffer was added to 50 μL of sample. For cerebrospinal fluid (CSF), extraction was performed using a matched-volume approach to account for limited and variable sample input. Each CSF sample was combined with an equal volume of ice-cold 6% TCA extraction buffer. Due to the limited volume obtained per animal, a subset of CSF samples was pooled, prior to extraction, to ensure sufficient material for LC-MS/MS analysis. Pooling was performed across animals of the same genotype and sex, and pooled samples were processed identically to individual samples. In figures, individual samples are represented as filled symbols, whereas pooled samples are indicated as open symbols. For cortical brain tissue (15-30 mg), 250 μL of ice-cold TCA extraction buffer was used, and samples were homogenized in Ribolyser tubes to ensure thorough extraction. For absolute quantification of polyamine levels, internal standards were included in the extraction buffer. The standards used were^13^C5 ornithine (Cat# CLM-4724-H-PK), ^13^C4 putrescine (Cat# CLM-6574-PK), ^13^C4 spermidine (Cat# CLM-9435-PK) and D8 spermine (Cat# DLM-9262-PK) from Cambridge Isotope Laboratories, Inc.

To 40 μL of supernatant, 360 μL of 100 mM sodium carbonate buffer (pH 9.3) was added. Then, 10 μL of isobutyl chloroformate (Sigma) was introduced, and the mixture was incubated at 35 °C for 30 min. Afterward, the reaction mixture was transferred to an MS vial. 50 μL of the extract was loaded onto a Vanquish LC System (Thermo Scientific) coupled via heated electrospray ionization to a Q Exactive Orbitrap Focus mass spectrometer (Thermo Scientific) operated in positive mode equipped with an ACQUITY UPLC HSS T3 VanGuard Pre-column (100Å, 1.8 µm, 2.1 mm × 5 mm) from Waters^TM^ used as a separation column. Solvent A consisted of ultrapure H2O with 0.2% acetic acid while solvent B was acetonitrile with 0.2% acetic acid; all solvents used were LC-MS grade. A gradient for the separation of modified polyamines was applied as follows: Flow rate started at 1.8 mL/min with 5% B while running from column to the waste to prevent reagents from entering the mass spectrometer, changing from 5 to 35% B from 0.67 to 1 min. From 1 to 1.1 min the flow increased linearly from 1.8 mL/min to 0.3 mL/min and the % B increased to 36%, then the flow from the column was redirected to the MS. From 1.1 to 3 min a linear increase to 85% B was carried out and remained at 85% B until 4.7 min. From 4.7 min to 4.73 min the flow switched to the waste again, and increased to 1.8 mL/min. From 4.73 to 4.86 min %B was linearly decreased to 5% B and kept there for equilibration. The method stopped at 5.53 min. The column temperature remained constant at 25 °C. The mass spectrometer operated in full scan (range 70.0000-1050.0000) and positive mode using Sheath gas at 25, Aux Gas at 10. Ion Transfer tube was heated at 320 °C and Vaporizer temperature was set at 310 °C. The data analyses were performed by integrating the peak areas (El-Maven - Elucidata).

These protocols are described in more detail on protocols.io and can be accessed at doi.org/10.17504/protocols.io.dm6gp9z7dvzp/v1.

### Primary astrocyte and neuron cultures

#### shRNA plasmids

shRNA that targets mouse/rat Atp13a4 (5’-GCCCATGAACTTCAAGCTCTA-3’) or a scrambled sequence thereof (5’-GCTCACCACGTGCAATACTAT-3’) was cloned into the pLKO.1 TRC cloning vector (139) according to Addgene protocols (https://www.addgene.org/protocols/plko/). The shRNA sequence was verified by Basic Local Alignment Search Tool alignment to be specific to mouse and rat isoforms of *Atp13a4* and the scramble sequence was verified to have no known gene targets. pLKO.1 shRNA plasmids that express EGFP were synthesized from the pLKO.1 Puro by replacing the puromycin resistance gene with CAG-EGFP.

#### Cortical astrocyte and neuronal culture (rat)

Postnatal glia-free rat cortical astrocyte and neuronal cultures were prepared from WT Crl:CD(SD) Sprague-Dawley rats from Charles River Laboratories (RRID: RGD_734476). To obtain a single-cell suspension, the cerebral cortices were dissected in DPBS and enzymatically dissociated with papain (∼7.5 units mL^−1^; Worthington, Cat# LK003178) for 45 min at 33 °C. Next, the cortices were mechanically dissociated using low and high ovomucoid solutions. From this step, astrocyte and neuronal cultures diverge.

To obtain cortical astrocytic cultures the single-cell suspension was resuspended in astrocyte growth media (AGM; DMEM (GIBCO, Cat# 11960), 10% FBS (Thermo Fisher, Cat# 10-437-028), 10 μM hydrocortisone (Sigma, Cat# H0888), 100 U mL^−1^ penicillin/streptomycin (GIBCO, Cat# 15140), 2 mM L-glutamine (GIBCO, Cat# 25030-081), 5 μg mL^−1^ insulin (Sigma, Cat# I1882), 1 mM Na pyruvate (GIBCO, Cat# 11360-070), 5 μg mL^−1^ N-acetyl-L-cysteine (Sigma, Cat# A8199)) and filtered through a 20 μm mesh filter (Elko Filtering, Cat# 03-20/14). The cells were counted, and 20 million cells were seeded in T75 flasks coated with poly-*D*-lysine (PDL; Sigma, Cat# P6407). On day *in-vitro* (DIV) 3, the flasks were forcefully shaken to remove non-astrocyte cells, until only an adherent monolayer of astrocytes remained. From DIV 5 to DIV 7 AraC (Sigma, Cat# C1768) was added to the media to eliminate contaminating fibroblasts. On DIV 7, astrocytes were trypsinized (0.05% trypsin-EDTA) and plated into 12-well plates (200,000 cells/well) for neuron-astrocyte co-culture experiments and into 10 cm dishes (3 million cells per dish) for ACM production.

For neuron-astrocyte co-culture experiments, astrocytes were transfected with shRNA and/or expression plasmids using Lipofectamine^TM^ LTX Reagent with PLUS^TM^ Reagent (Invitrogen, Cat# 15338100) according to the manufacturer’s instructions at DIV 8. In brief, 2 μg of total DNA was diluted in Opti-MEM (GIBCO, Cat# 11058021) containing Plus Reagent, then combined with Opti-MEM containing LTX at a DNA-to-LTX ratio of 1:2 and incubated at room temperature for 30 min. The resulting transfection mixture was added to astrocyte cultures and incubated for 2.5 h at 37 °C in 10% CO2. On DIV 10, astrocytes were trypsinized, resuspended in NGM Plus, and plated at a density of 20,000 cells per well onto DIV 10 WT neurons or astrocytes. The co-culture was maintained for 48 h. The protocols for astrocyte isolation and transfection are described in more detail on protocols.io and can be accessed at doi.org/10.17504/protocols.io.bp2l6dy7kvqe/v1.

For ACM production, during the astrocyte passage step (DIV 7), the astrocytes were nucleofected with constructs carrying either a scramble sequence (shCtrl, 5’-GCTCACCACGTGCAATACTAT-3’) or shRNA (shAtp13a4, 5’-GCCCATGAACTTCAAGCTCTA-3’), using the Basic Nucleofector Kit for Primary Mammalian Glial Cells (Lonza, Cat# VPI-1006) according to the manufacturer’s protocol. On DIV 8 the media was changed with fresh AGM. On DIV 9, the media was replaced with minimal media (Neurobasal medium minus phenol red (GIBCO, Cat# 12348017), 100 U mL^−1^ penicillin/streptomycin (GIBCO, Cat# 15140), 2 mM L-glutamine (GIBCO, Cat# 25030-081), 1 mM Na pyruvate). At DIV 12, ACM was collected, filtered, and concentrated using Vivaspin 20, 5 kDa MWCO (Sartorius, Cat# VS2012). The protocols for astrocyte isolation and ACM production are described in more detail on protocols.io and can be accessed at doi.org/10.17504/protocols.io.261gedp57v47/v1.

To obtain pure cortical neuronal cultures, the single-cell suspension was resuspended in panning buffer (0.02% BSA (Sigma, Cat# A4161) with 0.5 μg mL^−1^ insulin (Sigma, Cat# I1882) in DPBS with calcium, magnesium, glucose, and pyruvate (Thermo Fisher, Cat# 14287080)) and filtered through a 20 μm mesh filter (Elko Filtering, Cat# 03-20/14). The filtered cells were incubated on sequential negative panning dishes coated with the following: i) *Bandeiraea simplicifolia* Lectin 1 (Vector Laboratories, Cat# L-1100) (twice), ii) goat anti-mouse IgG+IgM (H+L) (Jackson ImmunoResearch, Cat# 115-005-044, RRID:AB_2338451), iii) goat anti-rat IgG+IgM (H+L) (Jackson ImmunoResearch, Cat# 112-005-044, RRID: AB_2338094). The cells were incubated on positive panning dishes coated with mouse anti-rat L1 (Developmental Studies Hybridoma Bank, Cat# ASCS4, RRID:AB_528349) for 45 min to isolate cortical neurons. After this final incubation, the positive panning dishes were washed with panning buffer and the neurons were collected by forceful pipetting with a P1000 pipette. The isolated neurons were centrifuged (11 min, 200*g*) to obtain a pellet and resuspended in serum-free neuron growth media [NGM; Neurobasal (GIBCO, Cat# 21103049), B27 supplement (GIBCO, Cat# 17504044), 2 mM L-glutamine (GIBCO, Cat# 25030-081), 100 U mL^−1^ penicillin/streptomycin (GIBCO, Cat# 15140), 1 mM sodium pyruvate (GIBCO, Cat# 11360-070), 4.2 μg mL^−1^ forskolin (Sigma, Cat# F6886), 50 ng mL^−1^ brain-derived neurotrophic factor (BDNF; PeproTech, Cat# 450-02), and 10 ng mL^−1^ ciliary neurotrophic factor (CNTF; PeproTech, Cat# 450-13)]. The cells were then counted and subsequently plated.

For astrocyte-neuron co-culture assays, 100,000 neurons were plated onto 12-mm glass coverslips coated with 10 μg mL^−1^ PDL (Sigma, Cat# P6407) and 2 μg mL^−1^ Laminin (R&D Systems, Cat# 3400-010-02). For neuron morphology and synapse assays, 80,000 and 60,000 neurons, respectively, were plated onto similarly coated coverslips. The neurons were then incubated at 37 °C in 10% CO2. On DIV 2, half of the media was replaced with NGM Plus [Neurobasal Plus (GIBCO, Cat# A3582901), B27 Plus (GIBCO, Cat# A3582801), 100 U mL^−1^ penicillin/streptomycin (GIBCO, Cat# 15140), 1 mM sodium pyruvate (GIBCO, Cat# 11360-070), 4.2 μg mL^−1^ forskolin (Sigma, Cat# F6886), 50 ng mL^−1^ BDNF (PeproTech, Cat# 450-02), and 10 ng mL^−1^ ciliary neurotrophic factor (CNTF; PeproTech, Cat# 450-13)] supplemented with AraC (4 μM; Sigma, Cat# C1768) to inhibit the growth of proliferating contaminating cells. On DIV 3, all the media was replaced with NGM Plus to remove any traces of AraC. On DIV 6, DIV 8 and DIV 11, the neurons were fed by replacing half of the media with NGM Plus. The protocol for neuron isolation is described in more detail on protocols.io and can be accessed at doi.org/10.17504/protocols.io.36wgq3r35lk5/v1.

#### Cortical astrocyte and neuronal culture (mouse)

Primary cortical astrocyte and neuronal cultures were prepared from P1 *Atp13a4* WT and KO mice obtained from homozygous breedings.

Following removal of meninges, olfactory bulb, cerebellum, and brainstem, cerebral cortices were dissected in ice-cold astrocyte growth media (AGM; DMEM, GIBCO, Cat# 41965039; 10% FBS, PAN BioTech, Cat# P30-3306; 10 μM hydrocortisone, Sigma, Cat# H0888; 100 U mL^−1^ penicillin/streptomycin, Sigma, Cat# P4458; 5 μg mL^−1^ insulin, Sigma, Cat# I1882; 1 mM Na pyruvate, GIBCO, Cat# 11360-070; 5 μg mL^−1^ N-acetyl-L-cysteine, Sigma, Cat# A8199) for astrocyte cultures, or in HBSS-HEPES buffer for neuronal cultures. HBSS-HEPES buffer consisted of HBSS 1× without CaCl₂ or MgCl₂ (GIBCO, Cat# 14175-053) supplemented with 10 mM HEPES, pH 7.3 (GIBCO, Cat# 15630-056), and 1% penicillin/streptomycin (GIBCO, Cat# 15140-122).

For astrocyte cultures, cortical tissue was mechanically dissociated by repeated trituration using flame-polished glass pipettes and a 23G needle. The resulting cell suspension was diluted in culture medium and centrifuged at 300*g* for 10 min at room temperature. The pellet was resuspended in AGM and plated at a density of three brains per T75 flask pre-coated with PDL (Sigma, Cat# P6407). Thereafter, cultures were maintained and processed as described for rat primary astrocyte cultures.

For neuronal cultures, cortices were incubated in papain-based enzyme solution (400 U) supplemented with DNase (Sigma, Cat# DN25) for 20 min at 37 °C with occasional mixing. Tissue was subsequently washed three times in plating medium (MEM (1×) with Earle’s salts and L-glutamine, GIBCO, Cat# 31095029; supplemented with 10% heat-inactivated horse serum, GIBCO, Cat# 26050088; 0.6 % glucose, Sigma, Cat# G7021; and 1% penicillin/streptomycin, GIBCO, Cat# 15140-122) and mechanically dissociated by gentle trituration using flame-polished glass pipettes. The resulting cell suspension was collected, and viable cells were counted using a hemocytometer. Subsequent astrocyte–neuron co-culture experiments followed the same experimental paradigm as described for rat primary cultures, with culture conditions adapted for mouse neurons as detailed below.

Neurons were plated at a density of 100,000 cells onto glass coverslips coated overnight with PDL (0.1 mg mL^−1^; GIBCO, Cat# A3890401) and for 2 h with laminin (1 μg mL^−1^; Sigma, Cat# L2020, dissolved in sterile water) prior to plating. Cells were seeded in plating medium. After plating, cells were allowed to settle for 30 min at room temperature in the tissue culture hood, followed by 2 h incubation at 37 °C in plating medium. The plating medium was then replaced with serum-free neuronal medium consisting of Neurobasal (GIBCO, Cat# 21103049) supplemented with 2% B27 (GIBCO, Cat# 17504044), 12 mM glucose (Sigma, Cat# G7021), 0.2% penicillin/streptomycin (GIBCO, Cat# 15140-122), GlutaMAX™ (Thermo Fisher, Cat# 35050038), and 25 μM β-mercaptoethanol (Sigma, Cat# M3148).

Cultures were maintained at 37 °C in a humidified incubator with 5% CO₂ until further use. 48 h after plating, cultures were treated with 5-fluoro-2′-deoxyuridine (FUdR, 5 μM; Sigma, Cat# F0503). After 48 h, the medium was completely replaced with fresh neuronal culture medium.

These protocols are described in more detail on protocols.io and can be accessed at DOI: doi.org/10.17504/protocols.io.3byl4pynrlo5/v1.

### Analysis of shRNA knockdown efficiency in primary astrocytes

#### Lentiviral vector production and primary astrocyte transduction

Lentiviral vectors containing shRNA constructs were generated to induce knockdown of *Atp13a4* in primary astrocyte cultures. HEK293T cells were transfected with a pLKO.1 shRNA Puro plasmid, alongside an envelope plasmid (VSVG) and a packaging plasmid (dR8.91), using X-tremeGENE HP DNA Transfection Reagent (Roche, Cat# 6366236001). 24 h post-transfection, the medium was replaced with AGM. Lentiviral vector-containing media was collected on days 2 and 3 post-transfection. To assess shRNA knockdown efficiency in astrocytes, rat primary astrocytes were plated in 6-well dishes on DIV 7. On DIV 8, 500 μL of the medium was removed, and 500 μL of lentiviral vector-containing media and 1 μg mL^−1^ polybrene were added. On DIV 10 and DIV 12, puromycin (Invivogen, Cat# ant-pr-1) was added at a final concentration of 1 μg mL^−1^ to select for transduced cells. Cells were collected on DIV 15 for RNA extraction and subsequent qPCR analysis to assess knockdown efficiency. The protocol for lentiviral vector production and primary astrocyte transduction is described in more detail on protocols.io: doi.org/10.17504/protocols.io.x54v9rkb1v3e/v1.

#### RNA isolation and cDNA synthesis

Cells were harvested and stored in TRIzol (Invitrogen, Cat# 15596026) at-80 °C until further processing. Before RNA isolation, samples were thawed to room temperature and resuspended in 800 μL of TRIzol. To each sample, 200 μL of chloroform was added and thoroughly mixed. The samples were centrifuged at 12,000*g* for 15 min at 4 °C to separate the phases, and the clear aqueous phase was carefully collected. GlycoBlue Coprecipitant (2 μL of 15 mg mL^−1^; Invitrogen, Cat# AM9515) and 500 μL of isopropanol were then added to each sample, followed by centrifugation at 12,000*g* for 10 min at 4 °C, resulting in the precipitation of RNA as a blue pellet. The RNA pellet was washed with 75% ethanol, centrifuged at 7,500*g* for 5 min at 4 °C, air-dried, and resuspended in 40 μL of nuclease-free water. To remove genomic DNA contamination, samples were treated with DNase I (5 µL Zymosan DNaseI and 5 µL DNA digestion buffer; Zymo Research, Cat# E1010) for 15 min at room temperature. RNA cleanup was then performed using the RNA Clean & Concentrator-5 Kit (Zymo Research, Cat# R1014) according to the manufacturer’s instructions. The concentration of RNA in each sample was determined using a Nanodrop spectrophotometer. To generate cDNA, the RNA samples were incubated with qScript^TM^ cDNA SuperMix (VWR, Cat# 101414-102) and nuclease-free water, using the following conditions: 5 min at 25 °C, 30 min at 42 °C, and 5 min at 85 °C. The resulting cDNA was diluted with 20 μL of nuclease-free water to a final volume of 30 μL and stored at-80 °C until further use. The protocol for RNA isolation and cDNA synthesis is described on protocols.io: doi.org/10.17504/protocols.io.bp2l6dr6rvqe/v1.

#### qPCR

cDNA samples were plated in a 96-well qPCR plate along with Fast SYBR Green Master Mix (Cat# 43-856-16; Applied Biosystems), nuclease-free water, and specific forward and reverse primers. The reaction mix consisted of 5 μL SYBR Green Master Mix, 0.5 μL forward primer (10 μM), 0.5 μL reverse primer (10 μM), and 4 μL cDNA sample (diluted to 12.5 ng μL^-1^ per well). Each sample was plated in two to four technical replicates, and a no-cDNA control (containing all components except cDNA) was included as a negative control. Cycle threshold (Ct) values were obtained for each well and normalized to the housekeeping gene 18S. The sequences of the forward (F) and reverse (R) primers used (5′ → 3′) are as follows: Atp13a4: (F): 5’-GGCAGCCCACCTATACAAACTATATAT-3’ and (R): 5’-GAATGAAAAGACATACGCCCATCT-3’; and 18S: (F) 5′-GCAATTATTCCCCAT GAACG-3′ and (R) 5′-GGCCTCACTAAACCATCCAA-3′. For more details, the complete protocol can be found on protocols.io: doi.org/10.17504/protocols.io.261ger2b7l47/v1.

### Astrocyte morphology analysis

Astrocyte-neuron co-cultures were fixed with 4% PFA (Electron microscopy Sciences, Cat# 19210) at room temperature for 7 min. Following fixation, cells were washed with DPBS (GIBCO, Cat# 14190144) and then blocked for 30 min in antibody blocking buffer containing 50% normal goat serum (NGS; Thermo Fisher, Cat# 01-6201) and 0.4% Triton X-100 Surfact-Amps Detergent Solution (Thermo Fisher, Cat# 28314). After blocking, the cells were incubated overnight at 4 °C with primary antibodies diluted in blocking buffer containing 10% NGS. For rat cultures, astrocytes were labeled with chicken anti-GFP (1:1,000; Aves Labs, Cat# GFP-1020, RRID:AB_10000240) and rabbit anti-GFAP (1:2,000; Agilent, Cat# Z0334, RRID:AB_10013382). For mouse cultures, astrocytes were labeled with chicken anti-GFP (1:1,000; Abcam, Cat# ab13970, RRID:AB_300798) and rabbit anti-GFAP (1:1,000; Abcam, Cat# ab7260, RRID:AB_305808). The following day, samples were washed with DPBS and incubated with Alexa Fluor 488 goat anti-chicken IgY(H+L) (Thermo Fisher, Cat# A-11039, RRID:AB_2534096) and Alexa Fluor 594 goat anti-rabbit IgG(H+L) (Thermo Fisher, Cat# A-11037, RRID:AB_2534095) (diluted 1:1,000) for 2 h at room temperature, followed by additional DPBS washes. After staining, coverslips were mounted onto glass slides using VECTASHIELD Antifade Mounting Medium with DAPI (Vector Laboratories, Cat# H-1200-10) and imaged using a Keyence BZ-X800 microscope at 40× magnification. Astrocyte morphological complexity was evaluated using the Sholl analysis plugin in FIJI (http://fiji.sc,RRID:SCR_002285) (108). Image acquisition was conducted in a blinded manner. For rat cultures, each independent experiment used primary neurons and astrocytes derived from a unique litter of mixed-sex WT rats. For mouse cultures, each independent experiment used WT neurons co-cultured with astrocytes derived from a unique litter of mixed-sex WT or Atp13a4(KO) mice from pure breeding lines. Sholl analysis curves from at least three independent experiments were statistically compared using a mixed-effect model with Tukey post-test, treating variability per experiment as a random effect. The R script used for statistical analysis can be found at: https://github.com/Eroglu-Lab/In-Vitro-Sholl (rat cultures) and https://github.com/emsm3eus-crypto/van-Veen-et-al.-2026/tree/main/sholl%20analysis (mouse cultures). For more details, the complete protocol can be found on protocols.io at doi.org/10.17504/protocols.io.kxygxwobdv8j/v1.

### *In vitro* synapse analysis

On DIV 8 and DIV 11, neurons were treated with ACM (50 μg mL^−1^, as determined by previous studies (44)) or spermidine (10 nM). Immunostaining was conducted similarly to the astrocyte morphology protocol, utilizing the following primary antibodies (diluted 1:500): anti-Bassoon (Cat# ADI-VAM-PS003-F, RRID:AB_11181058; Enzo Life Sciences) and anti-Homer1 (Cat# 160 011, RRID:AB_2120992; Synaptic Systems). Alexa Fluor conjugated secondary antibodies (Alexa Fluor 488 mouse IgG2a and Alexa Fluor 594 mouse IgG1) were used at a 1:1000 dilution. Samples were imaged using a Leica Stellaris 8 confocal microscope with a 63× objective. Excitatory synapses were identified by colocalization of pre-synaptic Bassoon with post-synaptic Homer1 puncta. The number of co-localized synaptic puncta was quantified using SynBot, an ImageJ-based software developed by the Eroglu lab (140). For more details, the complete protocol can be found on protocols.io at doi.org/10.17504/protocols.io.j8nlkobodv5r/v1.

### Neuronal morphology analysis

Cortical neurons were transfected with a GFP plasmid on DIV 6 following AraC treatment. On DIV 8 and DIV 11, cultures were treated with either ACM (100 μg mL^−1^) or spermidine (10 nM). On DIV 13, the neurons were fixed using warm 4% PFA (Electron Microscopy Sciences, Cat# 19210) for 7 min, followed by three DPBS washes to remove residual PFA. For immunostaining, neurons were blocked for 30 min in a solution containing 50% NGS (Thermo Fisher, Cat# 01-6201) and 0.2% Triton X-100 (Thermo Fisher, Cat# 28314). The cells were then incubated overnight at 4 °C with primary anti-GFP antibody (1:1000; Aves Labs, Cat# GFP-1020, RRID:AB_10000240), diluted in blocking buffer containing 10% NGS. After incubation with the primary antibodies, the cultures were washed with DPBS and incubated with Goat anti-Chicken IgY (H+L) Secondary Antibody, Alexa Fluor™ 488 (1:1000; Thermo Fisher, Cat# A-11039, RRID:AB_2534096) in blocking buffer for 3 h at room temperature. Following three additional DPBS washes, the coverslips were mounted onto glass slides with VECTASHIELD Antifade Mounting Medium with DAPI (Vector Laboratories, Cat# H-1200-10). Images were acquired using a Keyence BZ-X800 microscope at 20× magnification, and dendrite length was manually analyzed using the NeuronJ plugin within the FIJI software (http://fiji.sc, RRID:SCR_002285). For more details, the complete protocol can be found on protocols.io at doi.org/10.17504/protocols.io.8epv52bw4v1b/v1.

### Multiplexed immunofluorescence and RNA-FISH for simultaneous detection of protein and RNA targets

Custom-made probes for *Atp13a4* were acquired from Molecular Instruments. Buffers were purchased from Molecular Instruments. Multiplexed immunofluorescence and RNA-FISH protocols were followed as indicated by the manufacturer. Briefly, five 20 μm sections containing the visual cortex from P7, P15 and P21 Aldh1L1-EGFP brains were directly mounted onto glass slides. All sections were kept at-80 °C until use. To remove the cryoprotective solution, the sections were thawed for 5 min at room temperature and washed three times with 1× DPBS containing 0.1% Tween20 (PBS-T). Next, the sections were blocked with antibody buffer at room temperature for 1 h and incubated in a wet chamber with anti-GFP (1:1,000; Aves Labs, Cat# GFP-1020, RRID:AB_10000240) primary antibody overnight at 4 °C. The next day, the sections were washed with PBS-T and incubated at room temperature with 100 μL initiator-labeled secondary antibody diluted in antibody buffer for 1 h. The sections were then washed with PBS-T, incubated with 4% PFA (Electron Microscopy Sciences, Cat# 19210) for 10 min, and immersed in 5× sodium chloride-sodium citrate (C6H7ClNa2O7) containing 0.1% Tween20 (5× SSCT) for 5 min. Next, the sections were incubated with 100 μL of 16 nM of *Atp13a4* probe solution overnight in a 37 °C humidified chamber. The following day, to remove the probe excess, the sections were incubated in a mixture of probe wash buffer and 5× SSCT at 37 °C. Each incubation had an increasing concentration of 5× SSCT, ending with a final 100% 5× SSCT incubation. Next, 200 μL of amplification buffer was added to the sections for 30 min at room temperature. Snap-cooled hairpins specific to the initiator-labeled secondary antibody and *Atp13a4* probe were mixed with the amplification buffers, resulting in a 60 mM hairpin solution. This mixture was added to the sections and incubated at room temperature overnight. Finally, the sections were washed with 5× SSCT for a total of 45 min to remove excess hairpins, then dried and mounted with mounting media. The resulting sections were imaged within two days at high magnification (60× objective with 2× optical zoom) and resolution (1 μm step size, 6 μm z-stack) on an Olympus FV 3000 microscope. For the entire dataset, images of the visual cortex were captured across cortical layers L1, L2/3, L4, and L5, with three sections analyzed per animal. The dataset included samples from three animals per time point: P7 (2 males, 1 female), P15 (3 males), and P21 (2 males, 1 female). The images were then processed using a FIJI custom pipeline that includes thresholding of *Atp13a4* puncta inside and outside the GFP-positive astrocyte area. The full protocol and analysis software are publicly available on protocols.io (DOI: 10.17504/protocols.io.yxmvm9zk9l3p/v1) and GitHub (https://github.com/Eroglu-Lab/Irala_2024_image_analysis/blob/main/20230717_irala_fish_macro.ijm), respectively.

### In vivo TurboID

A detailed version of this protocol is available on protocols.io at doi.org/10.17504/protocols.io.14egn4b9qv5d/v1.

#### Lamp1 plasmids

pZac2.1-GfaABC1D-Lck-GCaMP6f (RRID:Addgene_52924) was a gift from Dr. Baljit Khakh. pZac2.1-GfaABC1D-Lamp1-TurboID-HA was generated by amplifying Lamp1 via PCR from the plasmid pmCherry-Lysosomes-20 (RRID:Addgene_55073) using primers (F): 5’-AGGCTAGCCGCCACCATGGCGGCCCCGGGCGCCCGGCGGC-3’; and (R): 5’-GAGCTCGAGGGTGGCGACCGGTGGATCCGCG-3’). The amplified Lamp1 fragment was then inserted into pZac2.1-GfaABC1D-TurboID using directional cloning.

#### Adeno-associated virus (AAV) production and administration

Purified AAVs were produced as described previously (141). Briefly, HEK293T cells were transfected with pAd-DELTA F6, serotype plasmid AAV PHP.eB, and AAV plasmid (pZac2.1-GfaABC1D-Lamp1-TurboID-HA or pZac2.1-GfaABC1D-NES-TurboID-HA). Three days after transfection, cells were collected in 15 mM NaCl, 5 mM Tris-HCl, pH 8.5, and lysed with repeat freeze-thaw cycles followed by treatment with Benzonase (Novagen, Cat# 70664) at 37 °C for 30 min. Lysed cells were pelleted by centrifugation, and the supernatant, containing AAVs, was applied to an OptiPrep density gradient (15%, 25%, 40%, and 60%; Sigma, Cat# D1556) and centrifuged at 67,000 rpm using a Beckman Ti-70 rotor for 1 h. The AAV-enriched fraction was isolated from between 40% and 60% iodixanol solution and concentrated by repeated washes with sterile PBS in an Amicon Ultra-15 filtration unit (100 kDa MWCO; Millipore, Cat# UFC910008) to a final volume of ∼100 μL and aliquoted for storage at-80 °C. A step-by-step protocol is available at protocols.io: doi.org/10.17504/protocols.io.bp2l623b5gqe/v1.

#### *In vivo* BioID protein purification

*In vivo* BioID experiments were performed as previously described in Takano et al., 2020 with modifications (142). A comprehensive version of this protocol can be found on protocols.io (doi.org/10.17504/protocols.io.14egn4b9qv5d/v1). A timed pregnant CD1 dam was ordered from Charles River and gave birth to a single 16-pup litter. 6 pups were injected with AAVs carrying Astro-Lamp-TurboID (PHP.eB.GfaABC1D-Lamp1-TurboID-HA) or Astro-CYTO-BioID (PHP.eB.GfaABC1D-NES-TurboID-HA). 2 sex-and construct-matched cortices were pooled at the time of protein isolation, yielding 3 independent replicates per TurboID construct. WT CD1 P1 mouse pups were anesthetized by hypothermia (10 min on ice), and 1 µL of each concentrated AAV-TurboID vector was injected bilaterally into the cortex using a Hamilton syringe. Pups were monitored until they recovered on a heating pad. At P18, P19, and P20, biotin was subcutaneously injected at 24 mg/kg to increase the biotinylation efficiency. At P21, mice were anesthetized with 200 mg/kg Tribromoethanol (Avertin) and transcardially perfused with TBS. Cerebral cortices were dissected, frozen in liquid nitrogen, and stored at-80° C. 12 total samples (6 per construct) were subjected to LC-MS/MS and downstream analysis. For the protein purification, each cortex was lysed in a buffer containing 50 mM Tris-HCl, pH 7.5; 150 mM NaCl; 1 mM EDTA; protease inhibitor mixture (Roche); and phosphatase inhibitor mixture (PhosSTOP, Roche). Next, an equal volume of buffer containing 50 mM Tris-HCl, pH 7.5; 150 mM NaCl; 1 mM EDTA; 0.4 % SDS; 2 % TritonX-100; 2 % deoxycholate; protease inhibitor mixture; and phosphatase inhibitor mixture was added to the samples, followed by sonication and centrifugation at 15,000*g* for 10 min. The remaining supernatant was ultracentrifuged at 100,000*g* for 30 min at 4 °C. Finally, after addition of SDS detergent, samples were heated at 45 °C for 45 min. After cooling on ice, each sample was incubated with NeutrAvidin Agarose Resin (Thermo Fisher Scientific, Cat# 29202) at 4 °C overnight. Following incubation, the beads were serially washed: i) twice with a solution containing 2% SDS; ii) twice with a buffer containing 1% TritonX-100, 1% deoxycholate, 25 mM LiCl; iii) twice with 1M NaCl, and finally, five times with 50 mM ammonium bicarbonate. The biotinylated proteins attached to the agarose beads were eluted in a buffer of 125 mM Tris-HCl, pH 6.8; 4% SDS; 0.2% β-mercaptoethanol; 20% glycerol; 3 mM biotin at 60 °C for 15 min.

#### Sample preparation

The Duke Proteomics and Metabolomics Shared Resource (DPMSR) received 6 samples (3 of each Cyto and Lamp1) which were kept at-80 °C until processing. Samples were spiked with undigested bovine casein at a total of either 1 or 2 pmol as an internal quality control standard. Next, samples were reduced for 15 min at 80 °C, alkylated with 20 mM iodoacetamide for 30 min at room temperature, then supplemented with a final concentration of 1.2% phosphoric acid and 588 μL of S-Trap (Protifi) binding buffer (90% MeOH/100mM TEAB) (**Table S2**). Proteins were trapped on the S-Trap micro cartridge, digested using 20 ng/μL sequencing grade trypsin (Promega) for 1 h at 47 °C, and eluted using 50 mM TEAB, followed by 0.2% FA, and lastly using 50% ACN/0.2% FA. All samples were then lyophilized to dryness. Samples were resuspended in 12 μL of 1% TFA/2% acetonitrile with 12.5 fmol/μL of yeast ADH. A study pool QC (SPQC) was created by combining equal volumes of each sample.

#### LC-MS/MS Analysis

Quantitative LC/MS/MS was performed on 3 μL of each sample, using an MClass UPLC system (Waters^TM^) coupled to a Thermo Orbitrap Fusion Lumos high resolution accurate mass tandem mass spectrometer (Thermo) equipped with a FAIMSPro device via a nanoelectrospray ionization source. Briefly, the sample was first trapped on a Symmetry C18 20 mm × 180 μm trapping column (5 μL/min at 99.9/0.1 v/v water/acetonitrile), after which the analytical separation was performed using a 1.8 μm Acquity HSS T3 C18 75 μm × 250 mm column (Waters^TM^) with a 90-min linear gradient of 5 to 30% acetonitrile with 0.1% formic acid at a flow rate of 400 nL/min with a column temperature of 55 °C. Data collection on the Fusion Lumos mass spectrometer was performed for three difference compensation voltages (-40 V,-60 V,-80 V). Within each CV, a data-dependent acquisition (DDA) mode of acquisition with a r=120,000 (@ m/z 200) full MS scan from m/z 375 – 1500 with a target AGC value of 4 × 10^5^ ions was performed. MS/MS scans with HCD settings of 30% were acquired in the linear ion trap in “rapid” mode with a target AGC value of 1 × 10^4^ and max fill time of 35 ms. The total cycle time for each CV was 0.66 s, with full MS scans occurring every 2 s. A 20 s dynamic exclusion was employed to increase depth of coverage. The total analysis cycle time for each sample injection was approximately 2 h.

#### Quantitative data analysis

Following 9 total UPLC-MS/MS analyses (excluding conditioning runs, but including 3 replicate SPQC samples, **Table S3**), data were imported into Proteome Discoverer 3.0 (Thermo Scientific Inc.). Individual LC-MS data files were aligned based on the accurate mass and retention time of detected precursor ions (“features”) using Minora Feature Detector algorithm in Proteome Discoverer. Relative peptide abundance was determined by measuring peak intensities of selected ion chromatograms of the aligned features across all runs. The MS/MS data was searched against the SwissProt *M. musculus* database, a common contaminant/spiked protein database (bovine albumin, bovine casein, yeast ADH, etc.), and an equal number of reversed sequence “decoys” for False Discovery Rate (FDR) determination. Sequest with INFERYS was utilized to produce fragment ion spectra and to perform the database searches. Database search parameters included fixed modification on Cys (carbamidomethyl) and variable modification on Met (oxidation). Search tolerances were 2 ppm precursor and 0.8 Da product ion with full trypsin enzyme rules. Peptide Validator and Protein FDR Validator nodes in Proteome Discoverer were used to annotate the data at a maximum 1% protein FDR based on q-value calculations. Note that peptide homology was addressed using razor rules in which a peptide matched to multiple different proteins was exclusively assigned to the protein with the highest number of identified peptides. Protein homology was addressed by grouping proteins that had the same set of peptides to account for their identification. A master protein within a group was assigned based on % coverage.

Raw intensity values for each identified peptide are presented in **Table S3**. Prior to normalization, a filtering step was applied such that a peptide was removed if it was not detected at least twice across all samples and in at least 50% of replicates within a single experimental group. Following this filtering step, total intensity normalization was performed, where the total intensity of all peptides in each sample was summed and normalized across all samples). Next, the following imputation strategy was applied to handle missing values. If less than 50% of the values were missing in a biological group, missing values were imputed from a normal distribution defined by measured values within the same intensity range (20 bins). If more than 50% of values were missing for a peptide in a group and the peptide intensity was > 5 × 10^6^, the peptide was considered misaligned, and its intensity was set to 0. All remaining missing values were imputed using the lowest 2% of all detected values (**Table S4**). After imputation, peptide intensities were then subjected to a trimmed-mean normalization in which the top and bottom 10% of intensity signals were excluded, and the average of the remaining values was used to normalize across all samples. Lastly, all peptides corresponding to the same protein were summed into a single intensity value (**Table S5**). These normalized protein level intensities were used for all subsequent analyses.

To assess technical reproducibility, we calculated the percentage coefficient of variation (%CV) for each protein across three injections of an SPQC pool that were interspersed throughout the study (**Table S5**). The mean %CV of the SPQC pools was 9.2%, which falls well within expected analytical tolerances. To evaluate both biological and technical variability, we calculated %CVs for each protein across individual experimental groups, yielding an average %CV of 18.9%, indicating reproducible immunoprecipitation and sample processing.

As an initial statistical analysis, we calculated fold-changes in protein abundance between sample groups based on log2-transformed expression values. Two-tailed heteroscedastic *t* tests were performed on the log2-transformed data, and fold changes with corresponding p-values are provided in **Table S6**. To control for multiple testing, FDR correction was applied using the Benjamini-Hochberg procedure, with adjusted p-values reported as “p.adjusted” in **Table S6**. However, to prioritize discovery, we opted to use the less stringent unadjusted p-value for downstream analyses. Within supplemental tables, we have labeled proteins significantly more abundant (“Up”, fold change >2 and unadjusted p-value <0.05), or less abundant (“Down”, fold change >-1.5 and unadjusted p-value <0.05) in a particular genotype or BioID sample group. These annotated proteins were subjected to downstream analyses, including protein interaction networks using Cytoscape (version 3.9.1) and Gene Ontology (GO) enrichment analysis using the ClusterProfiler package for R, with all *M. musculus* genes as the reference background.

## PALE

pPB-shRNA-mCherryCAAX plasmids were synthesized by DNA amplification of hU6 promoter and shRNA from pLKO.1-shRNA-GFP using Phusion High-Fidelity DNA Polymerase (Cat# M0530S; NEB) using primers that also introduced SpeI restriction site ((F): 5′-GGACTAGTCAGGCCCGAAGGAATAGAAG-3′; (R): 5′-GGACTAGTGCCAAAGTGGATCTCTGCTG-3′). After PCR purification and digestion with SpeI, the resulting hU6 and shRNA DNA fragment were ligated into pPB-mCherryCAAX. This protocol is described in more detail on protocols.io and can be accessed at DOI: doi.org/10.17504/protocols.io.4r3l2q34pl1y/v1.

Late P0/early P1 CD1 mouse pups were anesthetized through hypothermia. 1 μL of plasmid DNA (containing 1 μg of pGLAST-PBase and 1 μg of pPB-shRNA-mCherryCAAX, mixed with FastGreen Dye) was injected into the lateral ventricles of one hemisphere using a mouth pipette (Drummond Scientific). After the injection, electrodes were positioned with the positive terminal above the visual cortex and the negative terminal below the chin, and five 50 ms pulses of 100 V, spaced 950 ms apart, were delivered. The pups were then placed on a heating pad to recover from sedation, returned to their home cage for monitoring, and euthanized at P21. Experimental groups were randomly assigned within each litter. Only animals that appeared healthy at the time of collection were included in the analysis. All brains were examined for the presence of electroporated cells through immunohistochemistry, and those lacking successful astrocyte labeling were excluded from the study. For more details, the complete protocol can be found on protocols.io at doi.org/10.17504/protocols.io.n2bvj9z4xlk5/v1.

### Immunohistochemistry

#### PALE experiments

Mice were anesthetized with an i.p. injection of 200 mg/kg tribromoethanol (avertin) and then transcardially perfused with 1X TBS and 4% PFA (Electron Microscopy Sciences, Cat# 19210). Brains were collected and post-fixed overnight in 4% PFA, cryoprotected by immersion in 30% sucrose, frozen in a 2:1 mixture of 30% sucrose and Tissue-Tek Optimal Cutting Temperature (OCT) solution (Electron Microscopy Sciences, Cat# 4583) and stored at-80 °C until sectioning. Coronal brain sections (100 μm thick) were collected and stored in a 1:1 solution of TBS and glycerol at-20 °C.

For immunohistochemical staining to analyze PALE knockdown astrocytes, 100 μm free-floating sections were first washed and permeabilized in 1× TBS containing 0.2% Triton X-100 (0.2% TBST), then blocked in 10% NGS (Thermo Fisher, Cat# 01-6201) diluted in 0.2% TBST. Sections were incubated with rabbit anti-RFP primary antibody (1:1,000; Rockland, Cat# 600-401-379, RRID:AB_2209751) at 4 °C for three nights. After primary antibody incubation, sections were washed in 0.2% TBST and incubated with Alexa Fluor 594 goat anti-rabbit IgG(H+L) secondary antibodies (1:200; Thermo Fisher, Cat# A-11037, RRID:AB_2534095) for 3 h at room temperature. Sections were then washed with 0.2% TBST. For nuclear staining, DAPI (Invitrogen, Cat# D1306) was added at a 1:50,000 concentration before the final wash. Finally, sections were mounted onto glass slides using a homemade mounting medium (90% glycerol, 20 mM Tris pH 8.0, 0.5% n-propyl gallate). Whole astrocytes in the primary visual cortex were imaged at high magnification (60× objective, 2× zoom) and resolution (0.5 μm step size) on an Olympus FV 3000 microscope. The researcher acquiring images was blinded to the experimental group. Imaged astrocytes were then reconstructed in 3D using a filament tracer on Imaris Bitplane 9.9, and “Convex Hull” was used to create a surface that represented the territory of the astrocyte (106). Astrocyte territory volume across experimental conditions was analyzed using nested *t* test. The exact number of astrocytes and animals analyzed in each condition is specified in the figure legend. For more details, the complete protocol can be found on protocols.io at doi.org/10.17504/protocols.io.n2bvj9z4xlk5/v1.

#### Synapse experiments

Mice were euthanized with an overdose of sodium pentobarbital (400 mg/kg, i.p., Dolethal), and transcardially perfused with saline and 4% PFA in DPBS, as described at doi.org/10.17504/protocols.io.5jyl8p3qrg2w/v1. After post-fixation overnight in 4% PFA, 40 µm thick sagittal brain sections were made with a vibrating microtome (HM 650 V, Microm). This protocol is described in more detail on protocols.io and can be accessed at doi.org/10.17504/protocols.io.j8nlko72xv5r/v1.

For immunohistochemical staining to analyze excitatory synapse numbers, 40 μm free-floating sections were first washed and permeabilized in 0.2% TBST, then blocked in 10% NGS (Dako, Cat# X0907) diluted in 0.2% TBST. Sections were incubated with rabbit anti-PSD95 (1:300; Thermo Fisher, Cat# 51-6900, RRID:AB_2533914) and guineapig anti-Bassoon (1:500; Synaptic Systems, Cat# 141 318, RRID:AB_2927388) primary antibodies at 4 °C for two nights. After primary antibody incubation, sections were washed in 0.2% TBST and incubated with Alexa Fluor 488 goat anti-rabbit IgG(H+L) (1:100; Thermo Fisher, Cat# A-11008, RRID:AB_143165) and Alexa Fluor 647 goat anti-guineapig IgG(H+L) secondary antibodies (1:100; Thermo Fisher, Cat# A-21450, RRID:AB_2535867) for 3 h at room temperature. Sections were then washed with 0.2% TBST. For nuclear staining, DAPI (Thermo Fisher, Cat# 62248) was added at a 1:4000 concentration before the final wash. Finally, sections were mounted onto glass slides using Mowiol (Calbiochem, Cat# 475904). High magnification 63× objective plus 1.6× optical zoom z-stack images containing 3 optical sections spaced 0.34 μm apart were acquired using the Zeiss LSM 880 microscope (Cell and Tissue Imaging Cluster (CIC), supported by Hercules AKUL/15/37_GOH1816N and FWO G.0929.15 to Pieter Vanden Berghe, University of Leuven). Excitatory synapses were identified by the colocalization of pre-(Bassoon) and post-synaptic (PSD95) puncta. The number of co-localized synaptic puncta were obtained using SynBot, an ImageJ-based software developed by the Eroglu lab (140). The number of animals analyzed in each condition is specified in the figure legend. This protocol is described in more detail on protocols.io and can be accessed at doi.org/10.17504/protocols.io.3byl4wxmzvo5/v1.

#### DAB-based Immunohistochemistry for Iba1 and Gfap

Free-floating sections used for chromogenic immunohistochemistry were prepared as described for synapse experiments. Sections were first rinsed in PBS (Gibco, Cat# 21600-096) and subjected to heat-induced antigen retrieval by incubation in citrate buffer (pH 6.0) at 80 °C for 30 min, followed by cooling on ice for 20 min in the same buffer and subsequent PBS washes, including a 5 min wash under agitation. Endogenous peroxidase activity was quenched by incubation in PBS containing 3% hydrogen peroxide (Chemlab, Cat# CL00.2306.1000) and 10% methanol (VWR, Cat# 20846.361) for 10 min at room temperature, followed by washes in PBS containing 0.1% Tergitol (Thermo Scientific, Cat# 464252500). Sections were then incubated overnight at room temperature with primary antibodies diluted in PBS supplemented with 10% serum (Iba1, 1:500; Abcam, Cat# ab5076, RRID:AB_2224402; GFAP, 1:2000; Agilent, Cat# Z0334, RRID:AB_10013382). After washing in PBS-T (0.1% Tergitol), sections were incubated with biotinylated secondary antibodies (1:600; rabbit anti-goat, Dako, Cat# E0466; goat anti-rabbit, Abcam, Cat# ab6720, RRID:AB_954902) for 30 min at room temperature, followed by further PBS-T washes. Sections were subsequently incubated with streptavidin-HRP (1:1000; Invitrogen, Cat# S911, RRID:AB_3741654) for 30 min at room temperature and washed again in PBS-T. For signal development, 3,3′-diaminobenzidine (DAB; Sigma-Aldrich, Cat# D5905) was dissolved in PBS (pH 7.6), filtered (0.22 µm), and supplemented with hydrogen peroxide immediately prior to use. Sections were incubated in DAB solution (∼250 µl per well) for 3-6 min at room temperature (GFAP ∼3 min; Iba1∼6 min), and the reaction was stopped by PBS-T washes. Sections were transferred to fresh PBS, briefly rinsed in a 1:1 mixture of PBS and distilled water, and mounted onto gelatin-coated slides. After overnight drying, sections were dehydrated through graded ethanol (70%, 90%, 100%, 100%), cleared in Histoclear II (Agar Scientific, Cat# A2-0105), and mounted with DPX mounting medium (Sigma-Aldrich, Cat# 06522). Slides were imaged using a Leica Aperio CS2 slide scanner at 20× magnification, and image quantification was performed using FIJI (ImageJ, http://fiji.sc, RRID:SCR_002285). This protocol is described in more detail on protocols.io: doi.org/10.17504/protocols.io.q26g7oj83vwz/v1.

### Validation of the *Atp13a4* knockout mouse model

To confirm successful generation of the *Atp13a4* KO mouse model, validation was performed at both the genomic and transcriptional level. gDNA was extracted from ear notches using standard lysis buffer. Genotyping PCR was conducted using two primer sets: *Atp(50715)CF* (5′-GGTCCCAGAATTCCTTGGCA-3′) and *Atp(50715)WTR* (5′-CAGTGCTGACAGGGAGATCC-3′) for WT amplification, and *Atp(50715)CF* and *Atp(50715)KOR* (5′-GCCTCACACTGCACTCTTCT-3′) for KO amplification. The PCR cycling conditions included an initial denaturation at 95 °C for 2 min, followed by 30 cycles of 95 °C for 15 s, 56 °C for 15 s, and 72 °C for 30 s, with a final extension at 72 °C for 7 min. Amplified products were visualized on a 1.5% agarose gel stained with Midori Green (1:10,000), with WT alleles producing a 410 bp band and KO alleles producing a 288 bp band. This protocol is described in more detail on protocols.io and can be accessed at doi.org/10.17504/protocols.io.kqdg3q8kev25/v1.

Transcriptional validation was performed by RT-PCR and Sanger sequencing of cortical RNA. RNA was isolated from cortical brain extracts of WT and KO mice using the NucleoSpin RNA Plus Kit (Macherey-Nagel, Cat# 740984.250), and 5 µg was converted to cDNA in 50 µl using the High-Capacity cDNA Reverse Transcription Kit (Applied Biosystems, Cat# 4374966). 100 ng of cDNA was used as template for PCR amplification using iProof™ High-Fidelity DNA Polymerase (Bio-Rad, Cat# 1725302) and the primer pair msAtp13a4_EXON1-7: forward 5′-ACCTTGAGAAGAGCCAGCAT-3′ and reverse 5′-GAGGTCATACACGGTCAAAGC-3′. Cycling conditions were as follows: initial denaturation at 98 °C for 3 min, followed by 35 cycles of 98 °C for 30 s, 56 °C for 30 s, and 72 °C for 30 s, with a final extension at 72 °C for 10 min. PCR products were resolved by agarose gel electrophoresis, after which bands of interest were excised and purified using the GeneJET Gel Extraction Kit (Thermo Scientific, Cat# K0692). Purified fragments were submitted for Sanger sequencing at Eurofins Genomics using the same primers as described above.

To quantify the reduction in Atp13a4 expression at the transcript level, RT-qPCR was performed targeting exon 6. RNA isolation and cDNA synthesis were carried out as described above. SYBR Green-based RT-qPCR was performed on a LightCycler (Roche) under the conditions described in the RT-qPCR section, using primers targeting exon 6 of msAtp13a4 (forward 5′-GAGGTTAATATGTGGGCCTA-3′, reverse 5′-ACCTCCTTGATGAGCAGTTTC-3′). GAPDH served as the normalization reference (msGapdh: forward 5′-TGTGTCCGTCGTGGATCTGA-3′, reverse 5′-CCTGCTTCACCACCTTCTTGA-3′).

### Whole cell electrophysiology

For whole-cell patch clamp recordings, acute slices were prepared from 8 weeks old WT and Atp13a4(KO) mice. In short, animals were sedated using isoflurane and after decapitation, the brain was quickly removed and transferred into ice-cold cutting solution: 110 mM choline chloride, 26 mM NaHCO3, 11.6 mM Na-ascorbate, 10 mM D-glucose, 7 mM MgCl2, 3.1 mM Na-pyruvate, 2.5 mM KCl, 1.25 mM NaH2PO4, 0.5 mM CaCl2; 300–315 mOsm, pH adjusted to 7.4, with 5% CO2/95% O2. Coronal slices (250 µm) containing primary visual cortex were made using a vibratome (Leica VT1200). Immediately after cutting, the slices were transferred to 32 °C cutting solution for 6 min to recover and afterwards were stored at room temperature in holding solution: 126 mM NaCl, 26 mM NaHCO3, 10 mM D-glucose, 6 mM MgSO4, 3 mM KCl, 1 mM CaCl2, 1 mM NaH2PO4, 295-305 mOsm, pH adjusted to 7.4, with 5% CO2/95% O2. Slices were stored for ∼1 h before experiments. During experiments brain slices were continuously perfused in a submerged chamber (Warner Instruments) at a rate of 3-4 mL/min with recording solution: 127 mM NaCl, 2.5 mM KCl, 1.25 mM NaH2PO4, 25 mM NaHCO3, 1 mM MgCl2, 2 mM CaCl2, 25 mM D-glucose (pH 7.4 with 5% CO2/95% O2), including AP-5 (100 µM) and TTX (1 µM). All recordings were done between 31-33 °C. For recordings we used borosilicate glass recording pipettes (resistance 3.5–5 MΩ, Sutter P-1000) filled with the following internal solution: 126 mM CsMSF, 10 mM HEPES, 2.5 mM MgCl2, 4 mM ATP, 0.4 mM GTP, 10 mM Creatine Phosphate, 0.6 mM EGTA, 5 mM QX-314 and 3 mg mL^−1^ biocytin. Whole-cell patch-clamp recordings were done using a double EPC-10 amplifier under control of Patchmaster v2 x 32 software (HEKA Elektronik, Lambrecht/Pfalz, Germany). Currents were recorded at 20 Hz and low-pass filtered at 3 kHz when stored. Series resistance was compensated to 70-80% and monitored throughout the experiments. We recorded L2/3 pyramidal in the primary visual cortex (V1). For each cell both miniature excitatory synaptic inputs (mEPSCs; Vm=-90 mV) and miniature inhibitory synaptic inputs (mIPSCs; Vm= 0 mV) were recorded. Miniature input was quantified using the Mini Analysis program (Synaptosoft). This protocol is described in more detail on protocols.io and can be accessed at doi.org/10.17504/protocols.io.3byl4ww52vo5/v1.

### Mouse tissue collection

#### Plasma

Facial vein blood samples of two-month-old mice were collected in EDTA coated tubes (Sarstedt, Cat# 20.1288) using an animal lancet (Goldenrod). Samples were centrifuged at 2,000*g* for 15 min at 4 °C and plasma was stored at-80 °C in protein low binding tubes (Thermo Fisher Scientific, Cat# 90410). A detailed protocol can be found on protocols.io: doi.org/10.17504/protocols.io.8epv5yq7jl1b/v1.

#### Brain tissue collection

Following euthanasia by cervical dislocation, the head was removed and a midline incision was made to expose the skull. The skull was carefully opened by making a caudal incision along the interparietal bone, followed by a frontal cut between the eyes to facilitate removal. The parietal bones were gently peeled away using fine forceps, taking care not to damage the underlying meninges or brain tissue. The brain was then carefully lifted from the skull by sliding forceps underneath the olfactory bulbs and severing cranial nerve connections. Immediately after extraction, the brain was rinsed in ice-cold DPBS to remove residual blood. For metabolomic analyses, the cortex was dissected on an ice-cold surface, rapidly isolated, and transferred into pre-labeled low-binding tubes. For immunoblotting experiments, whole brains were collected and processed as homogenates. All samples were snap-frozen in liquid nitrogen and stored at-80 °C until further processing. A detailed protocol can be found on protocols.io: doi.org/10.17504/protocols.io.yxmvm8jmng3p/v1.

#### CSF

CSF was collected from the cisterna magna of anesthetized mice using a glass capillary-based approach, as previously described (143), with minor modifications. Briefly, mice were anesthetized and positioned in a stereotaxic frame with the head flexed to expose the cisterna magna. Following surgical exposure of the dura, a pulled glass capillary was inserted into the cisterna magna, and CSF was collected by gentle negative pressure while avoiding blood contamination. Collected CSF was transferred into protein low-binding tubes, centrifuged at 10,000*g* for 10 min at 4 °C to remove debris, snap-frozen on dry ice, and stored at-80 °C until further analysis. This protocol is described in more detail on protocols.io: doi.org/10.17504/protocols.io.81wgbj8k1vpk/v1.

### Mouse behavior analysis

#### Developmental milestone assessment

Developmental milestones were assessed using a well-established protocol (144). Experimental cohorts were generated from heterozygous breedings, and *Atp13a4* WT and KO littermates were analyzed within the same litters to control for maternal and environmental effects. On the day of birth (P0), each pup was weighed and genotyped. From P1 onwards, pups were evaluated daily for the acquisition of developmental milestones and sensorimotor reflexes. The test battery included surface righting, negative geotaxis, cliff aversion, rooting, forelimb grasp, auditory startle, ear twitch reflex, open field traversal, eye opening, and air righting. During testing, pups were maintained under a heating lamp (60 W) to preserve physiological body temperature. All assessments were conducted at a consistent time each day to control for circadian influences. Tests were repeated daily until the predefined acquisition criterion was met on two consecutive days, and the age of acquisition was recorded accordingly. Acquisition criteria were defined as follows:

- Surface righting: pup placed supine rights itself onto its abdomen with all four paws contacting the surface. Latency to righting was measured, with a maximum cut-off of 30 s. Acquisition was defined as successful righting within 30 s for two consecutive days.
- Negative geotaxis: pup placed head-down on a 45° inclined mesh surface rotates 180° to a head-up position. Latency was measured with a 30 s cut-off. Acquisition was defined as completion within 30 s for two consecutive days.
- Cliff aversion: pup positioned with forepaws and snout over the edge of a platform withdraws and turns away from the edge. Latency was measured with a 30 s cut-
- off. Acquisition was defined as successful withdrawal within 30 s for two consecutive days.
- Rooting reflex: gentle stroking of the snout/head with a fine filament (three strokes per side) elicits a directed head turn toward the stimulus. Acquisition was defined as a clear response for two consecutive days.
- Forelimb grasp: pup grasps a horizontal rod and maintains grip. Latency to release was recorded. Acquisition was defined as maintaining grip for ≥1 s for two consecutive days.
- Auditory startle: pup exhibits a whole-body startle response to a sharp auditory stimulus (hand clap at ∼10 cm distance). Acquisition was defined as a consistent startle response for two consecutive days.
- Ear twitch reflex: light tactile stimulation of the ear induces ear flattening or twitching. Acquisition was defined as a reproducible response for two consecutive days.
- Open field traversal: pup placed in the center of a 13 cm diameter circle moves completely outside the circle. Latency was recorded with a 30 s cut-off. Acquisition was defined as exiting the circle within 30 s for two consecutive days.
- Eye opening: assessed daily and scored as 0 (no eyes open), 1 (one eye open), or 2 (both eyes open).
- Air righting: pup dropped from a supine position (∼10–12 cm) rights itself mid-air and lands on all four paws. Acquisition was defined as correct landing for two consecutive days.

To account for litter effects, a nested *t* test was used for statistical comparisons, with individual pup values nested within litters for WT and KO groups. These protocols are described in more detail on protocols.io: doi.org/10.17504/protocols.io.rm7vz4d1rlx1/v1.

### Adult behavioral assessment

Adult behavioral testing was performed in two-month-old WT and KO littermates.

### 24-h cage activity

Cage activity was recorded using a lab-built activity logger connected to three IR photo beams. Mice were placed individually in 20 x 30 cm^2^ transparent cages located between the photo beams. Over a period of 24 h, activity was measured and expressed as number of beam crossings. Counts were reset every 30 min. Recording began around noon, with lights automatically switching off at 19:00 and on again at 07:00. This protocol is described in more detail on protocols.io: doi.org/10.17504/protocols.io.j8nlkzj21l5r/v1.

#### Open field exploration

Open field exploration was examined using a 50 × 50 cm^2^ square arena. Animals were dark adapted for 30 min and placed in the arena for 10 min each. Movements of the mice in the arena were recorded using ANY-maze video tracking equipment and software (Stoelting Europe, Ireland). Total path length and corner crossings were included as measures of locomotor activity. These measures are highly correlated and mainly indicate thigmotactic walking near the walls of the arena. Entries into the center of the field were recorded as a measure of conflict resolution or anxiolysis. This protocol is described in more detail on protocols.io: doi.org/10.17504/protocols.io.rm7vz4d2rlx1/v1.

#### Morris water maze

Spatial memory abilities were examined in the standard hidden-platform acquisition and retention (i.e. long-term memory), and working memory versions of the Morris water maze (145). A 150-cm circular pool was filled with water, opacified with non-toxic opaciefier (ACUSOL™ OP 301 Opacifier, Dow Chemicals), and kept at 26 °C as previously described (146). A 15-cm round platform was hidden 1 cm beneath the surface of the water at a fixed position. Each daily trial block consisted of 4 swimming trials (15-30 min intertrial interval) starting randomly from each of 4 starting positions. Mice that failed to find the platform within 100 s were guided to the platform, where they remained for 15 s before being returned to their cages. Interspersed probe trials were conducted at the end of a working week. During these probe trials, the platform was removed from the pool, and the search pattern of the mice was recorded for 100 s. Swimming paths of the animals were recorded using EthoVision video tracking equipment and software (Noldus bv, Wageningen, The Netherlands). This protocol is described in more detail on protocols.io: doi.org/10.17504/protocols.io.5jyl84zk7g2w/v1.

#### Rotarod

Motor coordination and equilibrium were tested on an accelerating rotarod (Ugo Basile, Italy). Mice were first trained at constant speed (5 rpm, 2 min) before starting with four test trials (intertrial interval, 10 min). During these test trials, the animals had to balance on a rotating rod that accelerated from 5 to 40 rpm in 5 min, and time until they dropped from the rod was recorded (up to the 5-min cut-off). 4 test trials were averaged and expressed as mean latency ± SEM for analysis. This protocol is described in more detail on protocols.io: doi.org/10.17504/protocols.io.261geybjyv47/v1.

#### Quantitative gait analysis

Gait analysis was performed using the DigiGait Imaging System (Mouse Specifics Inc., Framingham, Massachusetts, USA). Mice were placed on a transparent treadmill belt moving at 15 cm/s, and ventral-plane video was captured at high speed once steady-state locomotion was achieved. Gait parameters (including stride length, stride frequency, stance width, swing time, and stance time) were automatically computed using DigiGait Analyzer software (version 16A). This protocol is described in more detail on protocols.io: doi.org/10.17504/protocols.io.n92ld4xn9l5b/v1.

#### Three-chamber social preference test

Social behavior was assessed in a three-chamber apparatus made of transparent plexiglass. The setup consisted of a central chamber containing the test mouse (40 cm × 10 cm) connected on both sides with small side chambers (10 cm × 10 cm) that could each hold a stranger mouse. The walls separating the side chambers from the central chamber were perforated (8 mm holes) to allow sniffing while limiting direct physical contact. The setup was placed in an enclosure with dim lightning, and exploration was recorded with an overhead webcam connected to a computer running ANY-maze tracking software (Stoelting Europe, Ireland). The protocol consisted of three consecutive trials. First, during a 5 min habituation trial, the test mouse was placed in the central chamber and locomotion was recorded. This was followed by a social preference trial, where one stranger was placed either in right or left side-chamber (position was randomized across all animals to prevent side bias) and approach behavior was recorded for 10 min. After 10 min, a second stranger mouse was placed in the previously empty side chamber, and approach behavior was recorded again. Stranger mice were sex-and age-matched and originated from different cages. After testing, animals were placed back in their home cage, and the setup was cleaned with 50% ethanol to remove any odor traces. Four animals were tested in parallel. This protocol is described in more detail on protocols.io: doi.org/10.17504/protocols.io.6qpvrbnqolmk/v1.

#### PTZ challenge

Seizures were recorded in a small box (15 × 15 cm and 30 cm high) in front of a camera and tracking software (EthoVision XT, Noldus Information Technology). To induce seizures pharmacologically, PTZ was dissolved in saline (2.5 mg/ml) and injected i.p. at 1% body weight. After injection, animals were immediately placed in the box and behavior was video recorded for 15 min. Occurrence and onset of behavior such as supine position (laying flat on floor), head jerks, Straub tail, clonic and tonic seizures were scored (147). This protocol is described in more detail on protocols.io: doi.org/10.17504/protocols.io.q26g7ok6qvwz/v1.

Behavioral data were evaluated using the appropriate multiplicity-adjusted tests for each assay. As the battery encompasses independent behavioral domains (e.g. anxiety, spatial learning, motor coordination), corrections were applied within, rather than across, tests to maintain sensitivity while adhering to reduction principles. Importantly, the female-specific pattern was reproducible across multiple independent assays, indicating that the observed effects do not reflect false positives arising from multiple testing.

### Quantification and statistical analysis

All statistical analyses were conducted using GraphPad Prism (versions 9 or 10), except for astrocyte Sholl analysis, body weight, and eye opening, which were analyzed in R using custom code. Exact value of *n*, what *n* represents, and the statistical tests used for each experiment are provided in the figure legends. Data are represented as mean ± standard error of the mean, with individual data points displayed where applicable. Exact P-values are reported in the figures. Sample sizes were chosen based on prior experience with each assay to ensure sufficient power to detect meaningful effects; no formal statistical methods were used to predetermine the sample size.

## Resource availability

Further information and requests for resources and reagents should be directed to and will be fulfilled by the lead contact, Sarah van Veen.

## Materials availability

This study did not generate new unique reagents.

## Data and code availability

All datasets generated or analyzed in this study can be found through the Zenodo repository at DOI: https://doi.org/10.5281/zenodo.14719448 (Fig. 1-6) and DOI: https://doi.org/10.5281/zenodo.14717838 (Supplementary Fig. 1-16). The mass spectrometry proteomics data have been deposited to the ProteomeXchange Consortium via the PRIDE (148) partner repository with the dataset identifier PXD063347. All datasets will be made publicly available as of the date of publication. All original code has been deposited at the Eroglu lab GitHub site (https://github.com/Eroglu-Lab/) and at https://github.com/emsm3eus-crypto/van-Veen-et-al.-2026. All experimental protocols are shared via protocols.io (doi.org/10.17504/protocols.io.4r3l29p3xv1y/v1). Respective DOIs are listed in the Key Resources Table. For the purpose of open access, the author has applied a CC BY 4.0 public copyright license to all Author Accepted Manuscripts arising from this submission.

## Supporting information

Key Resource Table

## Acknowledgments

This work was funded by the Fonds voor Wetenschappelijk Onderzoek (FWO, Research Foundation Flanders) (G094219N to P.V., G011424N to P.V. and S.v.V.), the Queen Elisabeth Medical Foundation for Neurosciences (to S.v.V.) and Aligning Science Across Parkinson’s (ASAP-000458 to P.V. and V.B.; and ASAP-020607 to C.E.) through the Michael J. Fox Foundation for Parkinson’s Research (MJFF). S.v.V. was supported by postdoctoral fellowships from the FWO (1253721N) and KU Leuven (PDMt1/24/010). Cartoon elements of figure panels were created using BioRender.com. We thank Anna Vercauteren for assistance with mouse genotyping. We also acknowledge our frequent use of the facilities and equipment of the Leuven Viral Vector Core facility (KU Leuven), Metabolomics Expertise Center (VIB-KU Leuven), Cell and Tissue Imaging Cluster (supported by Hercules AKUL/15/37_GOH1816N and FWO G.0929.15 to P. Vanden Berghe, KU Leuven), the KU Leuven FACS Core, and the mouse behavioral phenotyping core mINT (Z. Callaerts-Vegh, KU Leuven). S.v.V. thanks S. Martin for his continuous support and insightful discussions.

## Author contributions

The study was designed by S.v.V., C.E. and P.V., with M.G.H. and J.E. providing critical input and discussions. E.M. performed bioinformatic analyses of human and mouse expression datasets, contributed to developmental milestone experiments and performed the corresponding data analysis, and conducted Sholl analysis of mouse astrocytes.

C.V.d.H. and J.A. generated all stable cell lines, analyzed ATP13A4 knockdown, and performed RT-PCR analysis of cortical RNA samples. G.S. and D.S.B. conducted and analyzed the TurboID experiments. D.I. and J.S. performed and analyzed the FISH experiments, and D.I. performed ACM collections. K.S. performed statistical analysis of the primary rat astrocyte morphology experiments and conducted spermidine treatment experiments in neurons, including imaging. Z.L. analyzed dendritic morphology following spermidine treatment. K.W. performed and analyzed the electrophysiology experiments.

E.A. contributed to BODIPY-polyamine uptake experiments and western blot analysis of stable cell lines. H.D. and N.S. performed and analyzed mouse genotyping and provided brain sections for synapse analysis; H.D. additionally prepared CSF, plasma, and cortex samples for metabolomics and performed DAB staining and imaging. H.E.G. contributed to CSF sample collection and provided valuable input on statistical analysis. M.M.G.M. performed mouse neuron isolation. K.P., K.R. and K.L.Y. provided patient data; S.v.V conducted and analyzed all other experiments; S.v.V., V.B., P.V. and C.E. secured funding; and S.v.V. and P.V. wrote the manuscript, which was reviewed and approved by all authors.

## Supplementary Information

**Key Resources Table.** Detailed list of protocols, datasets, code/software, reagents, genetically modified mouse strains and cell lines used in this study.

**Table S1.** Oligonucleotide list.

**Table S2.** Sample information and experimental conditions for TurboID study. **Table S3.** Raw peptide intensity data before filtering and normalization for TurboID. **Table S4.** Peptide intensity data after imputation of missing values for TurboID. **Table S5.** Normalized peptide intensity data for TurboID.

**Table S6.** Differentially abundant proteins between experimental groups.

**Supplementary Data.** Uncropped blots and autoradiograms.

---

**Supplementary Fig. 1:**
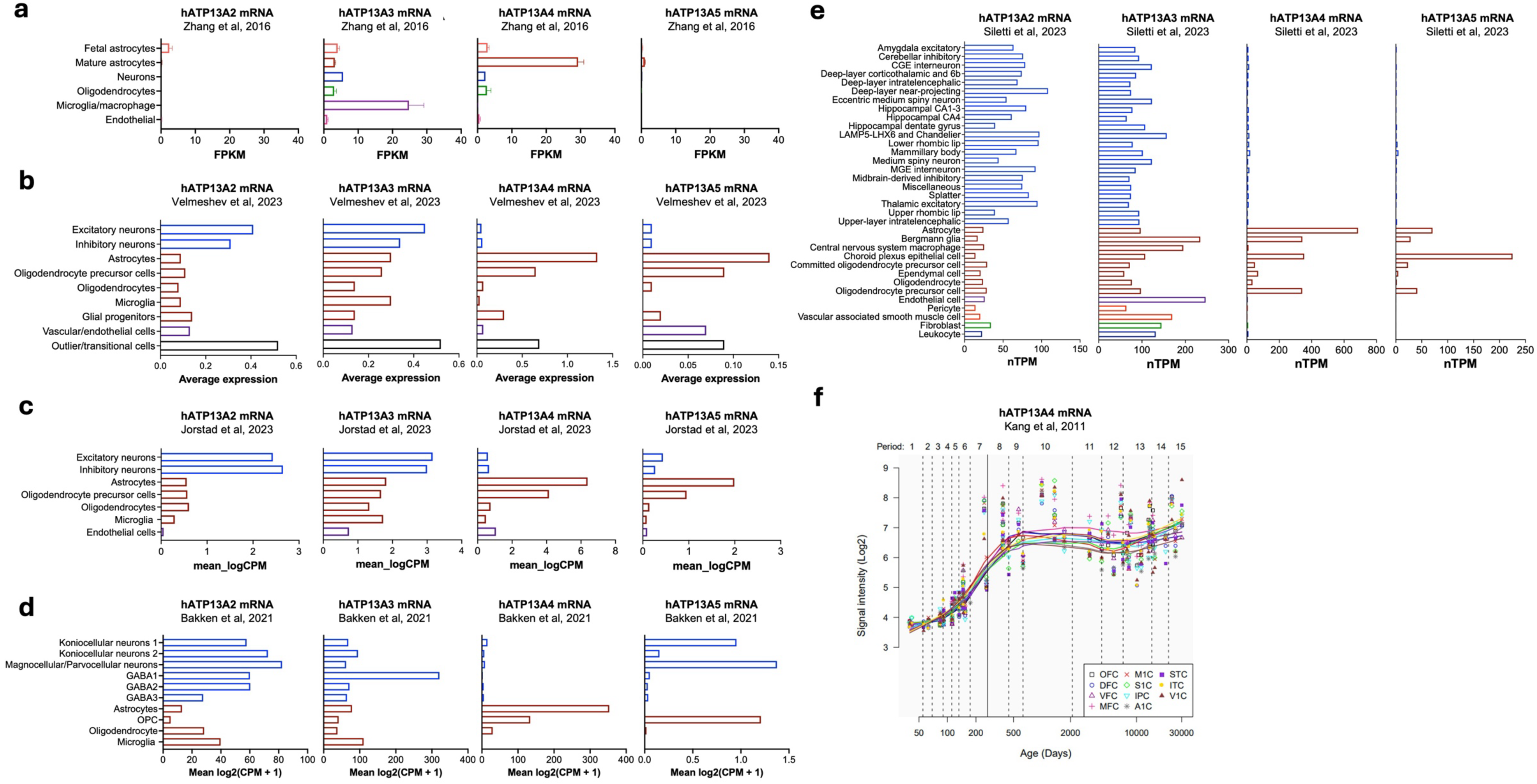
hATP13A4 is enriched in astrocytes and oligodendrocyte precursor cells in the human brain. Analysis of *hATP13A2-5* mRNA expression across major human brain cell types using publicly available transcriptomic datasets. **a**, Bulk RNA-seq from purified human brain cell populations (80), shown as FPKM (Fragments Per Kilobase Million), demonstrating strong enrichment of *hATP13A4* in astrocytes, with higher expression in mature compared to fetal astrocytes; **b-e**, Single-cell and single-nucleus RNA-seq datasets from human prenatal and postnatal cortex (82), neocortex (83), dorsal lateral geniculate nucleus (84), and adult human brain (85), shown as CPM (counts per million) or nTPM (normalized transcript-per-million) as indicated. Across datasets, *hATP13A4* expression is predominant in astrocytes, with lower expression in oligodendrocyte precursor cells (OPCs) and additional enrichment in specialized glial populations such as Bergmann glia and choroid plexus epithelial cells, while neuronal and microglial expression remains low to undetectable. **f**, *hATP13A4* expression across human cortical regions (81) (Human Brain Transcriptome Atlas; http://hbatlas.org/), showing increased expression during late fetal and early postnatal development, consistent with potential developmental regulation. OFC, orbital prefrontal cortex; DFC, dorsolateral prefrontal cortex; VFC, ventrolateral prefrontal cortex; MFC, medial prefrontal cortex; M1C, primary motor cortex; S1C, primary somatosensory cortex; IPC, posterior inferior parietal cortex; A1C, primary auditory cortex; STC, posterior superior temporal cortex; ITC, inferior temporal cortex; V1C, primary visual cortex.

**Supplementary Fig. 2:**
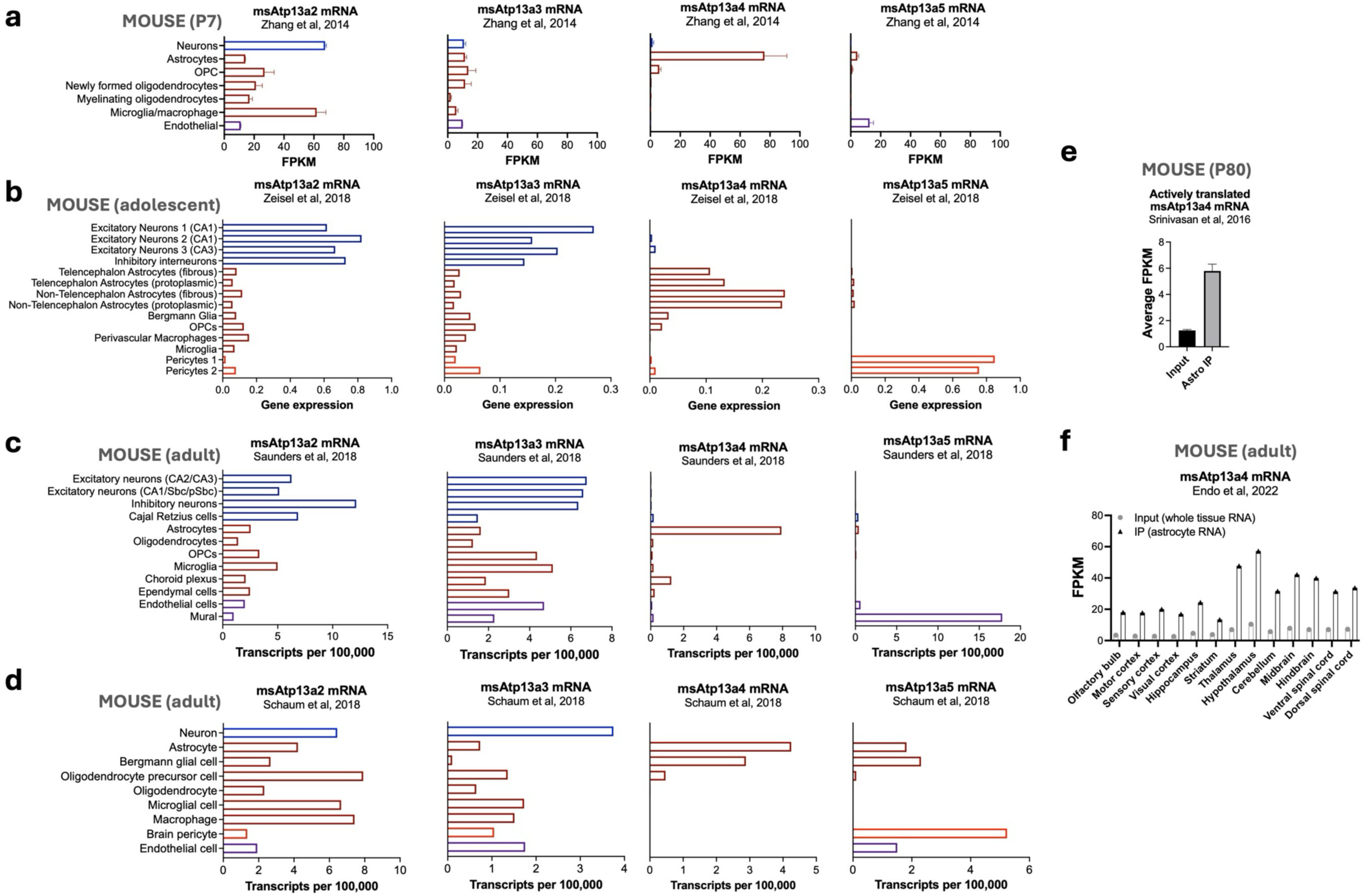
msAtp13a4 is predominantly expressed in astrocytes in the mouse brain. Analysis of *msAtp13a2-5* mRNA levels across major mouse brain cell types and developmental stages using publicly available transcriptomic datasets. **a-d**, Bulk and single-cell RNA-seq datasets from mouse cortex and brain, including purified cortical cell populations at postnatal day 7 (P7) (86), adolescent brain (87), adult brain (88), and the Tabula Muris dataset (89), demonstrating strong and selective enrichment of *msAtp13a4* in astrocytes, with negligible expression in neurons and microglia and minimal expression in oligodendroglial lineages. **e**, Ribosome profiling (Astro-IP) showing active translation of *msAtp13a4* mRNA in P80 mouse astrocytes compared to bulk tissue RNA (input) (90). **f**, Region-specific astrocyte RNA-seq datasets showing consistent enrichment of *msAtp13a4* across 13 CNS regions in adult mice (91), identifying msAtp13a4 as part of a core astrocyte-enriched gene set.

**Supplementary Fig. 3:**
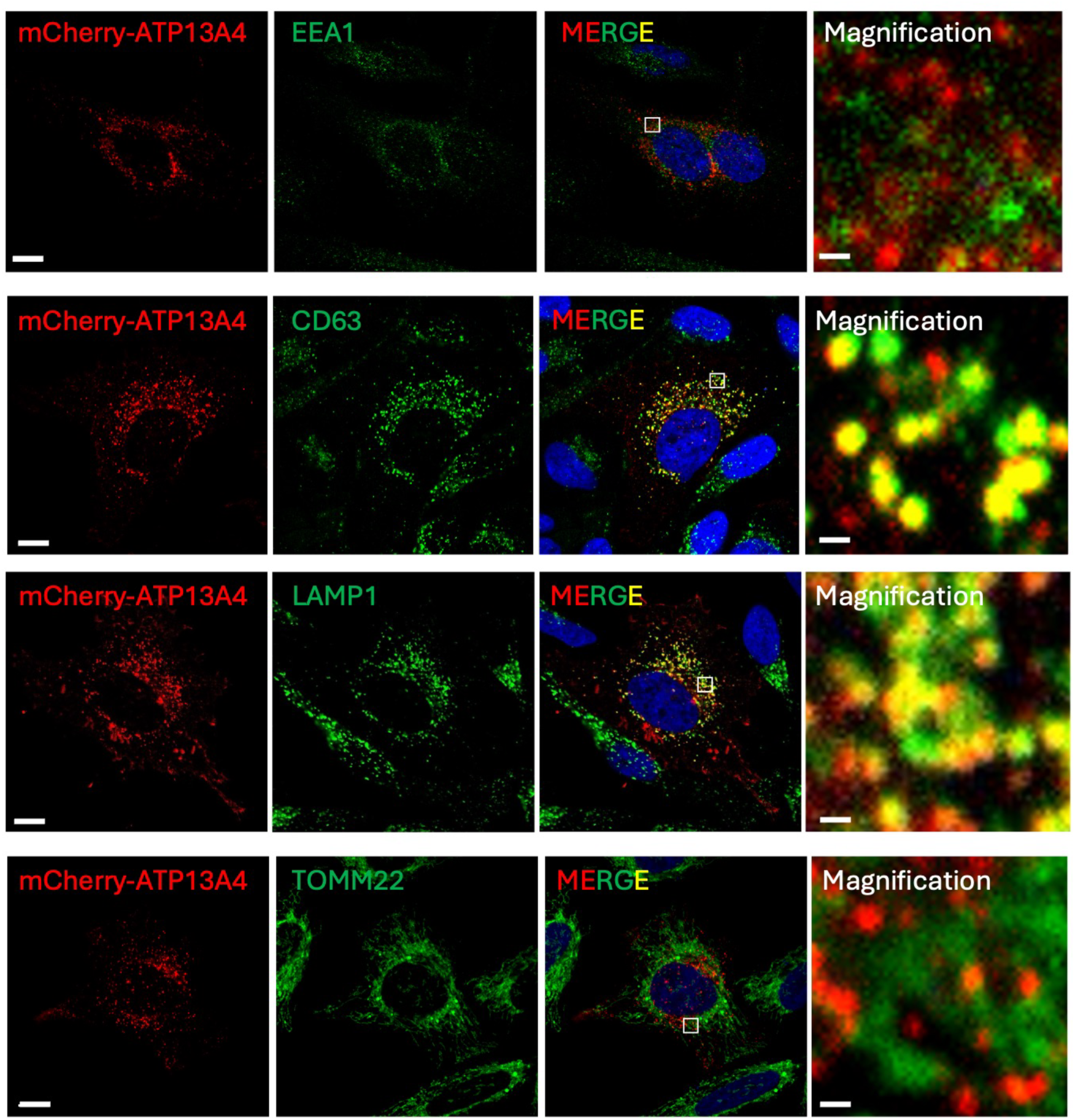
Atp13a4 localizes to late endo-/lysosomal compartments. Immunocytochemistry of HeLa cells transiently expressing N-terminal mCherry-labeled mouse Atp13a4 (msAtp13a4), co-stained with markers for specific organelles: EEA1 (early endosomes), CD63 (late endosomes), LAMP1 (lysosomes), and TOMM22 (mitochondria). Scale bar, 10 μm. Inset shows a magnification of the indicated white rectangular region (scale bar, 625 nm). *n* = 41-51 cells per condition from three independent experiments.

**Supplementary Fig. 4:**
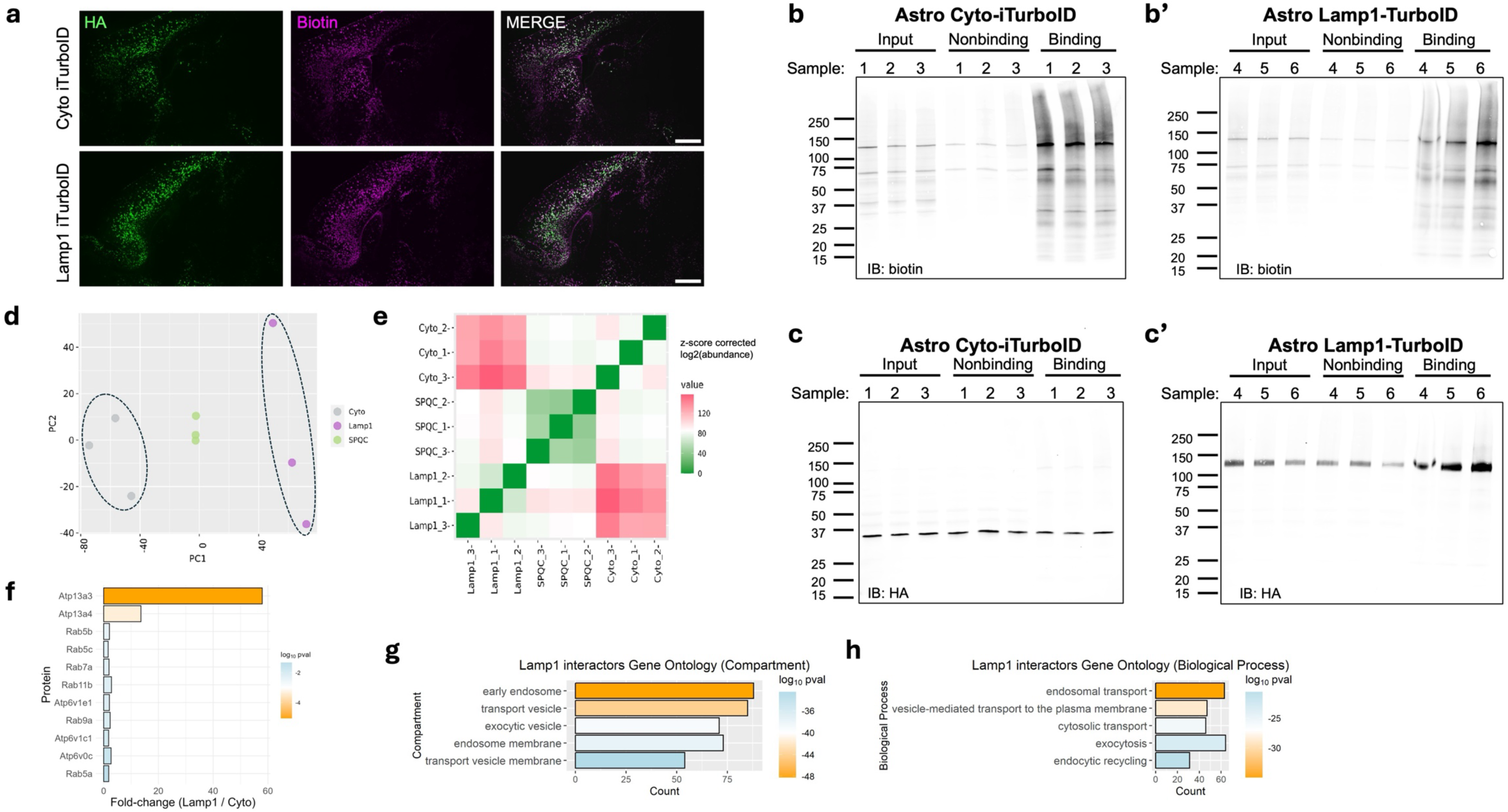
TurboID construct expression and biotinylation *in vivo.* **a**, Representative images of *in vivo* expression in the cortex of different TurboID constructs labeled with HA and the biotinylating activity labeled with streptavidin. Merged images show the colocalization of HA and biotin signals in astrocytes. Scale bar, 1000 μm. **b**-**c**, HA immunoblotting verifies expression of Astro-Cyto-TurboID (**b**) and Astro-Lamp1-TurboID (**b’**). Streptavidin blotting verifies biotinylation activity of Astro-Cyto-TurboID (**c**) and Astro-Lamp1-TurboID (**c’**) in cortical lysates (input) and subsequent immunoprecipitation (binding). **d**, Principal component analysis (PCA) of BioID data shows Astro-Lamp1-TurboID samples (purple) clustering separately from Astro-Cyto-TurboD samples (gray) and study pool QC (SPQC) samples (green). **e**, Heatmap displaying the correlation of protein abundance for all pairwise combinations of samples. Unsupervised hierarchical clustering shows Astro-Lamp1-TurboID samples clustering separately from Astro-Cyto-TurboID and SPQC samples. **f**, Bar graph depicting the detection ratio of Lamp1-TurboID versus Astro-Cyto-TurboID for putative polyamine transporters (Atp13a3 and Atp13a4) and selected endo-lysosomal proteins. (**g**-**h**) Bars show the top 10 most significant Gene Ontology (GO) terms, ordered by lowest adjusted p-value, for the proteins differentially detected by Astro-Lamp1-TurboID compared to Astro-Cyto-TurboID. (**g**) Cellular Compartment; (**h**) Biological Process.

**Supplementary Fig. 5:**
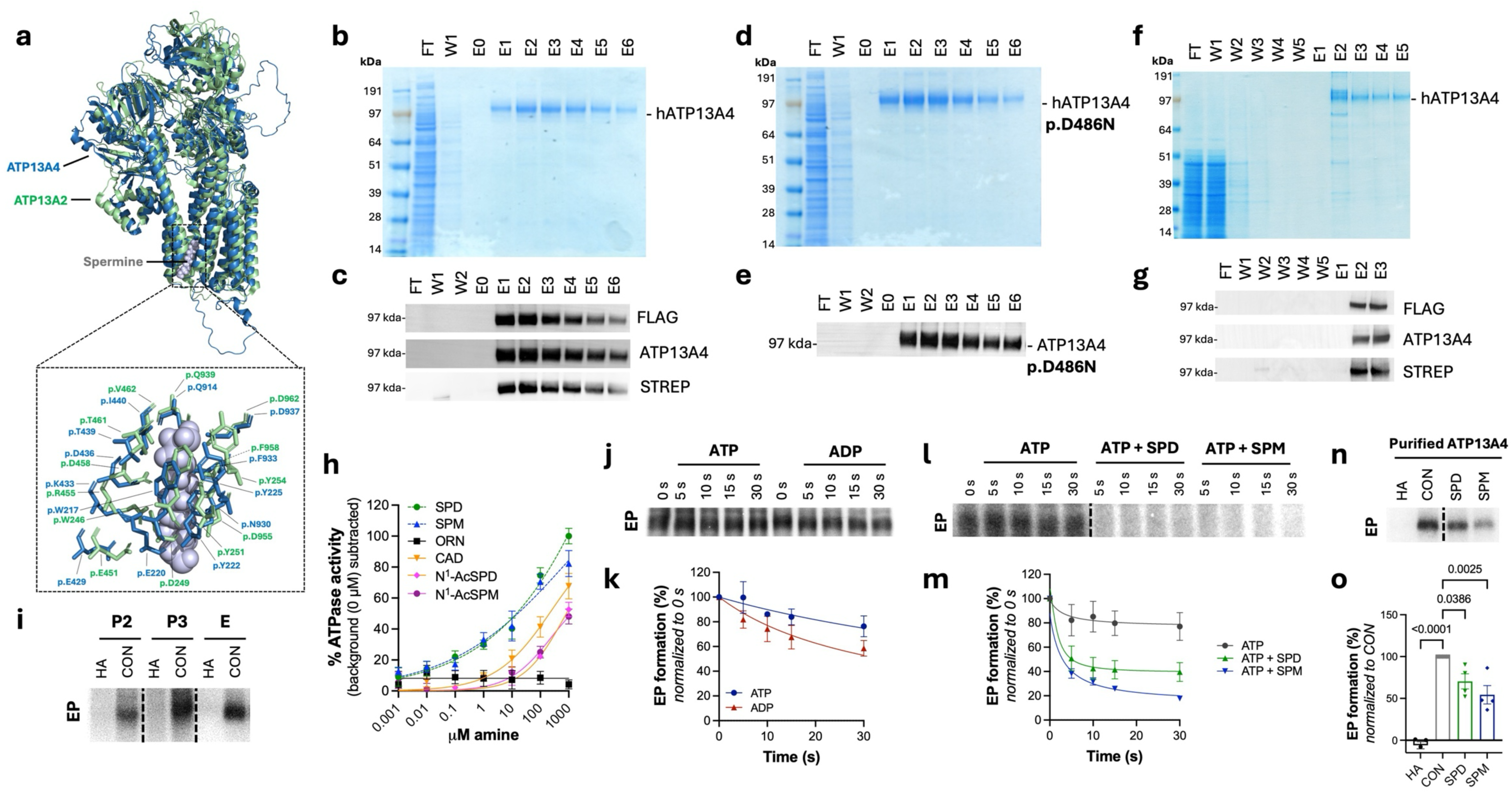
hATP13A4 spontaneously forms a phospho-enzyme that is sensitive to polyamines. **a**, Structural visualization of hATP13A4 (AlphaFold prediction) and hATP13A2 (PDB 7n78) using PyMOL. Top: overlay of hATP13A4 (blue) and hATP13A2 (green), with spermine shown in grey. Bottom: close-up view of the spermine binding pocket. **b**-**g**, Coomassie staining (**b**, **d**, **f**) and Western blot analysis (**c**, **e**, **g**) showing the purification process for WT hATP13A4 (**b**-**c**, **f**-**g**) or the p.D486N mutant (**d**-**e**), starting from detergent-solubilized HEK293T cell lysates (**b**-**e**) or yeast membranes (**f**-**g**), followed by streptavidin affinity chromatography and subsequent elution with biotin. *n* ≥ 3 independent purifications. FT, flow-through; W, wash; E, elution. **h**, Dose-response curves showing the effect of spermidine (SPD), ornithine (ORN), cadaverine (CAD), N^1^-acetylspermidine (N^1^-AcSPD) and N^1^-acetylspermine (N^1^-AcSPM) on the ATPase activity of solubilized purified ATP13A4 WT. A spermine (SPM) reference curve, derived from Fig. 2b, is included for comparison. *n* = 4 independent biological experiments for ORN, CAD, N^1^-AcSPD, and N^1^-AcSPM. For SPD, *n* = 4 at 1000 µM and *n* = 2 at the other concentrations. **i**, Representative autoradiogram showing phosphoenzyme (EP) formation of WT hATP13A4 in yeast membrane fractions (heavy membrane fraction P2 and light membrane fraction P3) and purified protein, which is sensitive to 0.3 M hydroxylamine (HA). **j**-**k**, Representative autoradiogram (**j**) and quantification (**k**) of pulse ([γ-^32^P]-ATP) chase (cold ATP or ADP) experiment to determine ATP and ADP sensitivity of hATP13A4 EP. *n* = 3 independent experiments. **l**-**m**, Representative autoradiogram (**l**) and quantification (**m**) of pulse ([γ-^32^P]-ATP) chase (cold ATP) experiment in the presence and absence of spermidine (SPD) or spermine (SPM) in yeast membranes. *n* = 4 (ATP + SPM) or 5 (ATP, ATP + SPM) independent experiments. **n**-**o**, Representative autoradiogram (**n**) and quantification (**o**) of purified ATP13A4 EP levels in the presence of 0.3 M HA, 1 mM SPD or SPM. *n* = 4 independent experiments. One-way ANOVA with Dunnett’s post-test.

**Supplementary Fig. 6:**
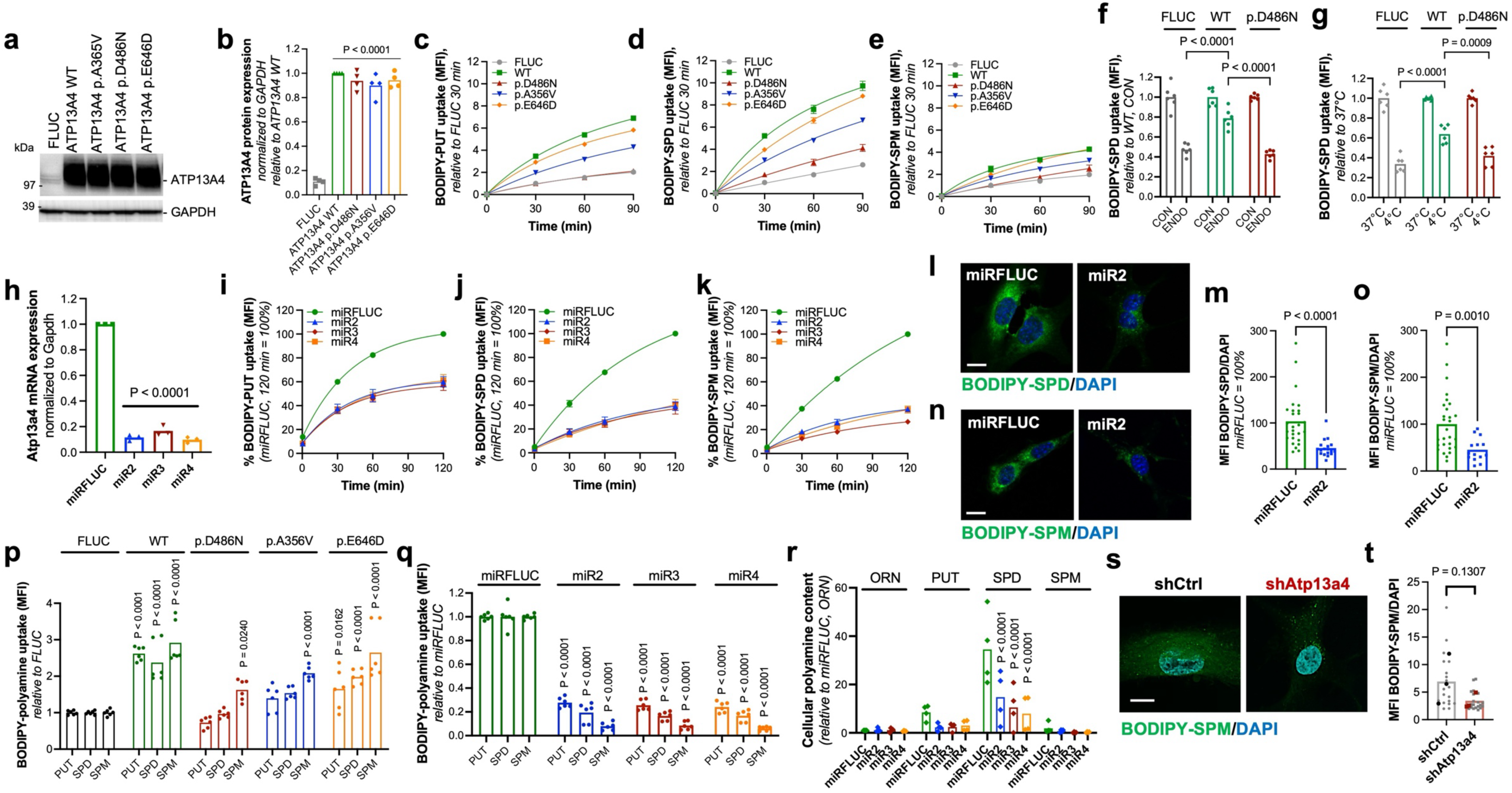
ATP13A4 promotes cellular polyamine uptake. **a**, Overexpression of firefly luciferase (FLUC), WT hATP13A4, the p.D486N mutant, or disease-associated variants p.A356V and p.E646D in H4 cells was confirmed by immunoblotting. GAPDH was used as a loading control. **b**, Densitometric quantification of hATP13A4 expression shown in **a**. *n* = 4 independent cultures. One-way ANOVA with Dunnett’s multiple comparisons test. **c-e**, Flow cytometric analysis of cellular BODIPY-putrescine (PUT, **c**),-spermidine (SPD, **d**) or-spermine (SPM, **e**) uptake in serum-containing medium. *n* = 3 independent experiments. **f**, Flow cytometric quantification of BODIPY-SPD in the presence or absence of endocytosis inhibitors (ENDO). Cells were pre-treated with a combination of endocytosis inhibitors (Dynasore, Genistein, Pitstop 2) prior to addition of BODIPY-SPD. **g**, Flow cytometric quantification of BODIPY-SPD uptake under low-temperature (4 °C) or physiological (37 °C) conditions. **f**-**g**, *n* = 3 independent experiments. Two-way ANOVA with Šídák’s multiple comparisons test. **h**, *msAtp13a4* mRNA expression following knockdown with lentiviral shRNA transduction (miR2-4) was assessed by qPCR. *n* = 3 independent experiments. One-way ANOVA with Dunnett’s multiple comparisons test. **i-k**, Flow cytometric analysis of cellular BODIPY-PUT (**i**),-SPD (**j**) or-SPM (**k**) uptake in serum-containing medium. *n* = 3 independent experiments. **l-o**, Confocal microscopy analysis of BODIPY-SPD (**l**, **m**) and-SPM (**n**, **o**) distribution. Scale bar, 10 μm. Mean fluorescence intensities (MFI) of BODIPY-SPD (**m**) and-SPM (**o**) corresponding to images in **l** (miRFLUC, 27 cells; miR2, 18 cells) and **n** (miRFLUC, 28 cells; miR2, 14 cells), respectively. *n* = 3 independent cultures. Mann-Whitney U test. **p**-**q**, Flow cytometric analysis of BODIPY-PUT,-SPD or-SPM uptake in H4 cells overexpressing FLUC, WT hATP13A4 or indicated variants (**p**) and in C8-D1A cells with *msAtp13a4* knockdown (**q**), performed in serum-free medium. **r**, Metabolomics of cellular polyamines in *msAtp13a4* knockdown cells (miR2-4) compared with C8-D1A control (miRFLUC). **p**-**r**, *n* = 3 independent experiments. Two-way ANOVA with Šídák’s multiple comparisons test. **s**, Confocal images of BODIPY-labelled SPM distribution in control (shCtrl) and *msAtp13a4* knockdown (shAtp13a4) primary astrocytes. Scale bar, 10 μm. **t**, Mean fluorescence intensities (MFI) of BODIPY-SPM (shCtrl, 19 cells; shAtp13a4, 20 cells). *n* = 3 independent cultures. Paired *t* test.

**Supplementary Fig. 7:**
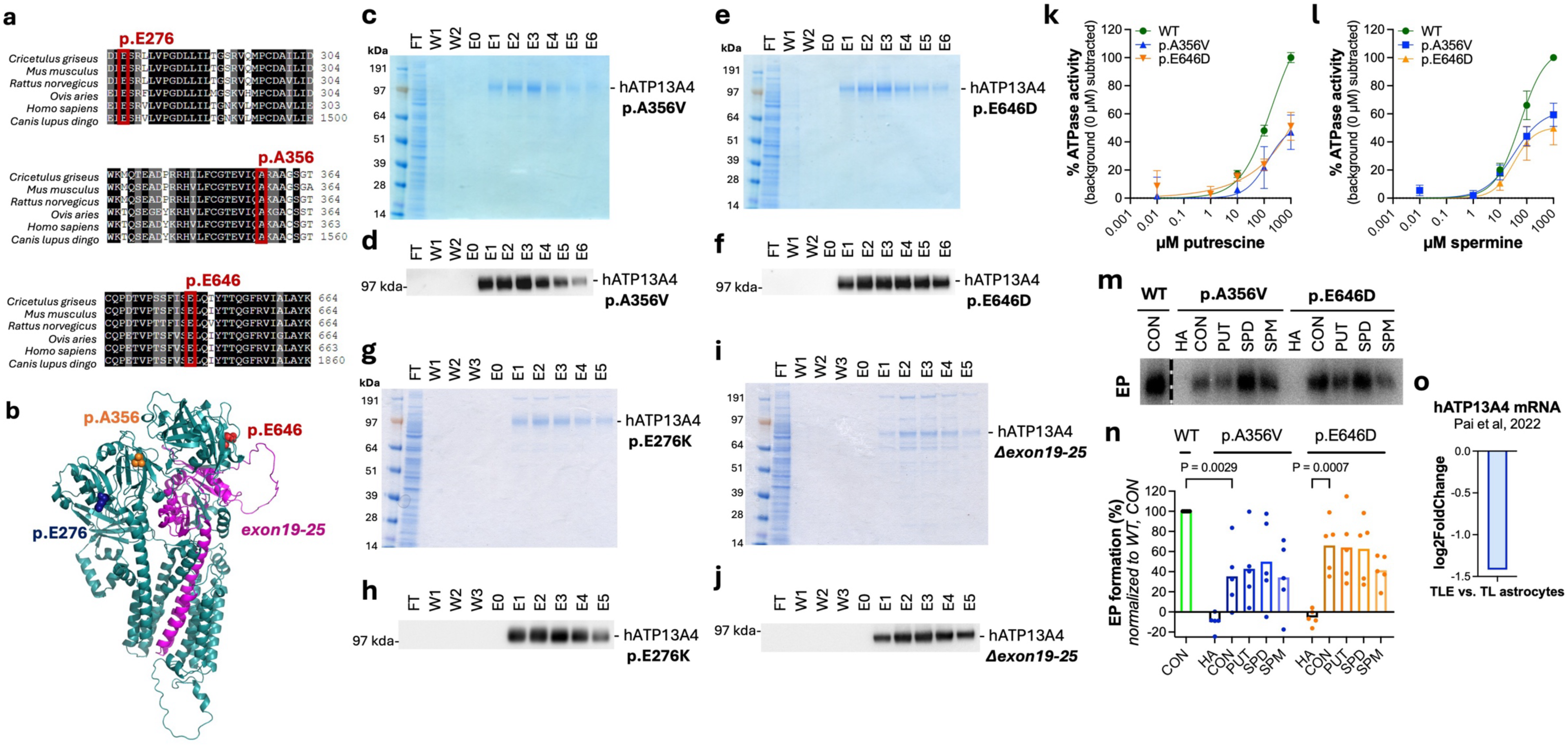
Disease-associated ATP13A4 variants. **a**, Sequence alignment showing disease-associated residues are highly conserved. The alignment was generated using Clustal Omega (149). **b**, Predicted 3D structure of ATP13A4 (AlphaFold) with disease-associated residues indicated. **c**-**j**, Coomassie staining (**c**, **e**, **g**, **i**) and Western blot analysis (**d**, **f**, **h**, **j**) showing the purification process for the disease-associated ATP13A4 variants p.A356V (**c**-**d**), p.E646D (**e**-**f**), p.E276K (**g**-**h**) and *Δexon19-25* (**i-j**) starting from detergent-solubilized HEK293T cell lysates, followed by streptavidin affinity chromatography and subsequent elution with biotin. *n* ≥ 3 independent purifications. FT, flow-through; W, wash; E, elution. **k**-**l**, Dose-response curve showing the effect of putrescine (**k**) and spermine (**l**) on the ATPase activity of solubilized purified ATP13A4 (WT or disease-associated p.A356V and p.E646D variants). *n* = 4 independent experiments. **m**-**n**, Phosphoenzyme (EP) levels of purified, solubilized ATP13A4 (WT and disease-associated variants p.A356V and p.E646D) under control (CON) conditions and in the presence of putrescine (PUT), spermidine (SPD), spermine (SPM), or hydroxylamine (HA). **m**, Representative autoradiogram. **n**, Quantification of EP in **m**. *n* = 5 independent experiments. Two-way ANOVA with Tukey’s multiple comparisons test. Individual data points are shown. **o**, Human RNA-seq data (from (105)) showing that *ATP13A4* is differentially downregulated in human epilepsy astrocytes [astrocyte temporal lobe epilepsy (TLE) *versus* astrocyte temporal lobe (TL) control].

**Supplementary Fig. 8:**
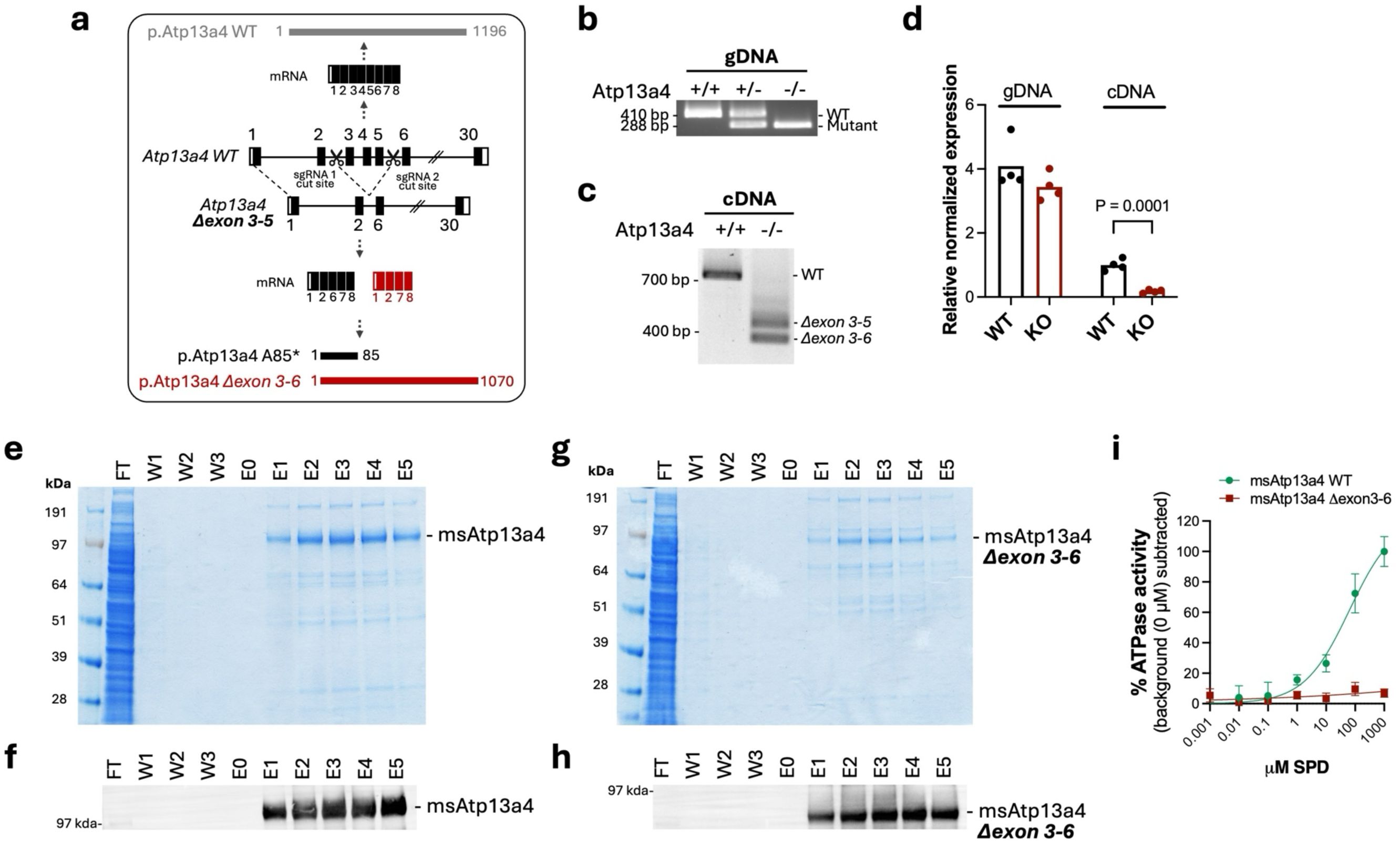
Validation of the *Atp13a4* knockout mouse model. **a**, Schematic representation of the CRISPR-Cas9-generated *Atp13a4* knockout allele, showing deletion of exons 3-5 and the resulting transcript variants. The deletion introduces a premature stop codon (Ala85*) and is predicted to generate a truncated protein lacking essential functional domains. An alternative splice variant lacking exon 6 (Δexon3-6) is indicated. **b**, Genomic PCR confirming deletion of exons 3-5 in homozygous *Atp13a4* KO mice compared to WT and heterozygous mice. **c**, RT-PCR analysis of cortical RNA showing loss of the full-length *Atp13a4* transcript and the presence of aberrant exon-deleted products in KO mice. Sanger sequencing of excised bands confirmed the expected exon 3-5 deletion and additionally identified a variant transcript lacking exon 6, indicating altered downstream splicing. **d**, qPCR analysis targeting exon 6 demonstrating reduced transcript levels in KO cDNA despite comparable genomic DNA levels, consistent with altered splicing and transcript instability. **e**-**h**, Purification of recombinant WT and Δexon3-6 msAtp13a4 proteins, visualized by Coomassie staining (**e-f**) and corresponding immunoblot analysis (**g-h**). **i**, ATPase activity assay showing robust spermidine-stimulated activity of WT msAtp13a4, whereas the Δexon3-6 variant displays no detectable stimulation, demonstrating loss of catalytic function.

**Supplementary Fig. 9:**
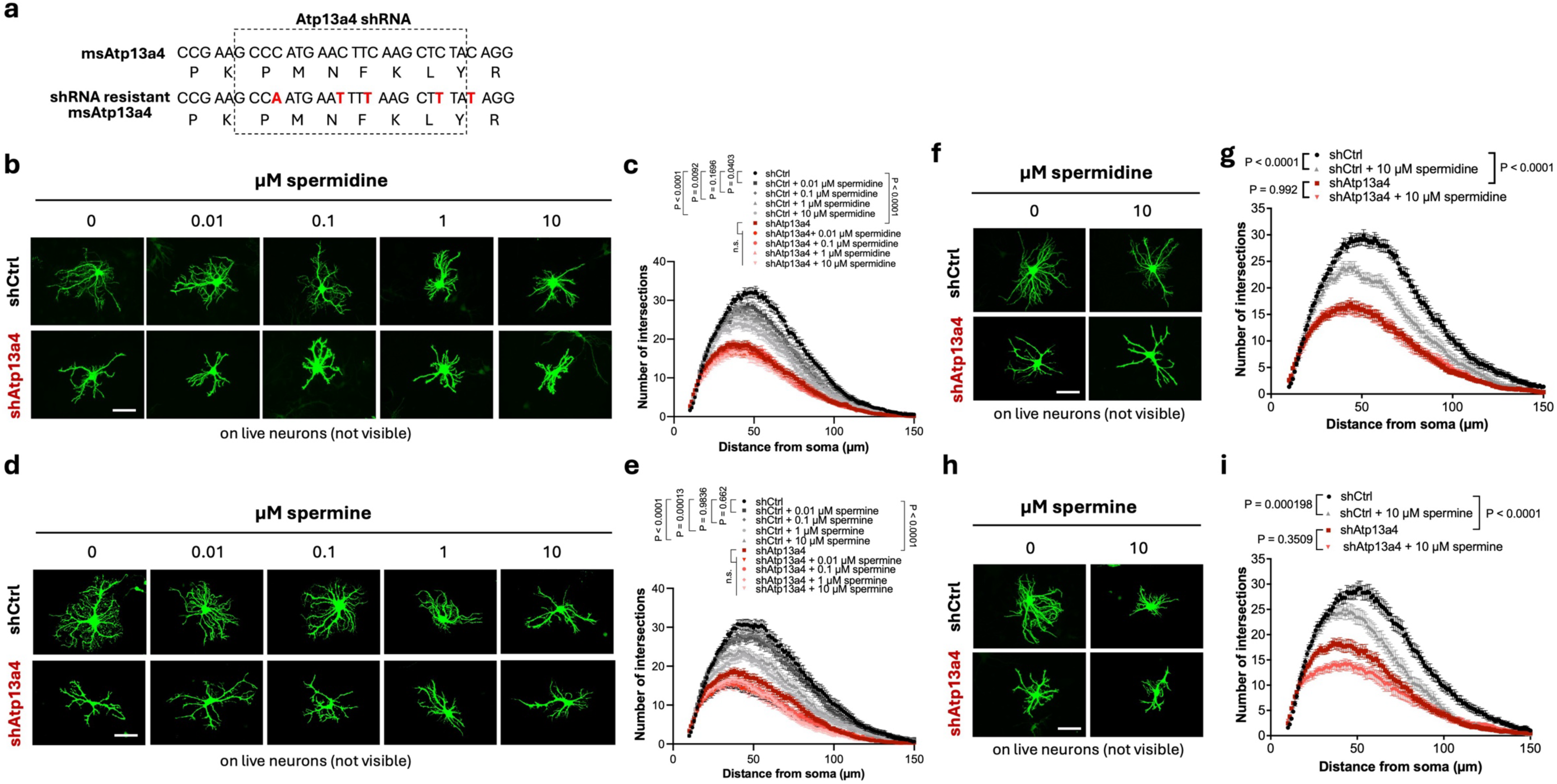
Polyamines decrease astrocyte complexity in an ATP13A4-dependent manner. **a**, shRNA-resistant ATP13A4 construct with silent mutations (red) in the shRNA target site to prevent knockdown. **b**, Representative images of astrocytes (green) transfected with shRNA targeting mouse/rat Atp13a4 (shAtp13a4) or a scrambled control (shCtrl), co-cultured with WT neurons (not labeled) for 48 h, and treated with spermidine during the final 8 h of co-culture. Scale bar, 50 μm. **c**, Quantification of astrocyte complexity via Sholl analysis for conditions in **b**. *n* = 46-68 astrocytes per condition from three independent experiments. **d**, Representative images of shRNA expressing astrocytes co-cultured with WT neurons for 48 h and treated with spermine during the final 8 h of co-culture. Scale bar, 50 μm. **e**, Quantification of astrocyte complexity for conditions in **d**. *n* = 50-60 astrocytes per condition from three independent experiments. **f**, Representative images of shRNA expressing astrocytes (green), treated with spermidine for 24 h and then co-cultured with WT neurons for 48 h. Scale bar, 50 μm. **g**, Quantification of astrocyte complexity for conditions in **f**. *n* = 60-63 astrocytes per condition from three independent experiments. **h**, Representative images of shRNA expressing astrocytes (green), treated with spermine for 24 h and then co-cultured with WT neurons for 48 h. Scale bar, 50 μm. **i**, Quantification of astrocyte complexity for conditions in **h**. *n* = 60 astrocytes per condition from three independent experiments. All data are presented as mean ± SEM. **c**, **e**, **g**, **i**, Mixed effects ANOVA with Tukey’s post-hoc test.

**Supplementary Fig. 10:**
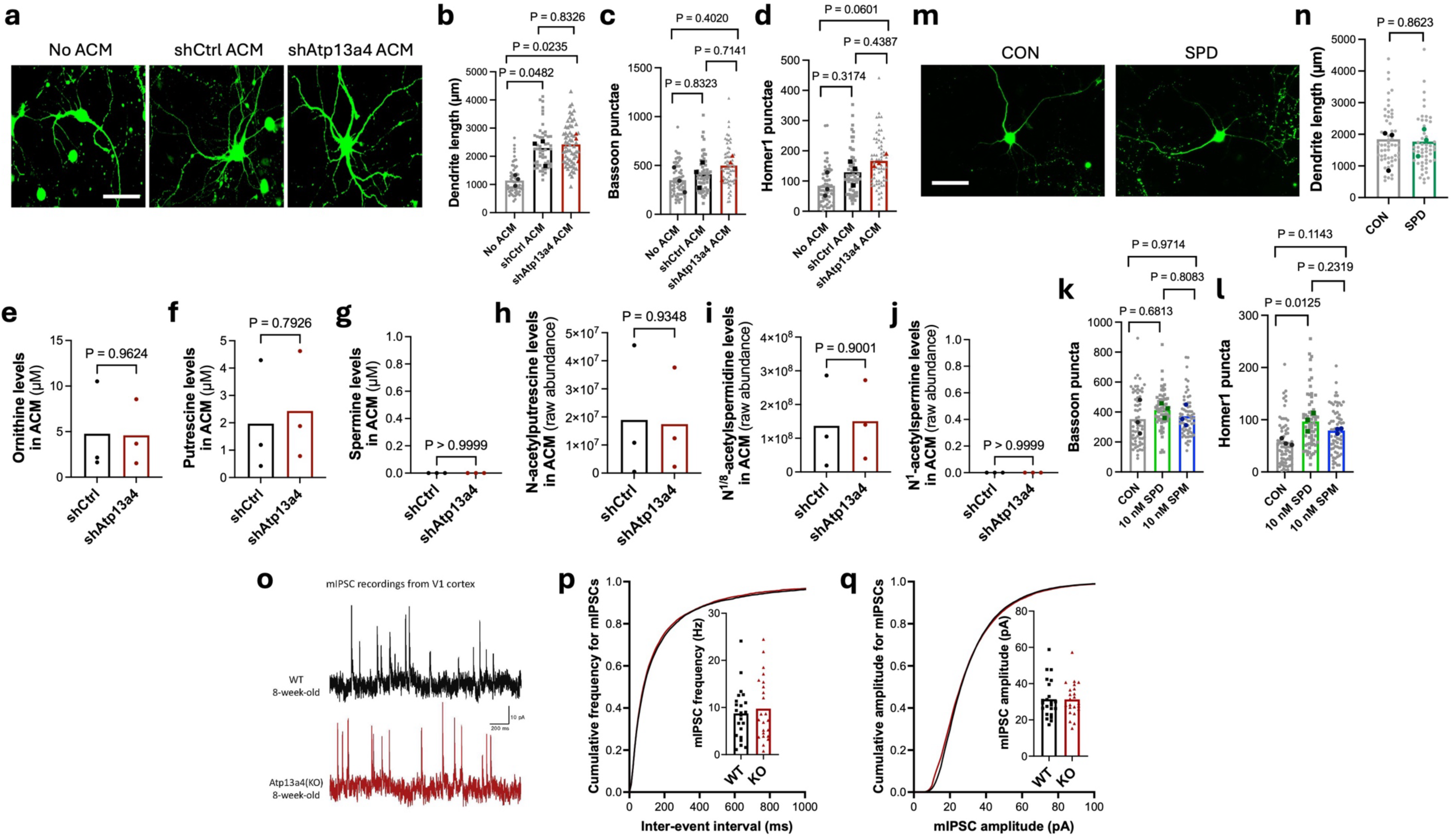
Atp13a4 knockdown in astrocytes does not impact neuronal morphology or inhibitory synapse function. **a**, Representative images of neurons transfected with GFP and treated with astrocyte-conditioned medium (ACM). Scale bar: 100 μm. **b**, Quantification of dendrite tree length in **a**. *n* = 52-60 cells per condition from three independent experiments. **c**-**d**, Quantification of Bassoon (**c**) and Homer1 (**d**) puncta of neuronal cultures treated with ACM (Fig. 4b), *n* = 58-64 cells per condition from three independent experiments. **b**-**d**, Individual data points are shown, with the average of each experiment plotted in grey (no ACM), black (shCtrl ACM), red (shAtp13a4 ACM). Nested one-way ANOVA with Tukey’s multiple comparisons test. **e**-**g**, Metabolomics analysis of ornithine (**e**), putrescine (**f**), spermine (**g**), N-acetylputrescine (**h**), N^1/8^-acetylspermidine (**i**), and N^1^-acetylspermine (**j**) levels in shCtrl ACM *versus* shAtp13a4 ACM. *n* = 3 independent experiments. Unpaired two-tailed *t* test, except for panels **g** and **j** where Mann-Whitney test was used. **k-l**, Quantification of Bassoon (**k**) and Homer1 (**l**) puncta of neuronal cultures treated with SPD or SPM (Fig. 4e). *n* = 64-66 cells per condition from three independent experiments. Individual data points are shown, with the average of each experiment plotted in grey (no ACM), green (SPD) or blue (SPM). Nested one-way ANOVA with Tukey’s multiple comparisons test. **m**, Representative images of neurons transfected with GFP and treated with spermidine (10 nM). Scale bar: 100 μm. **n**, Quantification of dendrite tree length in **m**. *n* = 52-54 cells per condition from three independent experiments. Individual data points are shown, with the average of each experiment plotted in grey (control, CON) or green (SPD). Nested *t*-test. **o**, Representative mIPSC (miniature inhibitory postsynaptic currents) traces from V1 cortex in acute brain slices of WT and *Atp13a4* KO mice. **p**, Quantification of frequency average (inset) and cumulative probability of mIPSC from *Atp13a4* WT and KO neurons. **q**, Quantification of amplitude average (inset) and cumulative probability of mIPSC from *Atp13a4* WT and KO neurons. **p**-**q,** *n* = 26 (WT), 23 (*Atp13a4* KO) neurons from 3 mice per genotype. Mann-Whitney U test (inset). Kolmogorov-Smirnov test (cumulative probability). Individual data points are shown.

**Supplementary Fig. 11.**
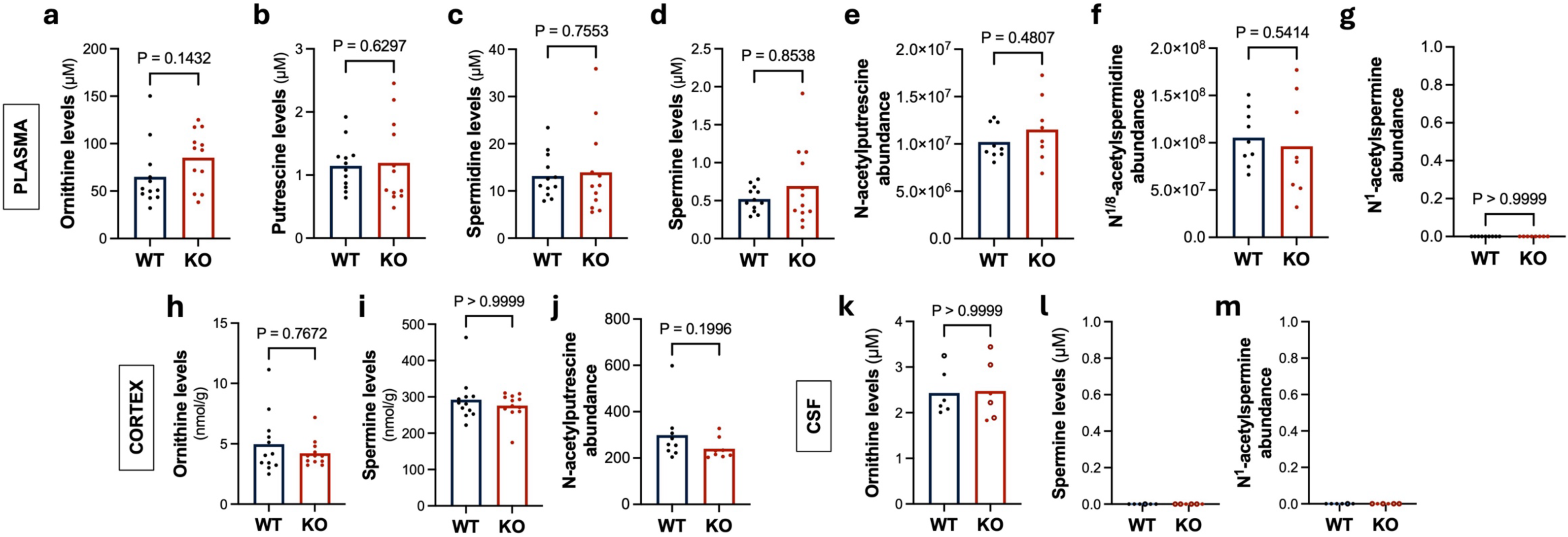
Extended metabolomic profiling of plasma, cortex, and CSF in two-month-old *Atp13a4* WT and KO mice. Targeted LC-MS/MS quantification of ornithine (**a**), putrescine (**b**), spermidine (**c**), spermine (**d**), N-acetylputrescine (**e**), N^1/8^-acetylspermidine (**f**) and N^1^-acetylspermine (**g**) in plasma from *Atp13a4* WT and KO mice. For **a**-**d**, *n* = 12 (WT) and 12 (KO) mice; for **e**-**g**, *n* = 9 (WT) and 8 (KO) mice. **h**-**j**, Quantification of ornithine (**h**), spermine (**i**), and N-acetylputrescine (**j**) in cortical tissue. For **h**-**i**, *n* = 12 (WT) and 12 (KO) mice; for **j**, *n* = 9 (WT) and 8 (KO) mice. **k**-**m**, Quantification of ornithine (**k**), spermine (**l**), and N^1^-acetylspermine (**m**) in cerebrospinal fluid (CSF). *n* = 6 biological samples per genotype. Filled symbols indicate single-animal samples; open symbols indicate pooled samples derived from 2-3 mice. Data are presented as mean, with individual data points representing individual mice (**a**-**j**) or biological samples (**k**-**m**). Statistical comparisons were performed using two-tailed Mann-Whitney U tests.

**Supplementary Fig. 12:**
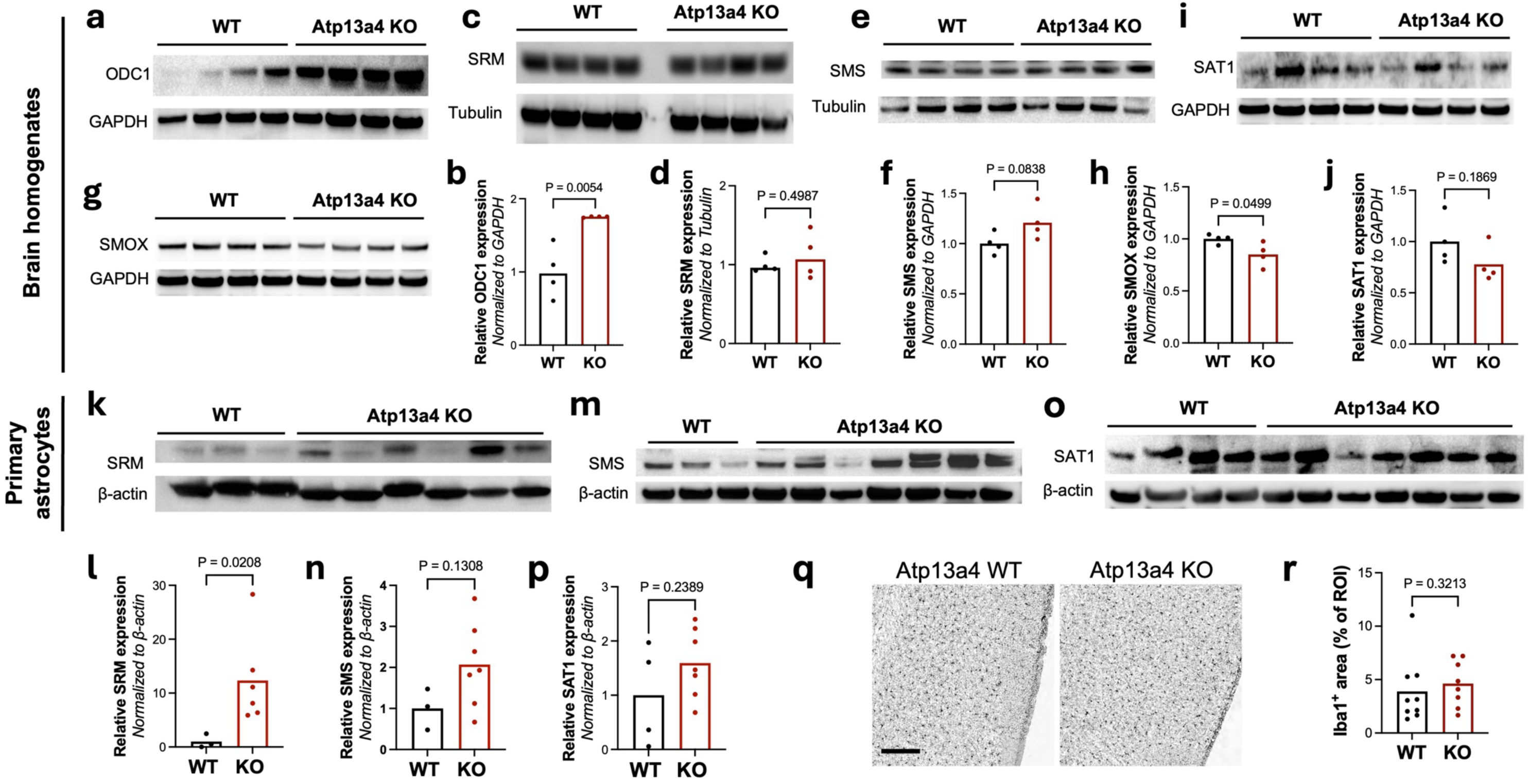
ATP13A4 loss induces selective adaptations in the polyamine pathway without evidence of microglial activation. **a**-**j**, Western blot analyses and corresponding quantifications of ODC1 (**a**-**b**), SRM (**c**-**d**), SMS (**e**-**f**), SMOX (**g**-**h**) and SAT1 (**i**-**j**) expression in brain homogenates from *Atp13a4* WT and KO mice. GAPDH and tubulin served as loading controls. **k**-**p**, Western blot analyses and corresponding quantifications of SRM (**k**-**l**), SMS (**m**-**n**) and SAT1 (**o**-**p**) expression in primary astrocytes from *Atp13a4* WT and KO mice. β-actin served as a loading control. Data are presented as mean, with individual data points representing independent biological replicates (**a**-**j**, individual mice; **k**-**p**, individual primary astrocyte preparations). Statistical comparisons were performed using unpaired two-tailed *t* tests. **q**, Iba1 immunostaining in cortex of two-month-old *Atp13a4* WT and KO mice. Scale bar, 50 µm. **r**, Quantification of cortical Iba1 immunoreactivity. Data points represent mouse averages. *n* = 8 (KO) and 9 (WT) mice, with 2 sections per mouse and 5 ROIs per section. Statistical comparisons were performed using an unpaired two-tailed *t* test.

**Supplementary Fig. 13:**
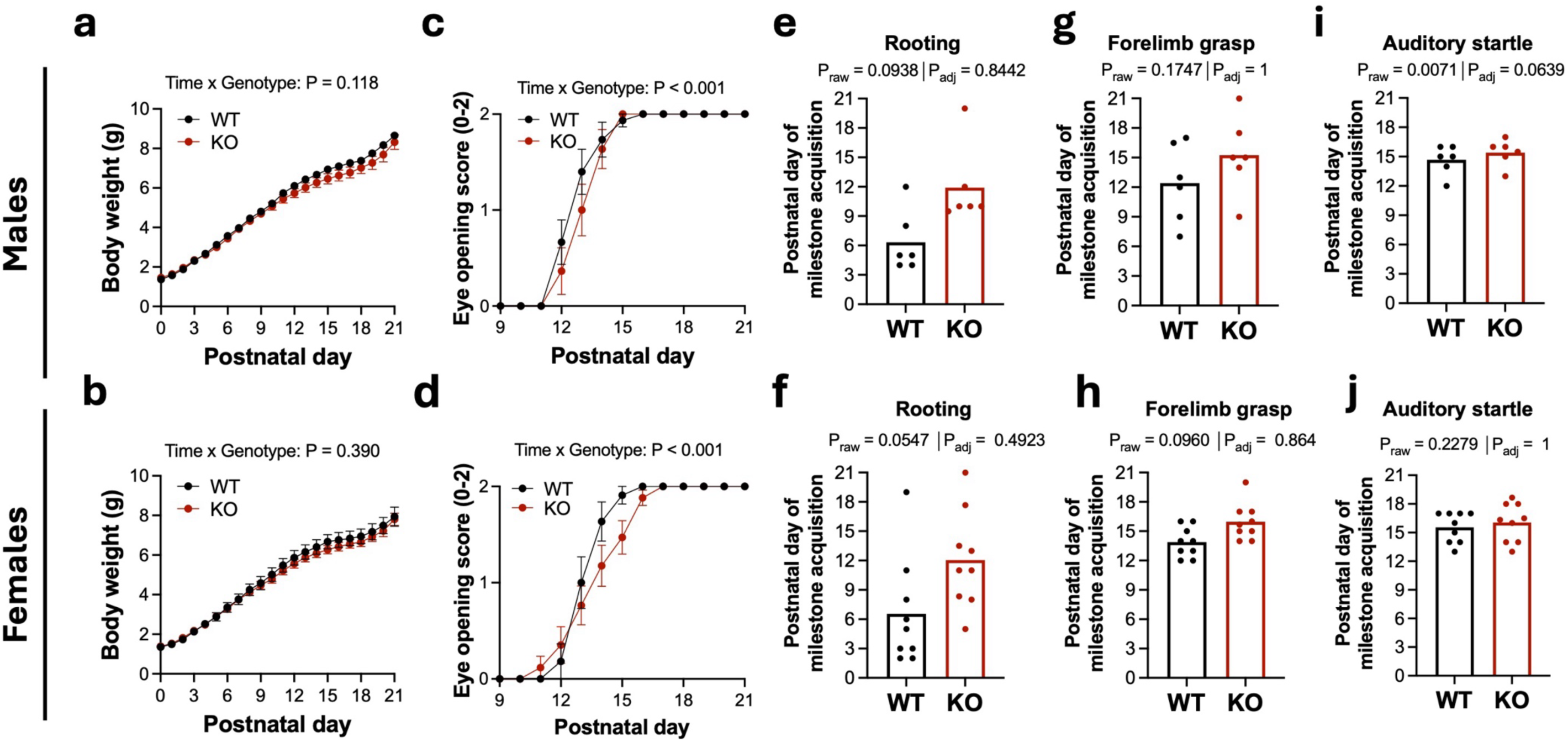
Early neurodevelopmental impairments in *Atp13a4* KO mice occur similarly in male and female pups. Developmental milestone acquisition (from Fig. 6a, b, f, h, j) was analyzed separately in male (**a**, **c**, **e**, **g**, **i**) and female (**b**, **d**, **f**, **h**, **j**) *Atp13a4* WT and KO littermates. **a**-**b**, Body weight progression over time. **c**-**d**, Eye opening acquisition. For **a**, *n* = 11 (WT, female), 15 (WT, male), 19 (KO, female) and 12 (KO, male) mice; for **b**, *n* = 11 (WT, female), 15 (WT, male), 17 (KO, female) and 11 (KO, male) mice; animals with unilateral eye development were excluded from this analysis. Statistical comparisons were performed using a linear mixed-effects model (**a**) and a cumulative link mixed model (**b**). **e**-**f**, Rooting. **g**-**h**, Forelimb grasp. **i**-**j**, Auditory startle. Data are presented as mean ± SEM (**a**-**d**) or as mean with individual data points representing litter-level genotype averages from matched litters. For milestone analyses, acquisition is defined as the second consecutive day on which pups met the predefined criterion. Statistical comparisons were performed using paired two-tailed *t* tests (**g**-**j**) or Wilcoxon matched-pairs signed-rank tests (**e**-**f**), as appropriate based on normality. P-values were adjusted for multiple comparisons across the nine milestone tests using Bonferroni correction (adjusted threshold α = 0.0056; or equivalently, raw p-values multiplied by 9 and compared to 0.05).

**Supplementary Fig. 14:**
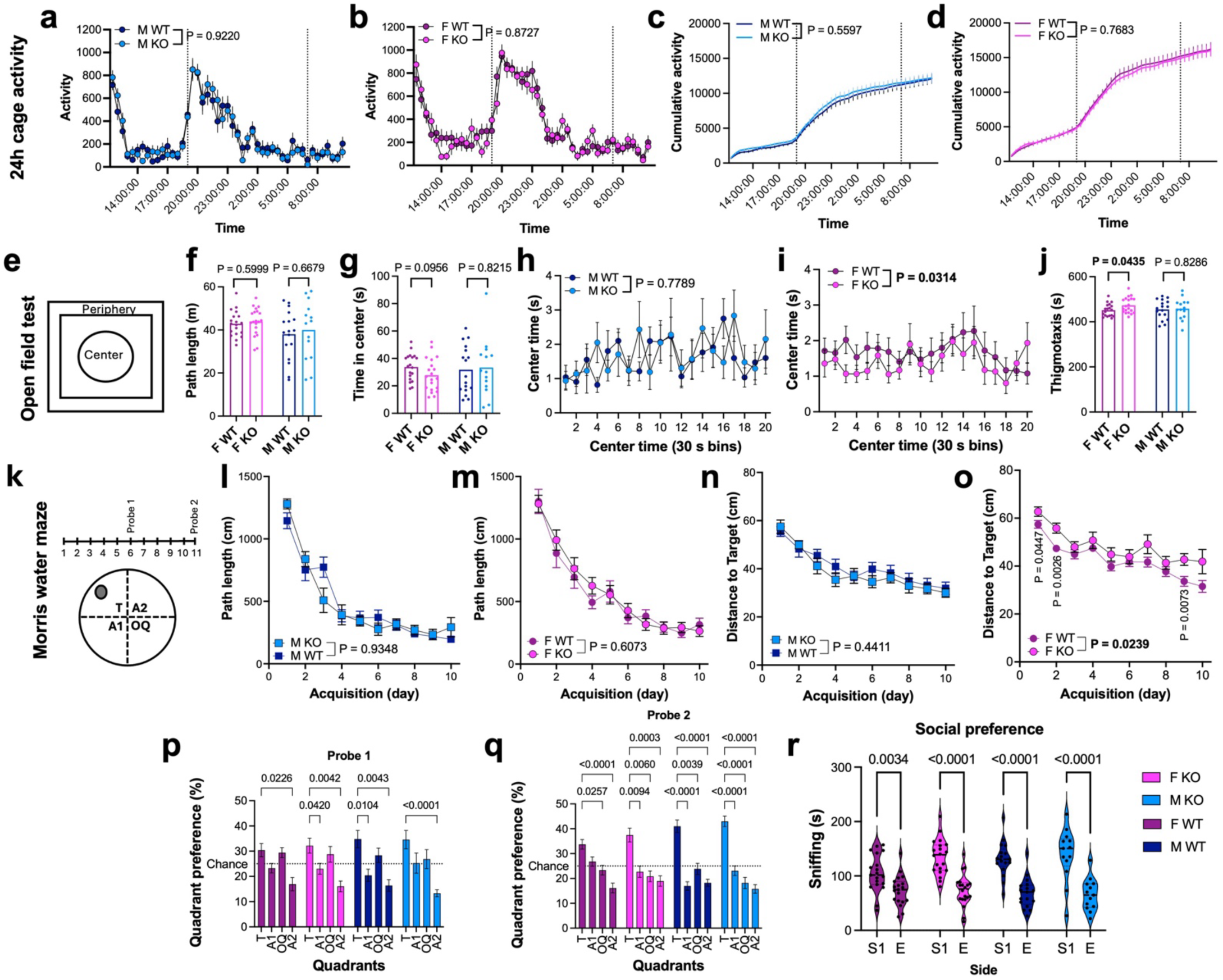
Behavioral characterization of two-month-old *Atp13a4* KO mice reveals mild, female-biased differences. **a**-**d,** 24 h home-cage activity monitoring. Cage activity over 24 h in male (**a**, **c**) and female (**b**, **d**) mice. Beam breaks are recorded in 30 min bins and plotted as time-resolved activity (**a**-**b**) or cumulative activity (**c**-**d**). Dotted vertical lines indicate lights off (at 19:00) and lights on (at 07:00). No genotype-dependent differences were observed in overall locomotor activity or circadian patterns. **e**-**j**, Open field test. **e**, Schematic of the arena indicating center and periphery zones. **f**, Total distance traveled (path length), showing no differences between genotypes. **g**, Time spent in the center of the arena. **h-i**, Center time analyzed in 30 s time bins for male (**h**) and female (**i**) mice. A global nonlinear regression tested whether a single curve fit all datasets (null) or if separate curves were required (alternative). Female KO mice show altered center exploration dynamics, whereas male mice are unaffected. **j**, Time spent in the periphery (thigmotaxis). Female KO mice display increased thigmotaxis, consistent with subtle alterations in exploratory behavior. **k**-**q**, Morris water maze test. **k**, Schematic representation of the Morris water maze (MWM) with 10 training days and probe trials on day 6 and day 11. T, the target quadrant where the hidden platform is normally located; A1 and A2, the adjacent quadrants that lie on either side of the target quadrant; and OQ, the opposite quadrant, which is diagonally across from the target. **l**-**m,** Total distance traveled (path length) to locate the hidden platform during acquisition in male (**l**) and female (**m**) mice. **n-o,** Distance to the hidden platform during acquisition across training days in male (**n**) and female (**o**) mice. Female KO mice show increased distance during acquisition, indicating a mild learning delay, whereas male mice are unaffected. **p**-**q,** On day 6 (**p**) and day 11 (**q**), the platform was removed from the pool and the amount of time spent in each quadrant was recorded. Dotted line is chance level (25 %). All groups show comparable target quadrant preference, indicating intact memory retention. **r**, Three-chamber social preference test. Violin plots represent the distribution of time spent in the sniffing zones (Stranger mouse 1, S1; empty, E) during the first 5 min of the test. All groups display normal social preference behavior. Data are presented as mean ± SEM, except in **f**, **g**, **j** and **r** where individual data points are shown. Statistical comparisons were performed using unpaired two-tailed *t* test (**f, g, j**), mixed-effects analysis (REML) with Tukey’s (**a**-**d**) or Dunnett’s (**l**-**o**) multiple comparisons test, two-way RM ANOVA with Dunnett’s post-test (**p**, **q**), and two-way ANOVA with Tukey’s post-test (**r**).

**Supplementary Fig. 15:**
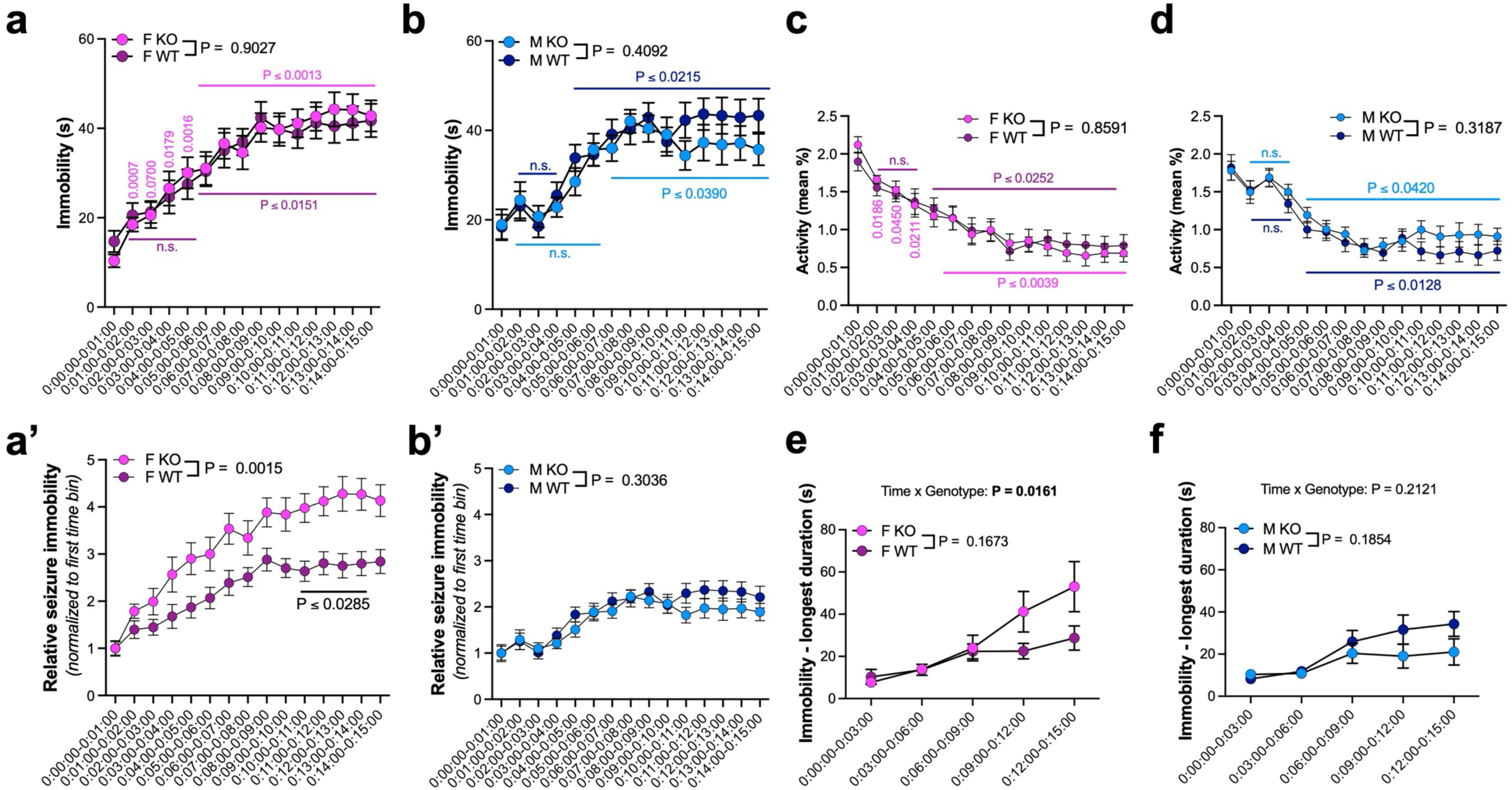
Female-biased alterations in PTZ-induced immobility dynamics in two-month-old *Atp13a4* KO mice. **a**-**b**, Immobility time following PTZ (pentylenetetrazol) challenge. Immobility duration is analyzed in 60 s time bins in female (**a**) and male (**b**) mice. Female *Atp13a4* KO mice exhibit an earlier increase in immobility compared to WT, whereas male mice show no genotype-dependent differences. **a’-b’**, Normalized immobility (same dataset as **a**-**b**). Immobility is normalized to each mouse’s baseline (*i.e.* first time bin) and plotted over time in female (**a’**) and male (**b’**) mice. This normalization highlights differences in the temporal onset of immobility independent of baseline activity levels. Female KO mice show an earlier increase compared to WT, whereas males are unaffected. **c**-**d**, Locomotor activity following PTZ injection. Activity is expressed as mean % over time in female (**c**) and male (**d**) mice. While overall activity levels are comparable between genotypes, female *Atp13a4* KO mice exhibit an earlier decline in activity following PTZ injection, consistent with an earlier transition into immobility. Male mice show no genotype-dependent differences. **e**-**f**, Longest immobility episode. Duration of the longest immobility bout is analyzed in 3 min time bins in female (**e**) and male (**f**) mice. A significant genotype × time interaction is observed in females but not in males, indicating altered temporal progression of immobility. Data are presented as mean ± SEM. Statistical comparisons were made using mixed-effects analysis (REML) with Dunnett’s (**a**-**b**) or Šidák’s (**c-d**) multiple comparisons test, two-way RM ANOVA with Šidák’s post-test (**a’-b’, e-f**).

**Supplementary Fig. 16:**
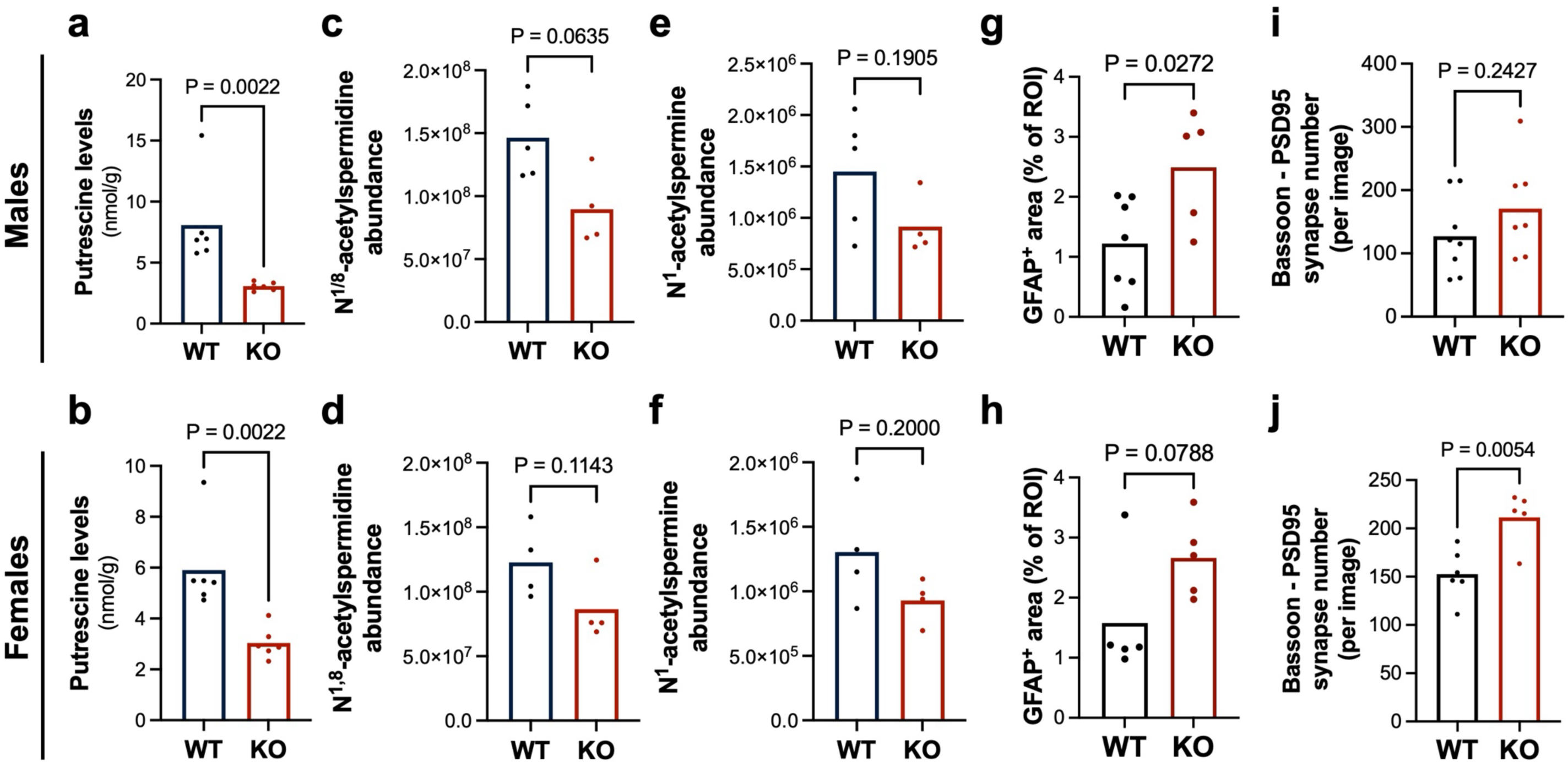
Polyamine homeostasis and astrocyte reactivity are similarly altered across sexes, while excitatory synapse density shows sex-dependent differences in two-month-old *Atp13a4* KO mice. **a**-**f**, Targeted LC-MS/MS quantification of polyamine species in cortical tissue (from Fig. 5a-d) was analyzed separately in male (**a**, **c**, **e**) and female (**b**, **d**, **f**) *Atp13a4* WT and KO mice. Data are presented as mean, with individual data points representing biological replicates (individual mice). Polyamine levels are comparably altered across sexes. **g**-**h**, Quantification of astrocyte reactivity based on GFAP immunostaining (from Fig. 5m-n) was analyzed separately in male (**g**) and female (**h**) mice. GFAP immunoreactivity is increased in *Atp13a4* KO mice relative to WT controls in both sexes. **i**-**j**, Quantification of excitatory synapse numbers (Bassoon and PSD95 co-localization) in V1 cortex (from Fig. 5o-p) was analyzed separately in male (**i**) and female (**j**) *Atp13a4* WT and KO mice. Data points represent mouse averages (2 sections/mouse, 3-4 images/section). Statistical comparisons were performed using two-tailed Mann-Whitney test (**a**-**f**) and unpaired two-tailed *t* test (**g**-**j**).

