## Supplementary material for "ATP13A4 gates extracellular polyamine levels to control excitatory synaptogenesis": Table S1

**Table S1. Oligonucleotide list.**

| GENE | SEQUENCE | ADDITIONAL INFORMATION |
|---|---|---|
| *msAtp13a4* | (F): 5’-CCAGCATGCTTTACTCAATG-3’<br>(R): 5’-GAAGATGGATCCAATGAGAC-3’ | Primers used for RT-qPCR analysis of C8-D1A cells |
| *msGapdh* | (F): 5’-TGTGTCCGTCGTGGATCTGA-3’<br>(R): 5’-CCTGCTTCACCACCTTCTTGA-3’ | Primers used for RT-qPCR analysis of C8-D1A cells |
| shAtp13a4 | 5’-GCCCATGAACTTCAAGCTCTA-3’ | shRNA sequence targeting *msAtp13a4* and *rAtp13a4* for knockdown |
| shCtrl | 5’-GCTCACCACGTGCAATACTAT-3’ | scrambled shRNA sequence used as a negative control |
| miRFLUC | 5' -GCGCTGAGTACTTCGAAATGTC- 3' | shRNA sequence targeting FLUC (firefly luciferase) used as a negative control in C8-D1A cells |
| miR2 | 5’-GCCAATATTTCAGCAGCTTATT-3’ | shRNA sequence targeting *msAtp13a4* for knockdown in C8-D1A cells |
| miR3 | 5’-AGCCCATGAACTTCAAGCTCTA-3’ | shRNA sequence targeting *msAtp13a4* for knockdown in C8-D1A cells |
| miR4 | 5’-GCGATACCTAAACAGTTATCAA-3’ | shRNA sequence targeting *msAtp13a4* for knockdown in C8-D1A cells |
| *msAtp13a4* | (F): 5’-GGCAGCCCACCTATACAAACTATATAT-3’<br>(R): 5’-GAATGAAAAGACATACGCCCATCT-3’ | Primers used for RT-qPCR analysis of primary astrocytes |
| *18S* | (F) 5′-GCAATTATTCCCCATGAACG-3′<br>(R) 5′-GGCCTCACTAAACCATCCAA-3′ | Primers used for RT-qPCR analysis of primary astrocytes |
| U6 | (F): 5′-GGACTAGTCAGGCCCGAAGGAATAGAAG-3′<br>(R): 5′-GGACTAGTGCCAAAGTGGATCTCTGCTG-3′ | Used for amplifying hU6 promoter and shRNA from pLKO.1-shRNA-EGFP |
| Atp13a4 KO/WT Fwd (*Atp(50715) CF*) | 5′-GGTCCCAGAATTCCTTGGCA-3′ | Genotyping |
| Atp13a4 WT Rv (*Atp(50715) WTR* ) | 5′-CAGTGCTGACAGGGAGATCC-3′ | Genotyping |

| | | |
|---|---|---|
| Atp13a4 KO Rv (*Atp(50715) KOR* ) | 5′-GCCTCACACTGCACTCTTCT-3′ | Genotyping |
| *msAtp13a4* (EXON1-7) | (F): 5′-ACCTTGAGAAGAGCCAGCAT-3′ (R): 5′-GAGGTCATACACGGTCAAAGC-3′ | Primers used for RT-PCR analysis of cortical RNA |
| *msAtp13a4* (EXON6) | (F): 5′-GAGGTTAATATGTGGGCCTA-3′ (R): 5′-ACCTCCTTGATGAGCAGTTTC-3′ | Primers used for RT-qPCR analysis of cortical RNA (exon 6 validation) |
| *msGapdh* | (F): 5′-TGTGTCCGTCGTGGATCTGA-3′ (R): 5′-CCTGCTTCACCACCTTCTTGA-3′ | Normalization reference for RT-qPCR analysis of cortical RNA (exon 6 validation) |
